# Application of Mendelian randomization to appraise causality in relationships between the gut microbiome and cancer

**DOI:** 10.64898/2026.01.05.26343428

**Authors:** Amy C. Dawes, Charlie Hatcher, Alec McKinlay, Kaitlin H. Wade

## Abstract

Cancer is among the leading causes of death worldwide; however, with incidence rising, there is a requirement to identify novel risk factors and biomarkers to prevent and diagnose cancer earlier. Previous work has implicated a role of the gut microbiome in cancer aetiology; however, its causal relevance is not clear. Whilst Mendelian randomisation (MR) is increasingly being applied to assess the causal role of the gut microbiome on several health outcomes, the complexities and limitations of the method are not fully addressed in much of the current literature. Here, we aimed to appropriately apply MR to understand the role of the gut microbiome in cancer aetiology, whilst demonstrating the range of sensitivity analyses required to provide appropriate causal inference. Whilst findings initially suggested that there were several bacterial genera, families and phyla that may alter cancer risk, further sensitivity analyses indicated that these results were unlikely to reflect causality given violations of core MR assumptions. This study highlights the need for a rigorous MR analysis pipeline, incorporating sensitivity analyses evaluating the plausibility of MR-derived effect estimates, to avoid incorrect conclusions around causality, where estimates likely reflect more complex relationships.

## INTRODUCTION

Cancer is a major public health burden, with ∼167,000 cancer deaths reported annually in the UK (2017–2019). Breast, prostate, lung and bowel cancers account for ∼53% of incident cases. Established risk factors include age, family history, genetics, and lifestyle and environmental exposures.^1^ Around four in ten cancers are estimated to be preventable via modification of known exposures, suggesting additional modifiable causal factors remain to be identified. Identifying such factors is important for informing interventions to reduce cancer incidence.^1^

The human gut microbiome is important in the regulation of human health^2^, with roles in host metabolism,^3,4^ inflammation,^5,6^ and immune response to both commensal and pathogenic microbes.^5,7–11^ Consequently, multiple studies have implicated the gut microbiome in carcinogenesis^12^ However, findings are inconsistent and inconclusive regarding causality,^13–24^ with limited evidence of replication across studies or between model organisms.^20^

Mendelian randomization (MR) can strengthen causal inference by utilizing human germline genetic variation (usually, single nucleotide polymorphisms [SNPs]) as instruments for an ideally modifiable trait in a manner that is analogous to randomized controlled trials (RCTs).^23,25,26^ Genetic variants robustly associated with microbial traits can be used to estimate their effect on cancer (Figure 1).^26–28^ As genotype is fixed at conception, MR is expected to be less susceptible to confounding and reverse causation than conventional observational studies, and can be applied where randomized trials are infeasible.^26–28^

**Figure 1.**
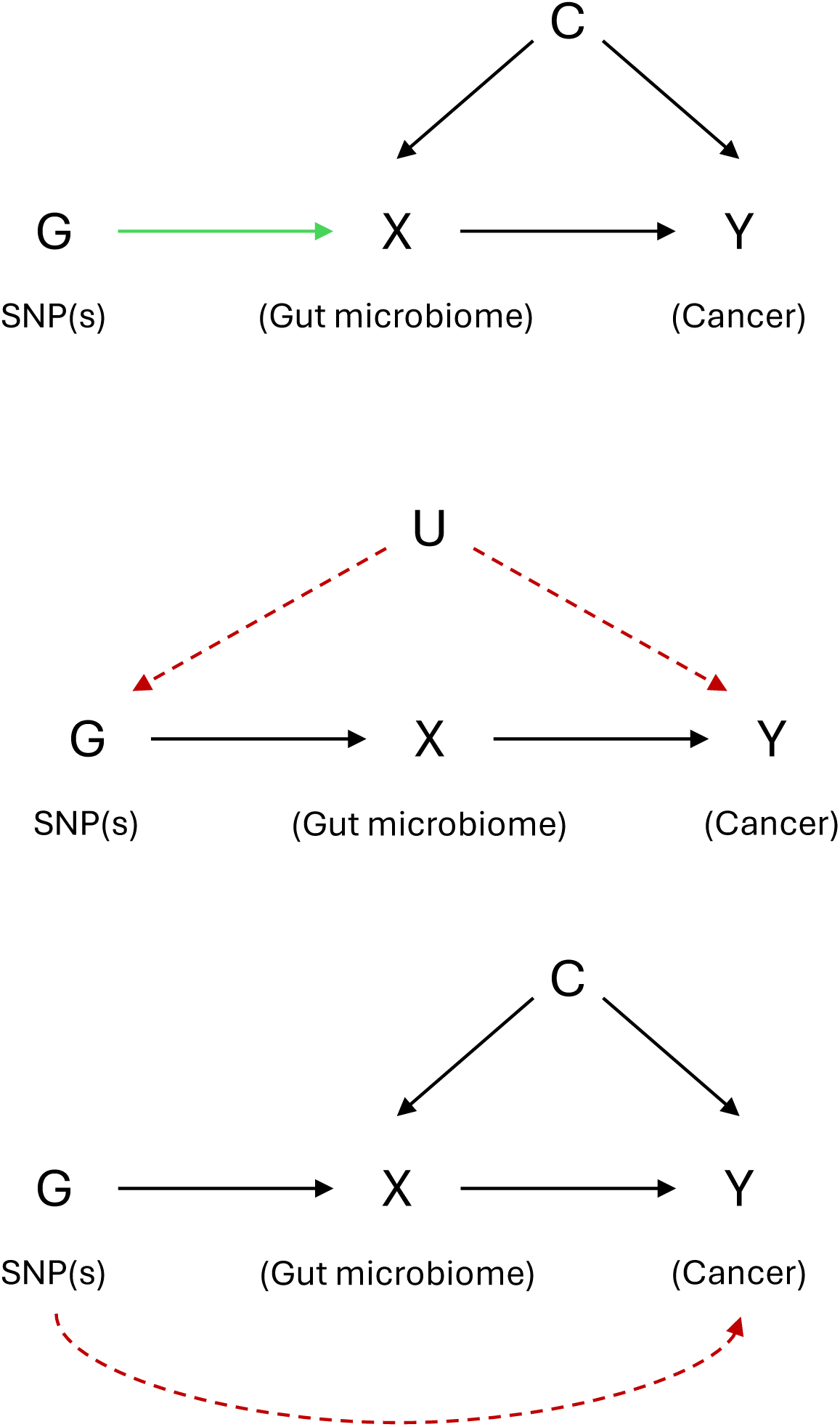
Core assumptions of MR when testing the relationship of the gut microbiome on cancer. Mendelian randomization relies on three core assumptions: (1) genetic variants used as instruments are robustly associated with the gut microbial trait of interest (relevance); (2) these variants are not associated with confounders of the microbiome–cancer relationship (independence); and (3) the variants influence cancer risk only through their effect on the gut microbiome and not via alternative pathways (exclusion restriction).^26,27^ In microbiome MR studies, violations of these assumptions—particularly due to pleiotropy and complex host genetic effects—may bias causal inference and require careful evaluation through sensitivity analyses.^14,20^

Two-sample MR using GWAS summary statistics has increasingly been applied to microbiome-related questions.^14,20,29–32^ However, there is a considerable lack of caution when interpretating causal estimates in the current literature, despite the established complexity of the host genetic effects on the gut microbiome.^14,20,23,29^ Building on our previous work, we aimed to use two-sample MR to estimate the causal relationship between the gut microbiome and cancer using some of the largest GWASs to date.^14,32^ We focused on cancers previously linked to the gut microbiome and with large GWAS available (i.e., cancers of the lung, breast, colorectum, prostate, endometrium, ovary, pancreas and oesophagus).^23,33–46^ Critically, we applied a structured set of sensitivity analyses to evaluate robustness to violations of MR assumptions and to contextualize causal claims.^14,23,32,33^

## RESULTS

### Main analysis

We performed MR analysis of 41 microbial traits using associated genetic variants identified by Hughes et al.^32^ (14 microbial traits) and the MiBioGen consortium (27 microbial traits),^14^ separately, on cancers of the lung, breast, colorectum, prostate, pancreas, endometrium, ovaries and oesophagus. Where previous work had performed these analyses using 14 microbial traits and the same methodology as presented here, these results are not reported.^23,45–47^

Across outcomes, 6–14 SNPs (Hughes et al.^32^) associated with 6-14 traits and 12–21 SNPs (MiBioGen^14^) associated with 18-30 traits were available after harmonisation (Supplementary Tables 1–3). F-statistics ranged from 21.4–56.2 (Hughes^32^) and 29.4– 88.4 (MiBioGen^14^), consistent with instrument strength for MR. SNPs were excluded if they were not available in the outcome cancer GWAS, no proxy was found or if they could not be harmonized (e.g., due to ambiguous MAFs for “palindromic” SNPs; Supplementary Table 3).

Of the 14 microbial traits identified in the Hughes et al.^32^ GWAS and four cancers analysed, there was evidence for five causal effects across four microbial traits and three cancer outcomes. Specifically, the bacteria in the *Parabacteroides* genus (G.Parabacteroides) decreased the odds of lung cancer (odds ratio (OR) with each rank normalized standard deviation (SD) higher relative abundance): 0.84; 95% CI: 0.70, 0.99; P = 0.04) and increased the odds of prostate cancer (OR: 1.17; 95% CI: 1.05, 1.31; P = 0.004) (Figure 2, Supplementary Table 4). The risk of prostate cancer also decreased with higher relative abundances of the unclassified groups of bacteria in the *Firmicutes* phylum (G.unclassified P.Firmicutes; OR: 0.87; 95% CI: 0.77, 0.98; P = 0.02) and in the *Porphyromonadaceae* family (G. Unclassified F. Porphyromonadaceae; OR: 0.91; 95% CI: 0.83, 1.00; P = 0.04) (Figure 2, Supplementary Table 4). The genetic liability to a per doubling increase in abundance of G.Unclassified P.Firmicutes bacteria increased the odds of ovarian cancer (OR 1.054; 95% CI: 1.026, 1.109; P = 0.04) (Figure 2, Supplementary Table 4). There was little evidence for an effect of any of the 14 microbial traits tested on pancreatic cancer (Supplementary Table 4).

**Figure 2.**
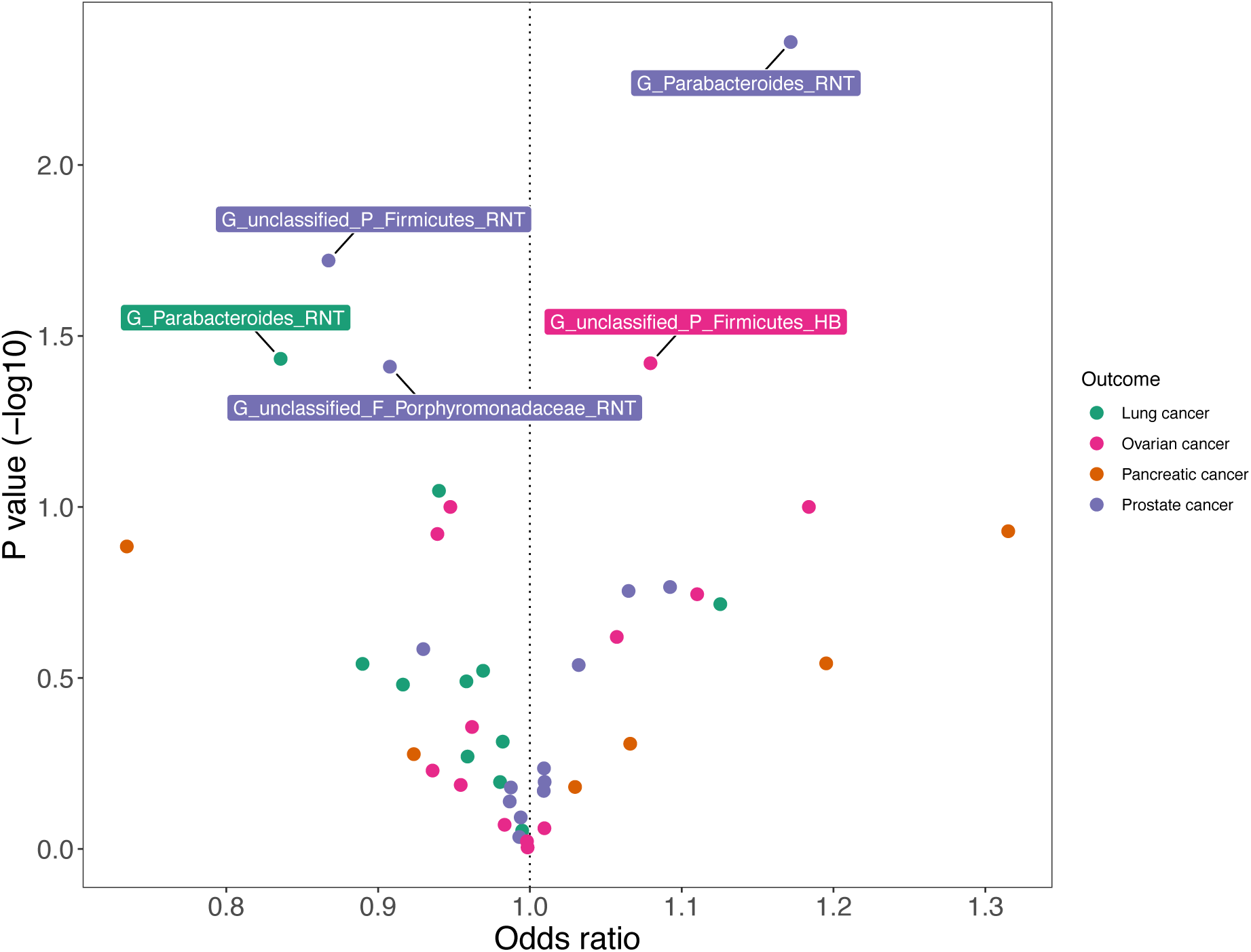
Mendelian randomisation (MR) estimates of the effect of each microbial trait from the Hughes et al. GWAS on the risk of cancers of the lung, ovaries, pancreas and prostate^32^. Abbreviations: RNT=rank normal transformed; HB = hurdle binary; letters in the microbial trait name represent the taxon classification level from which that microbial trait was observed, with “C”, “F”, “G”, “O” and “P” representing “class”, “family”, “genus”, “order” and “phylum”, respectively. All microbial traits that were not confidently classified at the genus level were organised into unclassified groups within higher classification levels (represented by “unclassified”). MR estimates represent the odds ratio (OR) for cancer risk per standard deviation (SD) unit change for continuous microbial traits (labelled as “RNT” in the microbial trait name) or per approximate doubling of the genetic liability to presence (versus absence) of each binary microbial trait (labelled as “HB” in the microbial trait name). Coloured points corresponding to each cancer type, while labels correspond to the microbial traits where there was evidence of a causal effect on a cancer outcome.

Of the 27 microbial traits described in the MiBioGen mGWAS and eight cancers analysed, there was evidence for 18 causal effects across 12 microbial traits on seven cancers. Specifically, the relative abundance of bacteria in the *Intestinibacter* genus (genus.Intestinibacter.id.11345) decreased the risk of lung cancer (OR with each log-transformed SD higher relative abundance: 0.70; 95% CI: 0.51, 0.97; P = 0.03) (Figure 3, Supplementary Table 4). Additionally, a log-transformed SD higher relative abundance of bacteria in the *Streptococcaceae* family (family.Streptococcaceae.id.1850) increased the risks of both lung cancer (OR: 1.47; 95% CI: 1.02, 2.12; P = 0.04) and endometrial cancer (OR: 1.764; 95% CI: 1.10, 2.83; P = 0.02) (Figure 3, Supplementary Table 4). Similarly, with each log-transformed SD higher relative abundance of bacteria in the *Streptococcus* genus (genus.Streptococcus.id.1853) increased the risks of both lung cancer (OR: 1.44; 95% CI: 1.02, 2.04; P = 0.04) and endometrial cancer (OR: 1.71; 95% CI: 1.09, 2.68; P = 0.02) (Figure 3, Supplementary Table 4).

**Figure 3.**
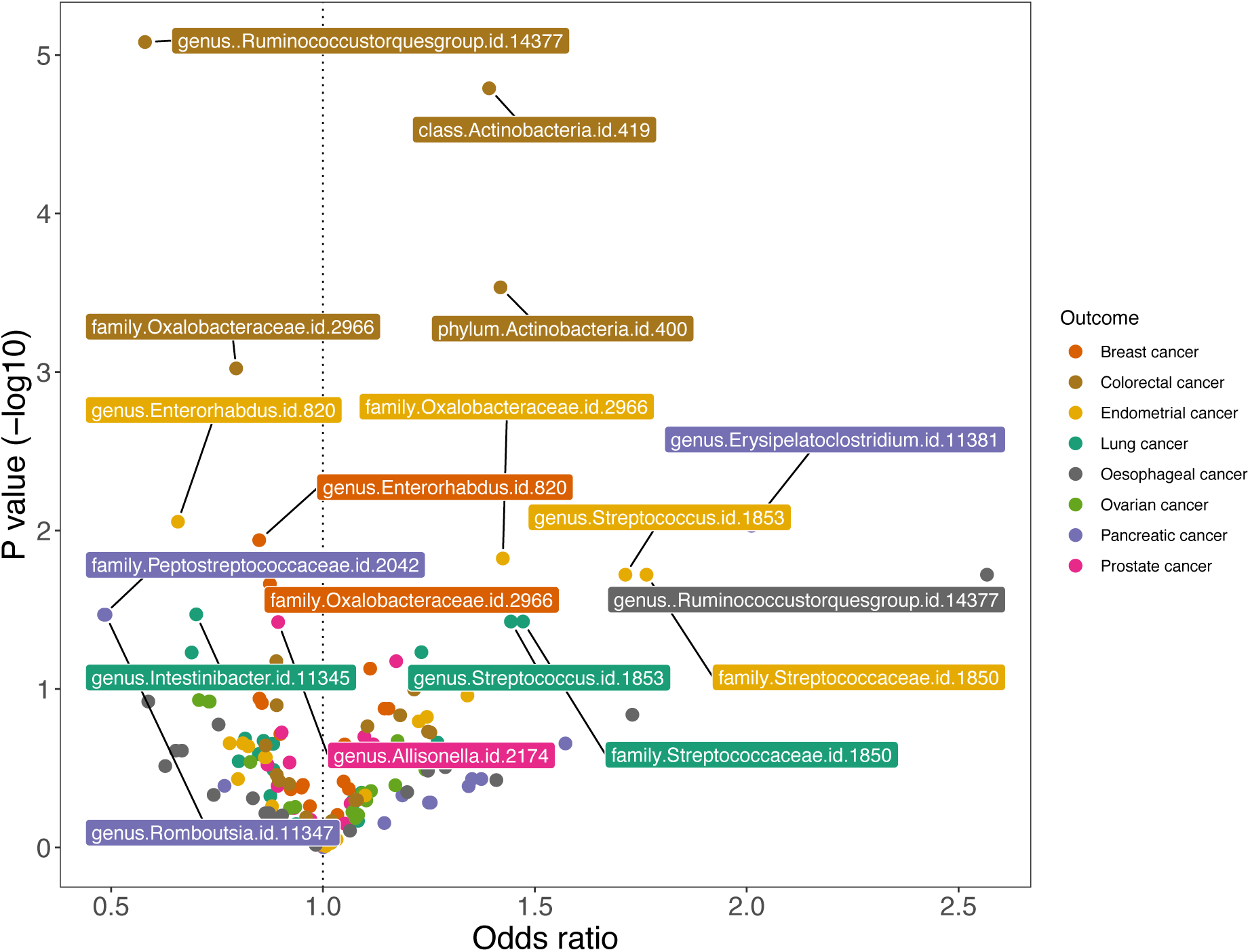
Mendelian randomisation (MR) estimates of the effect of each microbial trait from the MiBioGen consortium on the risk of cancers of the breast, colorectum, endometrium, lung, oesophagus, ovaries, pancreas and prostate^14^. MR estimates represent the odds ratio (OR) for cancer risk per standard deviation (SD) unit change for each continuous microbial trait. Coloured points correspond to each cancer type, while labels correspond to the microbial traits for which there was evidence of a causal effect on a cancer outcome.

There was evidence for a causal role of a higher relative abundance of bacterium within the *Enterorhabdus* genus (genus.Enterorhabdus.id.820) on endometrial cancer (OR: 0.66; 95% CI: 0.48, 0.90; P = 0.009) and breast cancer (OR: 0.85; 95% CI: 0.75, 0.96; P = 0.01) (Figure 3, Supplementary Table 4). Additionally, a higher relative abundance of bacteria in the *Oxalobacteraceae* family (family.Oxalobacteraceae.id.2966) decreased the risk of breast cancer (OR: 0.88; 95% CI: 0.78, 0.98; P = 0.02) and colorectal cancer (OR: 0.80; 95% CI: 0.69, 0.91; P = 0.001), and increased the risk of endometrial cancer (OR: 1.42; 95% CI: 1.07, 1.89; P = 0.02) (Figure 3, Supplementary Table 4).

A higher relative abundance of bacteria in the *Ruminococcus torques group* genus (genus..Ruminococcustorquesgroup.id.14377) increased the risk of oesophageal cancer (OR: 2.57; 95% CI: 1.17, 5.64; P = 0.02) and decreased the risk of colorectal cancer (OR: 0.58; 95% CI: 0.46, 0.74; P = 8.27x10^-06^) (Figure 3, Supplementary Table 4). The risk of colorectal cancer also increased with higher relative abundances of bacteria in the *Actinobacteria* phylum (phylum.Actinobacteria.id.400; OR: 1.42; 95% CI: 1.17, 1.71; P = 2.92x10^-04^) and in the *Actinobacteria* class (class.Actinobacteria.id.419; OR: 1.39; 95% CI: 1.20, 1.62; P = 1.62x10^-05^) (Figure 3, Supplementary Table 4).

There was evidence of a causal role for a higher relative abundance of bacteria in the *Allisonella* genus (genus.Allisonella.id.2174) on the risk of prostate cancer (OR: 0.89; 95% CI: 0.81, 0.99; P = 0.04) (Figure 3, Supplementary Table 4). The risk of pancreatic cancer decreased with a higher relative abundance of bacteria within the *Peptostreptococcaceae* family (family.Peptostreptococcaceae.id.2042; OR: 0.48; 95% CI: 0.25, 0.95; P = 0.03) and the *Romboutsia* genus (genus.Romboutsia.id.11347; OR: 0.49; 95% CI: 0.25, 0.95; P = 0.03) (Figure 3, Supplementary Table 4). In contrast, the risk of pancreatic cancer increased with a higher abundance of bacterium within the *Erysipelatoclostridium* genus (genus.Erysipelatoclostridium.id.11381; OR: 2.01; 95% CI: 1.19, 3.41; P = 0.01) (Figure 3, Supplementary Table 4). There was little evidence for an effect of any of the 27 microbial traits tested on ovarian cancer (Supplementary Table 4).

### Replication

Analyses were repeated for the 23 results showing evidence of a causal effect of gut microbial traits on cancer outcomes using summary statistics from the GWAS of each cancer outcome in both UK Biobank and FinnGen, where appropriate, via the IEU OpenGWAS.^48^ There were five effect estimates that were directionally consistent across the main and replication analyses, where higher relative abundances of bacteria in the (1) G_Parabacteroides decreased the risk of lung cancer, (2) G_Parabacteroides genus increased the risk of prostate cancer, (3) genus.Erysipelatoclostridium.id.11381 increased the risk of pancreatic cancer, (4) genus.Allisonella.id.2174 decreased the risk of prostate cancer and (5) unclassified bacteria in the *Porphyromonadaceae* family decreased the risk of prostate cancer (Supplementary Table 4).

Since UK Biobank was included in the main analysis of colorectal and oesophageal cancer, replication analyses were only undertaken in FinnGen for these outcomes. Here, there was consistency between main and replication analyses for three microbial traits, where higher relative abundances of class.Actinobacteria.id.419 and phylum.Actinobacteria.id.400 increased the risk of colorectal cancer and a higher relative abundance of genus.Ruminococcustorquesgroup.id.14377 increased the risk of oesophageal cancer.

There were several estimates that were directionally consistent in one (but not the other) replication dataset (Supplementary Table 6); however, it is worth noting that confidence intervals for most estimates overlapped the null. Confidence intervals were particularly wide in the FinnGen replication data, where there were the fewest cancer cases. Lastly, replication of results showing an effect of a genetic liability to a per doubling in presence vs. absence of G.unclassified.P.Firmicutes on ovarian cancer was not possible in either FinnGen (due to a lack of GWAS data available for this outcome) or UK Biobank (as the rs34656657 SNP associated with the microbial trait was not present).

### Sensitivity analyses

In addition to the main MR analyses, a series of sensitivity analyses were undertaken where there was evidence for a causal effect of any microbial trait on any cancer outcome to determine whether the identified effects were robust to violations of MR assumptions and likely reflected causality. Specifically, we firstly repeated analyses using the European-only data of the MiBioGen consortium to check for an effect of population stratification given that the MiBioGen consortium comprises individuals of multiple ancestries. We then performed colocalization analyses to check for the presence of a shared causal variant in the genomic region (which is a necessary but not sufficient criterion for causality). We next explored violations of the third MR assumption (Figure 2) by firstly undertaking a manual exploration of horizontal pleiotropy and secondly repeating analyses using a more lenient p-value threshold for instrument selection (whilst restricting to SNPs that were directionally consistent across mGWAS cohorts) to allow for pleiotropy-robust MR analyses. We then performed Steiger tests and, lastly, performed reverse MR analyses, treating each cancer as the exposure and the gut microbial traits as the outcome.

### Analyses with European-only mGWAS data from MiBioGen

Effect estimates when using the summary statistics from the European-only cohorts in the MiBioGen consortium were directionally consistent as the main MR analysis results (Supplementary Table 7). Note that it was not possible to undertake analyses of the effect of genus.Allisonella.id.2174 on prostate cancer, as the European-specific meta-analysis of the associated SNP was in fewer than three cohorts with sample size of <2,000 individuals.

### Colocalisation

Colocalisation analyses were performed to identify whether the SNP associated with the microbial trait was shared with the associated cancer outcome. Across the 23 analyses, 20 of the posterior probabilities for the H1 hypothesis (i.e., that only the gut microbiome has a genetic signal in the region) were greater than H2 (i.e., that only the cancer outcome has a genetic signal in the region) (Supplementary Table 8). Only four of these provided more evidence for colocalization (i.e., the H4 hypothesis) than there being a different causal variant in the genomic region (i.e., the H3 hypothesis). These traits included phylum.Actinobacteria.id.400, class.Actinobacteria.id.419, family.Oxalobacteraceae.id.2966 and genus..Ruminococcustorquesgroup.id.14377 with colorectal cancer (Supplementary Table 8, Supplementary Figures 1-4). However, in all these cases, the posterior probability for H1 was small (i.e., between 0.01-0.38) and many regional association plots suggested that there were multiple outcome-associated SNPs in the genomic region that, in some cases, were in high linkage disequilibrium (LD) with the microbiome-associated SNP (Supplementary Figures 4-8). Regional plots for all other analyses are presented in Supplementary Figures 9-23.

### Manual exploration of pleiotropy

A phenome-wide association analysis was performed to test for associations between each SNP (where a causal effect was observed between the gut microbiome and cancer) on other phenotypes, where associations could indicate the presence of pleiotropy. Indeed, there were many such cases, suggesting that main results may have been driven by horizontal pleiotropy (Supplementary Table 9). For example, the rs4428215 SNP (associated with family.Oxalobacteraceae.id.2966, for which there was evidence for a causal effect on cancers of the breast, colorectum and endometrium in main analyses) was associated with (among other traits) height, trunk fat-free mass, appendicular lean mass, basal metabolic rate, whole body water mass and whole-body fat-free mass (Supplementary Table 9). Given that body mass index is an established risk factor for cancer and likely influences the gut microbiome,^49,50^ the associations observed with adiposity-related phenotypes could be indicative of a bias-inducing pleiotropic pathway of the SNP on these cancers.

### Two-sample MR using a lenient p-value threshold to select genetic instruments

As in previous literature,^23,33,45^ we repeated MR analyses with a more lenient P-value threshold of 1x10^-5^ for selecting instruments. Whilst this allowed for more SNPs to be included in the analyses, doing so likely includes biased or invalid SNPs. To mitigate this possible bias, we additionally restricted to those SNPs that were also directionally consistent across the cohorts contributing to the mGWASs (Supplementary Tables 10-11). Firstly, focusing on the four microbial traits from Hughes et al.^32^ for which there was evidence of an effect on three cancers, no further SNPs were identified for two of the microbial traits (specifically, the relative abundances of G.Parabacteroides and G_unclassified_P_Firmicutes). Therefore, sensitivity analyses were only conducted with the remaining two traits (i.e., presence vs. absence of G_unclassified_P_Firmicutes with ovarian cancer using 11 SNPs and the relative abundance of G_unclassified_F_Porphyromonadaceae and prostate cancer using 3 SNPs). The IVW-derived estimate for the effect of G_unclassified_F_Porphyromonadaceae on prostate cancer was directionally consistent with the main analysis but this was not the case for G_unclassified_P_Firmicutes and ovarian cancer. Additionally, in both cases, estimates were not consistent across all pleiotropy-robust methods (Supplementary Table 10, Supplementary Figures 24-25).

Using the MiBioGen consortium, there were only two SNPs (rs442815 associated with family.Oxalobacteraceae.id.2966 and rs11110281 associated with genus.Streptococcus.id.1853) that were directionally consistent across all European cohorts at a P<1x10^-5^. These two SNPs were the same SNPs that were selected in the main analysis (Supplementary Table 11). Therefore, these results are the same as reported in Supplementary Table 7, for analyses using microbial traits as instruments from the European cohorts from MiBioGen at a genome-wide significance threshold).

Therefore the Wald ratio estimate for the effect family.Oxalobacteraceae.id.2966 on breast cancer (OR: 0.883, 95%CI: 0.795, 0.982, P=0.022), endometrial cancer (OR: 1.388, 1.066, 1.807, P=0.015) and colorectal cancer (OR: 0.809, 95%CI: 0.713, 0.917, P=0.001) and the effect of genus.Streptococcus.id.1853 on lung cancer (OR: 1,484, 95%CI: 1.023, 2.153, P=0.038) and endometrial cancer (OR:1.732, 95%CI: 1.097, 2.736, P=0.019) have already been presented in Supplementary Table 7, and are directionally consistent with the main analysis (Supplementary Table 11).

Due to the limited number of SNPs which were directionally consistent across the European cohorts in MiBioGen, we performed MR sensitivity analyses using all the lenient SNPs associated with microbial traits at P<1x10^-5^ (i.e., not restricting to those that were directionally consistent across European cohorts in MiBioGen). Of the 17 analyses undertaken between microbial traits from MiBioGen and cancer outcomes, 13 IVW-derived estimates were directionally consistent with main MR analyses. However, of these, only five estimates were consistent across all pleiotropy-robust methods (Supplementary Figures 26-30, Supplementary Table 12). Plots for all other analyses of microbial traits on cancer outcomes are presented in Supplementary Figures 31-42. Of these, whilst the confidence intervals for the MR-Egger estimates were expectedly large, there was persistently strong evidence for two microbial traits influence cancer. Specifically, higher relative abundances of genus.Erysipelatoclostridium.id.11381 increased the risk of pancreatic cancer (OR from the IVW analyses using 6 SNPs: 1.36; 95% CI: 1.05, 1.76; P = 0.02) and family.Oxalobacteraceae.id.2966 decreased the risk of breast cancer (OR using 15 SNPs: 0.95; 95% CI: 0.92, 0.99; P = 0.01). Leave-one-out-plots for the latter analyses showed that removing each SNP in turn did not influence the causal effect substantially (Supplementary Figure 26). However, for analyses of genus.Erysipelatoclostridium.id.11381, removing 3 of the 6 associated SNPs individually attenuated the causal effect towards the null, suggesting that the causal effect was not robust (Supplementary Figure 30).

### Steiger test and filtering

Steiger tests were performed to explore whether each SNP being used as an instrument for a microbial trait in the main analysis was explaining more variation in the exposure than the outcome. This would be expected if the causal direction was correct; however, the reverse may indicate that the MR effect estimate is due to reverse causality (i.e., that the SNP influences the outcome, which may in turn influence the exposure). Steiger tests could not be performed where colorectal cancer was the outcome due to some of the SNPs not having an EAF available.

For all main MR analyses across all exposures and outcomes using both the Hughes and MiBioGen data, and sensitivity analyses using the European-only MiBioGen GWAS data and lenient p-value thresholds, the Steiger test results suggested that the assumption that the exposure caused outcome was valid. Therefore, there was little evidence to suggest that the derived effect estimates were driven by reverse causality (Supplementary Tables 13-18).

### Reverse MR analyses

To formally test the potential for reverse causality, MR analyses were undertaken to test the causal effect of the genetic liability to each cancer on the features of the gut microbiome, where a causal effect had been observed in the reverse direction in the main analyses. For most microbial traits where there was evidence of an effect on cancer outcomes in main analyses, there was little evidence for an effect in the reverse direction (Supplementary Table 19, Supplementary Figures 43-65). There was, however, evidence that a higher genetic liability to pancreatic cancer decreased a log-transformed SD in relative abundance of genus.Romboutsia.id.11347 (SD change with the IVW method: - 0.05 SD; 95% CI: -0.09, -0.005; P = 0.03; Supplementary Figure 43) and family.Peptostreptococcaceae.id.2042 (SD change with the IVW method: -0.04; 95% CI: -0.08, -0.001; P = 0.04; Supplementary Figure 44), estimates of which were consistent with pleiotropy-robust methods (Supplementary Table 19). In both instances, the Steiger test indicated that all SNPs used as instruments for pancreatic cancer were in the hypothesised causal direction, suggesting that the effects observed may not be driven by reverse causality. However, leave-one-out plots did indicate that the removal of several SNPs independently did attenuate the causal effect.

## Discussion

In this study, we performed analyses to explore the causal role played by the gut microbiome and cancer. Whilst our initial MR analyses provided evidence that several genera, families and phyla of bacteria had a causal effect on cancers including those of the lung, breast, colorectum, pancreas, oesophagus, ovaries and endometrium, sensitivity analyses highlighted that many of these relationships are unlikely to reflect causality. Specifically, we have shown a range of sensitivity analyses that are imperative to provide context to MR analysis and to explore the underpinning of the apparent associations observed. There were no examples of consistency across the main analysis and all sensitivity analyses which were replicated in UK Biobank and FinnGen datasets. Specifically, whilst we did see some evidence of replication across primary analyses and analyses undertaken in both UK Biobank and FinnGen, results from sensitivity analyses performed did not satisfy evidence of lack of pleiotropy, evidence of a shared causal variant between the exposure trait and the cancer outcome, and consistency between pleiotropy-robust methods. The underlying biology of host genetic effects on the gut microbiome is not well understood, meaning that it is likely that the observed GWAS signals represent host-driven effects upstream of the gut microbiome (i.e., reverse causation) or independent associations between these host genetic variants and cancer (i.e., horizontal pleiotropy). This is further compounded by the lack of SNPs that reach genome-wide significance within mGWASs and that replicate across mGWASs, despite the F-statistics suggesting that these would be appropriate instruments for an MR analysis.

Notwithstanding the lack of clear evidence for a causal effect of the gut microbiome on cancer, our findings were informative for several important reasons. Firstly, results of analyses including cohorts of multiple ancestries from the MiBioGen consortium were consistent with results obtained when restricting to European-only cohorts. This suggests that the effects estimates observed did not arise from statistical artefact due to differences in genetic architecture between ancestral populations, likely due to the MiBioGen consortium comprising largely European populations (72%). However, many existing studies have used MiBioGen data without acknowledging or considering that it is fundamentally a consortium including individuals of mixed ancestry, which may lead to confounding attributable to population structure. Here, whilst we have demonstrated that this may not be a considerable bias, the enects of such bias may be context specific. Therefore, we recommend that future studies use homogeneous cohorts (e.g., those of similar ancestry) to minimise the effects of this potential population structure or at least as a sensitivity test to analyses using heterogeneous cohorts.

Secondly, we manually identified pleiotropy for every microbial trait implicated in the main MR analyses, except for rs11098863 (associated with genus.Enterorhabdus.id.820) and rs11110281 (associated with both genus.Streptococcus.id.1853 and family.Streptococcaceae.id.1850), where there were no phenotypes associated with this SNP using a P<1x10^-5^. Whilst it is not possible to tell if this indicates the presence of horizontal or vertical pleiotropy from these analyses alone, it does show that pleiotropy is very likely with SNPs associated with the gut microbiome. For example, the rs4428215 SNP was associated with a number of other traits with relevance to adiposity, including trunk mass, arm mass, weight and triglycerides. It could be that the relationships between rs4428215, family.Oxalobacteraceae.id.2966 and these adiposity-related traits reflect the causal pathway by which the gut microbiome influences breast cancer. However, it is equally plausible that rs4428215 independently influences both this gut microbial trait and adiposity, where the latter only influences breast cancer. Therefore, the MR results presented here should be treated with caution. Further work could investigate these pleiotropic pathways (for example, using multivariable MR to disentangle the relationship between adiposity-related traits, the gut microbiome on cancer); however, such analyses are challenging currently given the lack and complexity of SNPs associated with the gut microbiome compared to other host traits such as adiposity.

By relaxing our p-value threshold for instrument selection, we were able to apply pleiotropy-robust methods to formally test the validity of the third (exclusion restriction) core MR assumption. Importantly, however, for some traits we were unable to restrict instruments to those that had directional consistency across contributing mGWAS cohorts. In fact, only two MiBioGen SNPs were directionally consistent across all the contributing mGWAS cohorts of European ancestry, which highlights the level of heterogeneity across mGWAS results. These differences likely reflect population structure driven by differences in age, diet, body mass index, or medication^14^, but may also arise due to sampling methods, ^51^ especially given that seven methods of faecal extraction and three 16S rRNA regions were used to sequence the microbial species in the MiBioGen study samples.^14^ Whilst this is only an issue if such differences are correlated with both genetic variation and the gut microbiome, this still presents a major limitation when performing MR using these mGWASs.

When restricting analyses to use instruments that had the same direction of effect across the cohorts, the results for almost all traits were consistent with the main analysis. However, there was little consistency across pleiotropy-robust methods, indicating evidence of pleiotropy. Similarly, there was directional inconsistency across pleiotropy-robust methods and IVW estimates when using all SNPs associated at a P<1x10^-5^ threshold. Combined with the manual exploration of pleiotropy (as described above), this suggests that the MR analyses are very likely biased due to pleiotropic SNPs and questions whether the effects observed in the main analyses were indeed causal. This underscores the importance of using SNPs that are reliably and consistently associated with gut microbial traits in future MR analyses exploring the impact of the microbiome on health outcomes, and appropriately testing the validity of MR assumptions when making causal inferences.

When testing for reverse causality, there was very little evidence to suggest that a genetic liability to cancer influenced variation in microbial traits and steiger testing in the main analysis suggested that reverse causality was not likely. However, there was evidence that a higher genetic liability to pancreatic cancer decreased the relative abundance of genus.Romboutsia.id.11347 and family.Peptostreptococcaceae.id.2042. This suggests reverse causality could be a potential source of bias and should be cautioned in future MR studies of the microbiome.

Comparison with prior MR studies of the microbiome and cancer highlights how analytic differences can drive discrepant conclusions, with several published studies used lenient instrument thresholds (P < 1×10⁻⁵) and limited sensitivity frameworks increasing susceptibility to weak instrument bias and the inclusion of invalid genetic variants.^52–54^ In some cases adjusted for “confounders” including body mass index and smoking related traits, in ways that are conceptually inconsistent with MR assumptions and may induce collider bias.^52^ Indeed, as we have shown here, results obtained when using a lenient p-value threshold, even when restricting to SNPs that had the same direction of effect across discovery GWAS analyses, were mostly inconsistent with those obtained when using the strongest and currently most robust instruments for each microbial trait (i.e., those meeting P<5x10^-8^). Further to this, while other studies have not included colocalisation^53,54^or explored potential pleiotropy^53,54^ in the analysis framework, our results show that, even when primary associations are observed, formal interrogation of colocalisation, pleiotropy and reverse causality frequently weakens causal claims.

Long et al. reported evidence for 11 putative causal associations between microbial taxa and cancer outcomes including breast, colorectal, ovarian, head and neck, lung, endometrial and prostate cancer, using genetic instruments selected at a conventional genome-wide significance threshold (P <5 × 10⁻⁸).^54^ While some findings were concordant with those observed here—for example, associations involving bacteria within the *Ruminococcus torques* group and colorectal cancer—there were also notable discrepancies across other taxa and cancer sites. These discrepancies are likely attributable to substantive differences in analytical pipelines between the two studies. In contrast to the approach used here, Long et al. did not systematically assess colocalization between microbiome and cancer signals, limiting the ability to distinguish shared causal variants from distinct but correlated genetic effects.^54^ In addition, pleiotropy was evaluated using a restricted set of Mendelian randomization methods without comprehensive phenome-wide exploration of instrument validity, and no formal tests of reverse causality—such as Steiger filtering or bidirectional Mendelian randomization—were undertaken. Furthermore, analyses were not repeated using ancestry-restricted microbiome GWAS data to assess potential bias arising from population structure.

A key strength of this present study is the use of the strongest, most robust microbiome-related SNPs as instruments.^14,32^ Whilst we additionally used a more lenient p-value threshold, we also restricted this set of SNPs to those that had the same direction of effect across cohorts contributing to the discovery GWASs, where possible. This was also, importantly, a sensitivity analysis to complement and be explicitly compared to the main analysis results, under the assumption that, in a larger GWAS, these SNPs would have reached a traditional genome-wide p-value threshold. Another strength of this study is the replication performed in UK Biobank and FinnGen. Here, we showed that there were few examples of consistency across all three sources. This may be attributed to a smaller number of cancer cases present in UK Biobank and FinnGen datasets, meaning these GWAS may have been underpowered to detect real causal effects, different population structures across these studies that may lead to genetic confounding (i.e., a violation of the second MR assumption), or that the initial, non-replicated results may have arisen due to chance. Lastly, the implementation of a comprehensive set of sensitivity analyses exploring the likelihood of a shared causal variant, pleiotropy and reverse causality is a key strength in addressing the assumptions of MR in this study, which should be a requirement for all future MR studies of the gut microbiome to make appropriate causal inferences.

A key limitation of the present study, as with other Mendelian randomization analyses of the gut microbiome such as Hatcher and colleagues’ investigation of microbial traits in colorectal cancer,^23^ is the current limited understanding of host genetic influences on microbial variation and the relatively small sample sizes of existing microbiome GWASs. Hatcher et al. used summary statistics from a meta-analysis of ∼3,890 individuals to instrument 14 microbial traits and found that initial MR estimates suggesting causal effects were largely attenuated or inconsistent once sensitivity analyses, including exploration of pleiotropy and reverse causation, were applied, highlighting the complexity of available instruments and the difficulty of deriving robust causal inferences.^23^ Similarly, in our study the number of robust, independent genetic instruments available for many microbial traits remains limited, resulting in low variance explained and reduced power to detect causal effects, particularly for less abundant taxa. In both studies, genetic instruments may also be subject to horizontal pleiotropy, as variants influencing microbial traits often have multiple associations with other traits that could independently affect cancer risk.^23^ This is compounded by the challenge of colocalisation analysis, where even when colocalisation is formally tested, available GWAS data for some microbial traits may be insufficient to provide definitive evidence of shared causal variants. Additionally, both works are constrained by the use of 16S rRNA–based taxonomic profiling, which lacks species- or strain-level resolution and functional information, and by potential heterogeneity in microbiome measurement protocols across cohorts, which can introduce noise and bias in instrument discovery.^23^ Microbial traits from the MiBioGen consortium were analysed as log-transformed relative abundances that were residualised for covariates prior to genome-wide association analysis and combined across cohorts using z-score–based meta-analysis.^14^ As a result, genetic associations are not scaled to the same units as the Hughes et al,^32^ mGWAS, where traits reflect rank-transformed normalised SD units. Accordingly, Mendelian randomisation estimates for comparing MiBioGen traits and Hughes et al. traits should be interpreted by direction and relative magnitude rather than a precisely defined quantitative change.

Finally, as in Hatcher et al.,^23^ reverse causality cannot be fully excluded for all traits despite bidirectional and Steiger filtering tests, particularly given the modest numbers of instruments for many exposures. Together, these shared limitations underscore the need for larger, more harmonised microbiome GWASs with improved taxonomic resolution and for continued methodological development to strengthen causal inference in host–microbiome research. Progress in microbiome MR will depend on microbiome GWAS designs that improve instrument strength and validity: larger, harmonised cohorts; standardised sampling, sequencing and bioinformatic pipelines; and parallel application of high-resolution microbiome profiling.^32^ Shotgun metagenomic sequencing, could enable species- or strain-level instruments and improve biological interpretability of SNP–microbe associations, supporting more specific MR hypotheses. Interdisciplinary efforts integrating host genetics, microbial ecology and functional studies will be essential to determine whether host variants exert direct effects on microbial traits versus indirect or pleiotropic influences, thereby strengthening confidence in instrument validity.

To summarise, in this work, we aimed to appropriately apply MR to understand the role of the gut microbiome in cancer aetiology, whilst demonstrating the range of sensitivity analyses required to provide clarity in causal estimates. Whilst main MR findings initially suggested that there were a handful of bacterial genera, families and phyla that may alter cancer risk, further sensitivity analyses indicated that these results were unlikely to reflect causality. Through testing for violations of core MR assumptions, our work further demonstrates the importance of performing such sensitivity analyses when assessing the robustness of MR findings, especially in the context of understanding the causal relevance of the gut microbiome on health outcomes given the complexity of host-microbiome interactions.

## METHODS

### Study design

Two-sample MR was used to examine the causal relationship between features of the human gut microbiome and overall cancer risk for cancers of the lung, breast, colorectum, pancreas, prostate, ovary, endometrium and oesophagus. This study and all methods have been conducted in line with the Strengthening the Reporting of Observational Studies in Epidemiology MR (STROBE-MR) reporting guidelines for MR studies.^55^

### Gut microbiome GWAS data and instrument selection

Genetic variants associated with microbial traits were obtained from two of the largest microbiome GWASs (mGWASs).^14,32^ The first was a meta-analyses of bacterial abundance, presence (versus absence), alpha- and beta-diversity and enterotype conducted within the Flemish Gut Flora Project (FGFP, n = 2,223) and two independent validation cohorts – the Food-Chain Plus study (FoCus, n = 950) and the PopGen study (n = 717).^32^ Full details of the data sampling, preparation and analyses have been described previously.^23,32^ Information on the microbial traits analysed and SNPs selected as genetic instruments from this mGWAS are in the Supplementary Methods and Supplementary Table 1. Briefly, a total of 14 SNPs from this mGWAS were selected as genetic instruments for the 14 microbial traits for two-sample MR analyses with cancer outcomes. Effect estimates from the mGWAS represent an increase in the log-transformed odds ratio (OR) for a per doubling in the genetic liability to presence vs. absence of binary microbial traits, and the standard deviation (SD) change for log-transformed beta effect estimates that were derived from Pearsons Correlation, for the relative abundance of continuous microbial traits with each effect allele carried. The methodology of deriviation of units of relative abundance have previously been described in detail by Kurilshikov et al.

The second dataset was the mGWAS meta-analysis of bacterial abundance, presence (versus absence) and alpha-diversity in 18,340 participants from 24 cohorts in the MiBioGen consortium. Details on data sampling, preparation and mGWAS analyses have been described previously.^14^ Information on the microbial traits analysed and SNPs selected as genetic instruments for gut microbial traits from the MiBioGen mGWAS are in the Supplementary Methods and Supplementary Table 2. Briefly, a total of 21 unique SNPs from this mGWAS associated with 27 unique microbial traits were selected as genetic instruments for MR analyses with cancer outcomes. Effect estimates reflect an SD change for log transformed relative abundance of continuous microbial traits with each effect allele carried.

### Cancer GWAS data

For all cancer outcomes, summary-level data from the largest and most recent GWASs were obtained. Full details on data sources, cancer definitions and SNPs used as instruments for each cancer are presented in the Supplementary Methods and Supplementary Table 4.^56–70^ Effect estimates obtained from GWAS summary statistics reflected the log-transformed OR for the risk of each cancer.

### Replication

Where possible, as a replication, two-sample MR analyses were repeated using SNP-outcome associations obtained from GWAS analyses of cancer outcomes in both UK Biobank^71,72^ and FinnGen,^73^ where samples were independent from those used in the main analyses. All data used for replication analyses were available in the IEU OpenGWAS database.^48^ Full information on the data sources, cancer outcomes and SNPs used in analyses is provided in the Supplementary Methods and Supplementary Table 4.

### Statistical analyses

#### Two-sample Mendelian randomization

In our primary analyses, we performed two-sample MR using the *TwoSampleMR* package^74^ (version 0.6.8) in R (version 4.4.1) to examine the causal relationship between 41 microbial traits (i.e., 14 from the Hughes mGWAS and 27 from the MiBioGen mGWAS)^14,32^ and the risk of eight cancers. Summary-level data (i.e., the SNP name [rsid], effect allele, other allele, effect allele frequency, beta coefficient, standard error, p-value and sample size) for each of the SNPs associated with the 41 microbial traits across both mGWASs were extracted from the respective mGWAS data and each cancer GWAS. The exposure and outcome datasets were harmonized such that the effect of each SNP on the exposure and outcome was relative to the same effect allele. For ambiguously coded SNPs (i.e., “palindromic” SNPs where the effect/other allele were either an A/T or G/C combination), we used the effect allele frequency (EAF) where available to resolve strand ambiguity, where possible. Non-inferable SNPs (i.e., “palindromic” SNPs with a MAF > 0.42) were removed from the analysis. If there was no allele frequency available, we undertook analyses assuming that the alleles were all on the forward strand, after comparing the exposure and outcome data sources. Proxy SNPs were used if the exposure-related SNP was not available in the cancer outcome GWAS. Both the variance explained in each microbial trait by each SNP (R^2^) and an indicator of instrument strength (F-statistic) were calculated. Full details for obtaining proxy SNPs, R^2^, F-statistics and power calculations are presented in the Supplementary Materials. The number of SNPs removed at each stage of the analysis pipeline (i.e., availability in outcome data and harmonization) are provided in Supplementary Table 5.

Given that, for the majority of instances, there was only one SNP associated with each microbial trait, the Wald ratio method was used as the main analyses, which estimates the effect of the exposure (each microbial trait) on the outcome (each cancer outcome) by dividing the SNP-outcome association by the SNP-exposure association.^26,27^ Where there was more than one SNP associated with the microbial trait, the inverse-variance weighted (IVW) random effects method was used, which combines Wald ratios together in an IVW meta-analysis, weighted by the inverse variance of the SNP-outcome association, adjusting for heterogeneity. Cochran’s Q test for heterogeneity was used in this calculation to formally estimate the heterogeneity across individual SNPs.^74,75^

### Sensitivity analyses

Assessing the evidence of a causal relationship between the gut microbiome and cancer risk, there are three MR assumptions that must be met: (1) the relevance assumption (the genetic instruments used to proxy the exposure are strongly associated with that exposure), (2) the independence assumption (there is no confounding between the genetic instrument(s) used to proxy the exposure and the outcome) and (3) the exclusion restriction assumption (the genetic instruments used to proxy the exposure have no effect on the outcome other than through the exposure) – see Figure 1.^23,26,27^ With regards to the first assumption, as described above (and in more detail within the Supplementary Materials), we used the strongest SNPs associated with each microbial trait across both mGWASs (i.e., those meeting study- and genome-wide p-value thresholds) and estimated the F-statistic for each instrument to check validity of the first MR assumption. To ensure homogeneity across exposure and outcome datasets and thus lowering the likelihood of violations of the second MR assumption, we used independent SNPs associated with our exposures of interest and data that had adjusted for genetic principal components and were of European ancestry.

Importantly, microbial traits for which there was evidence of a causal effect on any cancer outcome were then followed up with a series of analyses that assessed the robustness of our findings to violations of MR assumptions. Full details on the sensitivity analyses undertaken are presented in the Supplementary Methods.

Briefly, as the MiBioGen mGWAS comprised individuals of multiple ancestries (∼72% European), we firstly repeated analyses using summary statistics obtained only from the European cohorts and compared these results to the main analyses. Secondly, genetic colocalisation was used to evaluate whether there was a shared causal variant between the microbial trait and cancer outcome, which is a necessary but not sufficient criterion for causality. Thirdly, we performed a phenome-wide analysis for SNPs associated with each microbial trait that showed a causal effect on a cancer outcome to test for possibly pleiotropy.^76^ Fourth, to formally test for directional pleiotropy, we selected genetic variants to proxy microbial traits at a more lenient p-value threshold of 1x10^−05^ (restricting to those that had directionally consistent effect estimates across GWAS cohorts, where possible) and then repeated MR analyses with several complementary, pleiotropy-robust methods including the MR-Egger, weighted median and weighted mode approaches.^77–79^ Fifth, to assess whether any causal effect observed between microbial traits and cancer risk in our main analyses was indicative of reverse causation (i.e., where the SNP influences the cancer outcome which, in turn, influences the microbial trait), the Steiger test was performed to identify genetic variants where there was evidence that the microbiome-related SNP explained more variation in the cancer outcome than the gut microbial trait, which may indicate that the observed effect is not in this hypothesised causal direction. Lastly, we performed formal two-sample MR analyses in the reverse direction, with cancer as the exposure and microbial traits as the outcome.

Effect estimates obtained from all two-sample MR analyses represent the OR for cancer risk for each rank normalized SD higher abundance of each continuous microbial trait (MiBioGen microbial traits and the Hughes traits denoted with “RNT”) and the OR for cancer risk for an approximate doubling of the genetic liability to presence (versus absence) of each binary microbial trait (those denoted with “HB”). As a key aim of this paper was to demonstrate the sensitivity analyses required to test the robustness of effects observed between the gut microbiome and cancer outcomes, coupled with the high correlation between microbial traits, we did not correct for multiple testing. Instead, conclusions were drawn based on effect sizes and their precision, where P-values were interpreted as continuous indicators of evidence strength.

## Supporting information

STROBE-MR checklist

## Data Availability

All data produced in the present study are available upon reasonable request to the authors.
Example code for one cancer outcome will be made available upon publication to a public github repository, detailing the R packages and analyses performed.

https://github.com/amydawes/gut-microbiome-and-cancer-MR-and-sensitivity-analyses-

## Declarations

### Data availability

GWAS summary-level data used in this study were publicly available for microbiome GWAS (mGWAS) conducted by Hughes et al. and Kurilshikov et al.^14,32^ Summary-level data for overall cancer were obtained from the IEU Open GWAS database where available^48^, or else is described in the Supplementary Methods, and data sources listed in Supplementary Table 4.

### Code availability

Example code for one cancer outcome will be made available upon publication to a public github repository (https://github.com/amydawes/gut-microbiome-and-cancer-MR-and-sensitivity-analyses-), detailing the R packages and analyses performed.

### Contributions

A.D. designed the study, produced code, completed the analysis and drafted/revised the manuscript. A.M. performed the European-specific MiBioGen meta-analysis. C.H. produced code which was adapted to run the analyses in this study. K.W. designed the study, provided support throughout the studyimplementation and drafting of manuscript. A.M., C.H, and K.W. provided feedback on manuscript prior to submission.

### Ethics declarations

Ethical approval was not required for this study.

## Acknowledgements

A.D. is supported by Cancer Research UK [grant number RCCPDF\100007, awarded to K.W.]. A.M. is supported by Cancer Research UK [grant number C18281/A30905]. K.W. is supported by the University of Bristol. The funders had no influence on study design, data collection and analysis, decision to publish or preparation of the manuscript.

## References

1. Macmillan Cancer Support. Causes and risk factors, <https://www.macmillan.org.uk/cancer-information-and-support/worried-about-cancer/causes-and-risk-factors> (

2. Bull, M. J. & Plummer, N. T. Part 1: The Human Gut Microbiome in Health and Disease. Integr Med (Encinitas) 13, 17–22 (2014).

3. Hanahan, D. & Weinberg, R. A. Hallmarks of cancer: the next generation. Cell 144, 646–674, doi:10.1016/j.cell.2011.02.013 (2011).

4. Visconti, A. et al. Interplay between the human gut microbiome and host metabolism. Nat Commun 10, 4505, doi:10.1038/s41467-019-12476-z (2019).

5. Blander, J. M., Longman, R. S., Iliev, I. D., Sonnenberg, G. F. & Artis, D. Regulation of inflammation by microbiota interactions with the host. Nat Immunol 18, 851–860, doi:10.1038/ni.3780 (2017).

6. Francescone, R., Hou, V. & Grivennikov, S. I. Microbiome, inflammation, and cancer. Cancer J 20, 181–189, doi:10.1097/PPO.0000000000000048 (2014).

7. Zheng, D., Liwinski, T. & Elinav, E. Interaction between microbiota and immunity in health and disease. Cell Res 30, 492–506, doi:10.1038/s41422-020-0332-7 (2020).

8. Manos, J. The human microbiome in disease and pathology. APMIS 130, 690–705, doi:10.1111/apm.13225 (2022).

9. Rinninella, E. et al. What is the Healthy Gut Microbiota Composition? A Changing Ecosystem across Age, Environment, Diet, and Diseases. Microorganisms 7, doi:10.3390/microorganisms7010014 (2019).

10. Plichta, D. R., Graham, D. B., Subramanian, S. & Xavier, R. J. Therapeutic Opportunities in Inflammatory Bowel Disease: Mechanistic Dissection of Host-Microbiome Relationships. Cell 178, 1041–1056, doi:10.1016/j.cell.2019.07.045 (2019).

11. Frank, D. N. et al. Molecular-phylogenetic characterization of microbial community imbalances in human inflammatory bowel diseases. Proc Natl Acad Sci U S A 104, 13780–13785, doi:10.1073/pnas.0706625104 (2007).

12. Garrett, W. S. Cancer and the microbiota. Science 348, 80–86, doi:10.1126/science.aaa4972 (2015).

13. Purushothaman, S., Meola, M. & Egli, A. Combination of Whole Genome Sequencing and Metagenomics for Microbiological Diagnostics. International Journal of Molecular Sciences 23, 9834 (2022).

14. Kurilshikov, A. et al. Large-scale association analyses identify host factors influencing human gut microbiome composition. Nature Genetics 53, 156–165, doi:10.1038/s41588-020-00763-1 (2021).

15. Urbaniak, C. et al. The Microbiota of Breast Tissue and Its Association with Breast Cancer. Appl Environ Microbiol 82, 5039–5048, doi:10.1128/AEM.01235-16 (2016).

16. Hieken, T. J. et al. The Microbiome of Aseptically Collected Human Breast Tissue in Benign and Malignant Disease. Sci Rep 6, 30751, doi:10.1038/srep30751 (2016).

17. Ambalam, P., Raman, M., Purama, R. K. & Doble, M. Probiotics, prebiotics and colorectal cancer prevention. Best Pract Res Clin Gastroenterol 30, 119–131, doi:10.1016/j.bpg.2016.02.009 (2016).

18. Gopalakrishnan, V., Helmink, B. A., Spencer, C. N., Reuben, A. & Wargo, J. A. The Influence of the Gut Microbiome on Cancer, Immunity, and Cancer Immunotherapy. Cancer Cell 33, 570–580, doi:10.1016/j.ccell.2018.03.015 (2018).

19. Ishikawa, H. et al. Randomized trial of dietary fiber and Lactobacillus casei administration for prevention of colorectal tumors. International Journal of Cancer 116, 762–767, 10.1002/ijc.21115 (2005).

20. Wade, K. H. & Hall, L. J. Improving causality in microbiome research: can human genetic epidemiology help? Wellcome Open Res 4, 199, doi:10.12688/wellcomeopenres.15628.3 (2019).

21. Zhang, Z. J. et al. Assessment of Causal Direction Between Gut Microbiota and Inflammatory Bowel Disease: A Mendelian Randomization Analysis. Front Genet 12, 631061, doi:10.3389/fgene.2021.631061 (2021).

22. Ahn, J. et al. Human gut microbiome and risk for colorectal cancer. J Natl Cancer Inst 105, 1907–1911, doi:10.1093/jnci/djt300 (2013).

23. Hatcher, C. et al. Application of Mendelian randomization to explore the causal role of the human gut microbiome in colorectal cancer. Scientific Reports 13, 5968, doi:10.1038/s41598-023-31840-0 (2023).

24. Rezasoltani, S. et al. The association between fecal microbiota and different types of colorectal polyp as precursors of colorectal cancer. Microb Pathog 124, 244–249, doi:10.1016/j.micpath.2018.08.035 (2018).

25. Dikeocha, I. J., Al-Kabsi, A. M., Eid, E. E. M., Hussin, S. & Alshawsh, AM. A. Probiotics supplementation in patients with colorectal cancer: a systematic review of randomized controlled trials. Nutr Rev 80, 22–49, doi:10.1093/nutrit/nuab006 (2021).

26. Davey Smith, G. & Hemani, G. Mendelian randomization: genetic anchors for causal inference in epidemiological studies. Hum Mol Genet 23, R89–98, doi:10.1093/hmg/ddu328 (2014).

27. Haycock, P. C. et al. Best (but oft-forgotten) practices: the design, analysis, and interpretation of Mendelian randomization studies. Am J Clin Nutr 103, 965–978, doi:10.3945/ajcn.115.118216 (2016).

28. Richmond, R. C. & Davey Smith, G. Mendelian Randomization: Concepts and Scope. Cold Spring Harb Perspect Med 12, doi:10.1101/cshperspect.a040501 (2022).

29. Wade, K. H. et al. Applying Mendelian randomization to appraise causality in relationships between nutrition and cancer. Cancer Causes Control 33, 631–652, doi:10.1007/s10552-022-01562-1 (2022).

30. Yang, Q., Lin, S. L., Kwok, M. K., Leung, G. M. & Schooling, C. M. The Roles of 27 Genera of Human Gut Microbiota in Ischemic Heart Disease, Type 2 Diabetes Mellitus, and Their Risk Factors: A Mendelian Randomization Study. Am J Epidemiol 187, 1916–1922, doi:10.1093/aje/kwy096 (2018).

31. Sanna, S. et al. Causal relationships among the gut microbiome, short-chain fatty acids and metabolic diseases. Nat Genet 51, 600–605, doi:10.1038/s41588-019-0350-x (2019).

32. Hughes, D. A. et al. Genome-wide associations of human gut microbiome variation and implications for causal inference analyses. Nat Microbiol 5, 1079–1087, doi:10.1038/s41564-020-0743-8 (2020).

33. Fryer, E., Hatcher, C., Knight, R. & Wade, K. Exploring the causal role of the human gut microbiome in endometrial cancer: a Mendelian randomization approach. medRxiv, 2024.2003.2006.24303765, doi:10.1101/2024.03.06.24303765 (2024).

34. Jia, J. et al. Assessment of Causal Direction Between Gut Microbiota-Dependent Metabolites and Cardiometabolic Health: A Bidirectional Mendelian Randomization Analysis. Diabetes 68, 1747–1755, doi:10.2337/db19-0153 (2019).

35. Zhao, Y. et al. Role of lung and gut microbiota on lung cancer pathogenesis. J Cancer Res Clin Oncol 147, 2177–2186, doi:10.1007/s00432-021-03644-0 (2021).

36. Zheng, Y. et al. Specific gut microbiome signature predicts the early-stage lung cancer. Gut Microbes 11, 1030–1042, doi:10.1080/19490976.2020.1737487 (2020).

37. Javier-DesLoges, J., et al. The microbiome and prostate cancer. Prostate Cancer Prostatic Dis 25, 159–164, doi:10.1038/s41391-021-00413-5 (2022).

38. Fujita, K., et al. Gut microbiome and prostate cancer. Int J Urol 29, 793–798, doi:10.1111/iju.14894 (2022).

39. Dhingra, A., Sharma, D., Kumar, A., Singh, S. & Kumar, P. Microbiome and Development of Ovarian Cancer. Endocr Metab Immune Disord Drug Targets 22, 1073–1090, doi:10.2174/1871530322666220509034847 (2022).

40. Riquelme, E. et al. Tumor Microbiome Diversity and Composition Influence Pancreatic Cancer Outcomes. Cell 178, 795–806.e712, doi:10.1016/j.cell.2019.07.008 (2019).

41. Attebury, H. & Daley, D. The Gut Microbiome and Pancreatic Cancer Development and Treatment. Cancer J 29, 49–56, doi:10.1097/PPO.0000000000000647 (2023).

42. Wei, M.-Y. et al. The microbiota and microbiome in pancreatic cancer: more influential than expected. Molecular Cancer 18, 97, doi:10.1186/s12943-019-1008-0 (2019).

43. Baba, Y. et al. Relationship between gut microbiome Fusobacterium nucleatum and LINE-1 methylation level in esophageal cancer. Esophagus 20, 704–712, doi:10.1007/s10388-023-01009-9 (2023).

44. Chen, X. et al. Oral Microbiota and Risk for Esophageal Squamous Cell Carcinoma in a High-Risk Area of China. PLoS One 10, e0143603, doi:10.1371/journal.pone.0143603 (2015).

45. Chataway, J., Hamilton, G., Hatcher, C. & Wade, K. H. Application of bidirectional Mendelian randomization to assess the relationship between the gut microbiome and esophageal cancer. medRxiv, 2025.2002.2006.25321825, doi:10.1101/2025.02.06.25321825 (2025).

46. Edmunds, G. L. et al. Exploring the causal role of the human gut microbiome in breast cancer risk. medRxiv, 2025.2002.2012.25322118, doi:10.1101/2025.02.12.25322118 (2025).

47. Fryer, E., Hatcher, C., Knight, R. & Wade, K. H. Exploring the causal role of the human gut microbiome in endometrial cancer: a Mendelian randomization approach. Scientific Reports 15, 11953, doi:10.1038/s41598-025-96740-x (2025).

48. Elsworth, B. et al. The MRC IEU OpenGWAS data infrastructure. bioRxiv, 2020.2008.2010.244293, doi:10.1101/2020.08.10.244293 (2020).

49. CancerResearchUK. How does obesity cause cancer?, <https://www.cancerresearchuk.org/about-cancer/causes-of-cancer/bodyweight-and-cancer/how-does-obesity-cause-cancer#:~:text=A%20BMI%20of%2025%20or%20higher%20increases%20the%20risk%20of,take%20to%20reduce%20your%20risk.> (

50. Hughes, D. A. et al. Estimating the causal effect of body mass index on gut microbiota variation. medRxiv, 2025.2008.2018.25333926, doi:10.1101/2025.08.18.25333926 (2025).

51. Sinha, R. et al. Assessment of variation in microbial community amplicon sequencing by the Microbiome Quality Control (MBQC) project consortium. Nat Biotechnol 35, 1077–1086, doi:10.1038/nbt.3981 (2017).

52. Ni, J. J. et al. Mendelian randomization study of causal link from gut microbiota to colorectal cancer. BMC Cancer 22, 1371, doi:10.1186/s12885-022-10483-w (2022).

53. Su, Q. et al. Association between gut microbiota and gastrointestinal cancer: a two-sample bi-directional Mendelian randomization study. Front Microbiol 14, 1181328, doi:10.3389/fmicb.2023.1181328 (2023).

54. Long, Y., Tang, L., Zhou, Y., Zhao, S. & Zhu, H. Causal relationship between gut microbiota and cancers: a two-sample Mendelian randomisation study. BMC Medicine 21, 66, doi:10.1186/s12916-023-02761-6 (2023).

55. Skrivankova, V. W. et al. Strengthening the Reporting of Observational Studies in Epidemiology Using Mendelian Randomization: The STROBE-MR Statement. Jama 326, 1614–1621, doi:10.1001/jama.2021.18236 (2021).

56. Fernandez-Rozadilla, C. et al. Deciphering colorectal cancer genetics through multi-omic analysis of 100,204 cases and 154,587 controls of European and east Asian ancestries.

57. Zhang, H. et al. Genome-wide association study identifies 32 novel breast cancer susceptibility loci from overall and subtype-specific analyses. Nat Genet 52, 572–581, doi:10.1038/s41588-020-0609-2 (2020).

58. O’Mara, T. A. et al. Identification of nine new susceptibility loci for endometrial cancer.

59. Wang, Y. et al. Rare variants of large effect in BRCA2 and CHEK2 affect risk of lung cancer. Nature Genetics 46, 736–741, doi:10.1038/ng.3002 (2014).

60. Schumacher, F. R. et al. Association analyses of more than 140,000 men identify 63 new prostate cancer susceptibility loci.

61. Klein, A. P. et al. Genome-wide meta-analysis identifies five new susceptibility loci for pancreatic cancer.

62. Amundadottir, L. et al. Genome-wide association study identifies variants in the ABO locus associated with susceptibility to pancreatic cancer. Nat Genet 41, 986–990, doi:10.1038/ng.429 (2009).

63. Petersen, G. M. et al. A genome-wide association study identifies pancreatic cancer susceptibility loci on chromosomes 13q22.1, 1q32.1 and 5p15.33. Nat Genet 42, 224–228, doi:10.1038/ng.522 (2010).

64. Wolpin, B. M. et al. Genome-wide association study identifies multiple susceptibility loci for pancreatic cancer. Nat Genet 46, 994–1000, doi:10.1038/ng.3052 (2014).

65. Childs, E. J. et al. Common variation at 2p13.3, 3q29, 7p13 and 17q25.1 associated with susceptibility to pancreatic cancer. Nat Genet 47, 911–916, doi:10.1038/ng.3341 (2015).

66. Wang, Z. et al. Imputation and subset-based association analysis across different cancer types identifies multiple independent risk loci in the TERT-CLPTM1L region on chromosome 5p15.33. Hum Mol Genet 23, 6616–6633, doi:10.1093/hmg/ddu363 (2014).

67. Schröder, J. et al. GWAS meta-analysis of 16 790 patients with Barrett’s oesophagus and oesophageal adenocarcinoma identifies 16 novel genetic risk loci and provides insights into disease aetiology beyond the single marker level. Gut 72, 612–623, doi:10.1136/gutjnl-2021-326698 (2023).

68. Levine, D. M. et al. A genome-wide association study identifies new susceptibility loci for esophageal adenocarcinoma and Barrett’s esophagus. Nat Genet 45, 1487–1493, doi:10.1038/ng.2796 (2013).

69. Song, H. et al. A genome-wide association study identifies a new ovarian cancer susceptibility locus on 9p22.2. Nat Genet 41, 996–1000, doi:10.1038/ng.424 (2009).

70. Phelan, C. M. et al. Identification of 12 new susceptibility loci for different histotypes of epithelial ovarian cancer. Nature Genetics 49, 680–691, doi:10.1038/ng.3826 (2017).

71. Burrows, K. & Haycock, P. Genome-wide Association Study of Cancer Risk in UK Biobank., <https://data.bris.ac.uk/data/dataset/aed0u12w0ede20olb0m77p4b9> (2021).

72. Bycroft, C. et al. The UK Biobank resource with deep phenotyping and genomic data. Nature 562, 203–209, doi:10.1038/s41586-018-0579-z (2018).

73. Kurki, M. I. et al. FinnGen provides genetic insights from a well-phenotyped isolated population. Nature 613, 508–518, doi:10.1038/s41586-022-05473-8 (2023).

74. Hemani, G. et al. The MR-Base platform supports systematic causal inference across the human phenome. Elife 7, doi:10.7554/eLife.34408 (2018).

75. Hemani, G., Tilling, K. & Davey Smith, G. Orienting the causal relationship between imprecisely measured traits using GWAS summary data. PLOS Genetics 13, e1007081, doi:10.1371/journal.pgen.1007081 (2017).

76. Hemani, G., Elsworth, B., Palmer, T. & Rasteiro, R. (2024).

77. Bowden, J., Davey Smith, G. & Burgess, S. Mendelian randomization with invalid instruments: effect estimation and bias detection through Egger regression. Int J Epidemiol 44, 512–525, doi:10.1093/ije/dyv080 (2015).

78. Bowden, J., Davey Smith, G., Haycock, P. C. & Burgess, S. Consistent Estimation in Mendelian Randomization with Some Invalid Instruments Using a Weighted Median Estimator. Genet Epidemiol 40, 304–314, doi:10.1002/gepi.21965 (2016).

79. Hartwig, F. P., Davey Smith, G. & Bowden, J. Robust inference in summary data Mendelian randomization via the zero modal pleiotropy assumption. Int J Epidemiol 46, 1985–1998, doi:10.1093/ije/dyx102 (2017).

