## Supplementary methods, Supplementary Figures 1-63 for "Application of Mendelian randomization to appraise causality in relationships between the gut microbiome and cancer"

**Data**

The following sections summarise the genome-wide association studies (GWASs) used for each gut microbiome exposure and cancer outcome in this Mendelian randomization (MR) analysis and sensitivity analyses.

*Hughes et al. gut microbiome GWAS (mGWAS)*

Hughes et al. performed a GWAS of bacterial abundance, presence (versus absence), alpha- and beta-diversity and enterotype conducted within the Flemish Gut Flora Project (FGFP, n = 2,223) and two independent validation cohorts (the Food-Chain Plus study (FoCus, n = 950) and the PopGen study (n = 717).(1) In the FGFP cohort, of the 92 identified taxa, 62 contained substantial zero-inflation.(1) These taxa were modelled using a two-step hurdle binary analysis, which includes a binary presence (versus absence) analysis (denoted throughout as "HB") and a zero-truncated rank normal transformed abundance analysis (denoted throughout as "RNT"). All other taxa were relatively normally distributed; therefore, were treated as abundance phenotypes and rank normal transformed accordingly. Of the 153 microbial traits tested in the mGWAS, two SNPs showed evidence of association that exceeded the study-wide meta-analysis p-value threshold ($1.57x10^{-10}$) and a further 11 exceeded the genome-wide meta-analysis threshold ($2.5x10^{-08}$), where each SNP was associated with one microbial trait. These 13 SNPs were selected as genetic instruments for the 13 microbial traits within this two-sample MR analysis with cancer outcomes, in addition to one SNP associated with bacteria of the *Bifidobacterium* genus that has previously been reported in the literature.(1-4) Effect estimates from the mGWAS represent an increase in the log-transformed odds ratio (OR) for a per doubling in the genetic liability to presence vs. absence of binary microbial traits and the standard deviation (SD) change for rank normalised relative abundance of continuous microbial traits with each effect allele carried. Full summary data are available here: https://data.bris.ac.uk/data/dataset/22bqn399f9i432q56gt3wfhzlc. In this work, full summary statistics were only available on the FGFP cohort, so sensitivity analyses requiring full summary statistics across the genome were performed using FGFP data rather than the meta-analysis across the three cohorts.

*MiBioGen mGWAS*

This dataset includes twenty cohorts of single ancestry, namely European (16 cohorts; *n* = 13,266), Middle Eastern (1 cohort; *n* = 481), East Asian (1 cohort; *n* = 811), American Hispanic/Latin (1 cohort; *n* = 1,097) and African American (1 cohort; *n* = 114), whereas four cohorts included samples from multiple ancestries (*n* = 2,571). In this work, we performed our main analyses using the full summary statistics across multiple ancestries (n = 18,340). In sensitivity analyses (see below), we also used the mGWAS results in individuals of European ancestry (n = 13,266). In total in the primary analysis, 21 unique SNPs were associated with 27 unique traits at a genome-wide significant P-value threshold of $5x10^{-8}$. Effect estimates reflect an SD change for log transformed relative abundance of continuous microbial traits with each effect allele carried. Log transformed relative abundance SD units were derived from a pearson correlation cooefficient analysis, from which Z-scores were generated. Further detail of the units of microbial traits are provided by Kurilshikov et al.(5)

*Cancer GWASs*

Data for colorectal cancer (CRC) were obtained from the most comprehensive GWAS of overall CRC to date from a meta-analysis of 185,616 individuals of European ancestry (comprising 78,473 CRC cases and 107,143 controls).(6) The GWAS meta-analysis included the Genetics and Epidemiology of Colorectal Cancer Consortium (GECCO), Colorectal Cancer Transdisciplinary Study (CORECT), OncoArray + Custom, OmniExpress + Exome Chip, COloRectal cancer Study of Austria (CORSA) and UK Biobank. Full information on genotyping, imputation and quality control (QC) was not provided. CRC case status was diagnosed by a physician.

Summary-level breast cancer GWAS data were obtained from a GWAS of overall breast cancer from the most recent Breast Cancer Association Consortium (BCAC) meta-analysis.(7) Women were included if they had a diagnosis of a primary invasive breast cancer without known metastases. Overall breast cancer data from BCAC comprised a meta-analysis of several GWASs including the Collaborative Oncological Gene-environment Study (iCOGs) (38,349 cases and 37,818 controls), Oncoarray (80,125 cases and 58,385 controls) and others (17,588 cases and 14,910 controls), resulting in a total meta-analysis sample size of 136,062 cases and 111,113 controls. Participants in BCAC were enrolled from 1970 until 2020 with all individuals being of European ancestry. Participants were enrolled from 28 countries (Australia, Austria, Belarus, Belgium, Canada, Czech Republic, Denmark, Finland, Germany, Greece,

Hungary, Ireland, Israel, Italy, Latvia, Lithuania, Macedonia, Netherlands, New Zealand, Norway, Poland, Portugal, Russia, South Africa, Spain, Sweden, UK, USA). Full information on genotyping, imputation and QC have been described previously (7). Briefly, genotyped or imputed variants (including bi-allelic and multi-allelic single-nucleotide polymorphisms (SNPs) and small indels) were determined using the iCOGS and OncoArray genotyping arrays and imputed to the 1000 Genomes Project (phase 3) reference panel. Variants were included with an imputation quality score of >0.3. Variants included in the analysis were restricted to a minor allele frequency of >0.005.

Data for endometrial cancer was downloaded from the IEU Open GWAS (GWAS ID: ebi-a-GCST006464-6). Data were obtained from a GWAS meta-analysis of 17 studies of participants of European ancestry, totalling 12,906 endometrial cancer cases and 108,979 country-matched controls.(8) Studies used in this GWAS meta-analysis included the Epidemiology of Endometrial Cancer Consortium (E2C2), the Endometrial Cancer Association Consortium (ECAC) and UK Biobank. These analyses also specified the different histological subtypes of endometrial cancer, endometrioid (cases=8,758; controls=46,126) and non-endometrioid (cases=1,230; controls=35,447). The remaining cases were either of mixed or unknown histology. If a study did not have any cases of non-endometrioid cancer, it was not included in the meta-analysis and, thus, the number of controls were lower than for endometrioid cancer. Further details on the GWAS meta-analysis are provided elsewhere.(8, 9) Data from the E2C2 genome-wide association studies (GWAS) have been previously described.(10, 11) There are a total of 7,077 endometrial cancer cases and 16,343 controls from 15 studies (ten case–control and five cohort, which were analysed as nested case–control). The Oncoarray genotyping chip was used to genotype 533,631 variants and used to genotype 5061 endometrial cancer cases from ten studies in Australia, Belgium, Germany, Sweden, UK, and USA.(12) SNPs with a call rate <95%, SNPs not in Hardy–Weinberg equilibrium (HWE) ($P < 10^{-7}$ in controls and $P < 10^{-12}$ in cases) and SNPs with concordance <98% among 5280 duplicate pairs of samples were excluded, leaving 483,972 SNPs. Prior to imputation, SNPs with MAF <1% and call rate <98% in any consortium were also excluded, in addition to SNPs that could not be linked to the 1000 Genomes Project reference panel or for which the MAF differed significantly from the European reference panel frequency. A further 1128 SNPs were excluded after review of cluster plots, leaving 469,364 SNPs used in the imputation. Additional exclusions were made for duplicates and close relatives, totalling 4710 cases and 19,438 controls. 2695 cases and 2777 controls from the E2C2 consortium were genotyped on the Illumina Human OmniExpress array or the Illumina Human 660W array and were imputed to 1000 genomes project V3 reference panel using standard QC procedures(13). 288 cases from six population based case-control studies within the Women's Health Initiative were genotyped using five different arrays and separately imputed using combined 1000 Genome Project V3 and UK10K reference panels, following standard QC procedures and SNPS excluded with a MAF<1%.(13) UK Biobank samples were genotyped using the Affymetrix UK BiLEVE Axiom array and Affymetrix UK Biobank Axiom® array and imputed to 1000 genome project V3 and UK10K reference panels.(14) Following SNP-wise QC, the analysis included a total of 12,906 endometrial cancer cases and 108,979 controls, which included 8,758 endometrioid cases and 1,230 non-endometrioid cases. The remaining cases were mixed histology or there was no data on histology available.

Summary-level lung cancer GWAS data were accessed through the Integrative Epidemiology Unit (IEU) Open GWAS database (GWAS ID: ieu-a-987). Data were obtained from the Transdisciplinary Research Into Cancer of the Lung (TRICL) GWAS meta-analysis of six previously reported lung cancer GWAS of European populations: the MD Anderson Cancer Center lung cancer study (MDACC-GWAS) (1,150 cases and 1,134 controls); the UK lung cancer GWAS from the Institute for Cancer Research (ICR-GWAS) (1,952 cases); the NCI lung cancer GWAS (NCI-GWAS)(2,100 cases and 2,120 controls) the IARC lung cancer GWAS (IARC-GWAS)(3,062 lung cancer cases and 4,455 controls) the Lung Cancer in the Young (LUCY) and Kora Studies from Germany and a hospital based case-control study from the Lunenfeld-Tanenbaum Research Institute of Mount Sinai Hospital and University of Toronto, of which there are 847 lung cancer patients in the LUCY study, >2000 lung cancer cases in the Heidelberg lung cancer case-control study (the number of cases in the KORA study was not specified) and 331 cases and 499 controls in the Toronto study.(15, 16) In each of the studies, SNP genotyping had been performed using Illumina HumanHap 317, 317+240S, 370, 550, 610 or 1M arrays. Details of imputation were not provided. Standard QC was performed excluding individuals with a lower call rate <90%, and extremes in high or low heterozygosity ($P<1.0x10^{-4}$), as well as individuals of non-European ancestry. This analysis used data from 29,863 overall lung cancer cases and 55,586 controls. This data is unpublished but details of the individual studies included in the meta-analysis are available online.(16) All cases had received a diagnosis of pathologically confirmed lung cancer. Tumors from patients were classified as adenocarcinomas (AD), squamous carcinomas (SQ), large-cell carcinomas (LCC), mixed adenosquamous carcinomas (MADSQ) and other non-small cell lung cancer (NSCLC) histologies following either the International Classification of Diseases for Oncology (ICD-O) or World Health Organization (WHO) coding. Tumours were classified as mixed where there were overlapping histologies.(16)

Summary level prostate cancer GWAS data were accessed through the IEU Open GWAS database using IEU Open GWAS ID: ebi-a-GCST006085, which was a meta-analysis of the Prostate Cancer Association Group to Investigate Cancer Associated Alterations in the Genome (PRACTICAL) consortium and the Elucidating Loci Involved in Prostate Cancer Susceptibility (ELLIPSE) consortium, which included a custom high-density OncoArray of 46,939 prostate cancer cases and 27,910 controls of European ancestry with previously genotyped data of 32,255 prostate cases and 33,202 controls of European ancestry from the United States and Europe.(17) In total, 79,148 overall prostate cancer cases and 61,106 controls were included in this analysis. For most included studies, prostate cancer was diagnosed in clinic, however some studies identified cases on cancer registry Information, including the European Prospective Investigation into Cancer and Nutrition (EPIC) where cases were identified through self-report on follow-up questionnaires and verified through medical records or cancer registries. Detail on SNP selection, genotyping, imputation and sample processing is described elsewhere.(17) Briefly, 50% of the Oncoarray was a list of SNPs reported by the Genetic Asssociations and Mechanisms in Oncology (GAME-ON) consortium of cancer, while the remaining content was selected as a Illumina HumanCore 'GWAS backbone' to provide high coverage for most common variants through imputation. 79,000 SNPs were selected for their relevance to prostate cancer. 533,631 variants were included on the array. SNPs were excluded with a call rate <98% by study, where not in HWE ($P < 10^{-7}$ in controls or $P < 10^{-12}$ in cases) or with concordance <98% among 11,260 duplicates. SNPs were excluded with a MAF<1%, where not linked to the reference from the 1000 Genomes project or where the MAF in Europeans differed from the 1000 genomes project reference. A further 16,526 SNPs were excluded based on the cluster plot. This left a panel of 498,417 SNPs among samples of European ancestry after QC. Duplicate samples and first-degree relatives within each study were excluded, along with samples with a call rate <95% and samples with extreme heterozygosity (>4.9 standard deviation from the mean reported ancestry). Genotypes for ~70 million SNPs were imputed for all samples across OncoArray and GWAS datasets using the 1000 Genomes project V3 reference panel.

We used data from the latest, and largest pancreatic cancer GWAS meta-analysis in this analysis, as previously described by Klein et al (18). This included 9,055 pancreatic cancer cases and 7,203 controls. Data from 16 cohorts and 13 case–control studies genotyped in four previous GWAS phases, were meta-analysed: The Pancreatic Cancer Cohort Consortium (PanScan) I, PanScan II, PanScan III, and the Pancreatic Cancer Case-Control Consortium (PanC4) (18-22). PanScan and PanC4 genome-wide association data were obtained through dbGAP (accession numbers phs000206.v5.p3 and phs000648.v1.p1, respectively). Blood or buccal cells were collected prior to pancreatic cancer diagnosis for each cohort that participated in PanScan.(19) Incident primary pancreatic adenocarcinoma cases were identified by self-report with subsequent medical record review, and/or linkage with a cancer registry. Cases were defined as primary adenocarcinoma of the exocrine pancreas (ICD-O-3 code C250–C259). Non-exocrine pancreatic tumors (histology type 8150, 8151, 8153, 8155 and 8240) were excluded. Genotyping for PanScan was performed at the Cancer Genomics Research Laboratory (CGR) of the National Cancer Institute (NCI) of the National Institutes of Health (NIH) using the Illumina HumanHap series arrays (Illumina HumanHap550 Infinium II, Human 610-Quad) for PanScan I and II, and the Illumina Omni series arrays (OmniExpress, Omni1M, Omni2.5, and Omni5M) for PanScan III (19-21).

Genotyping for the PanC4 GWAS was performed at the Johns Hopkins Center for Inherited Disease Research (CIDR) using the Illumina HumanOmniExpressExome-8v1 array. Imputation was performed using the 1000 Genomes project Phase 3, Release 1 reference data set and IMPUTE2.(21, 23-25) There was a large overlap of variants on genotyping arrays for PanScan I and II, so these data sets were imputed and analysed together, while PanScan III and PanC4 GWAS data sets were each imputed and analysed separately.

Oesophageal cancer GWAS data were obtained from the largest oesophageal cancer performed by Schröder et al. (26). This includes 5,582 cases of oesophageal adenocarcinoma (EA) and 32,476 controls. For all patients, the diagnosis was provided by cancer registry data of either an ICD-10 code of C155 (C15.5 Malignant neoplasm of lower third of oesophagus) or C152 (C15.2 Abdominal part of oesophagus) and was confirmed by histopathology. GWAS cohorts included the Barrett's and Esophageal Adenocarcinoma Consortium (BEACON, from United States, Canada, Australia, Sweden, and Cambridge and Oxford UK), Bonn V1 (Germany), Bonn V2 (Germany), 2,789 blood donors of the Institute of Transfusion Medicine, University Hospital of Schleswig-Holstein, Kiel, Germany, and 240 EA cases plus 12,534 controls from the UK Biobank (26, 27). Genotyping, quality control and imputation has been described previously (26, 27). Briefly, samples from the previous GWAS in the BEACON cohort were genotyped on Illumina HumanOmni1-Quad array.(28) Samples from the previous GWAS Cambridge cohort were genotyped on Illumina HumanOmni1-Quad and Illumina custom Human 1.2M-Duo arrays. Samples from the previous GWAS Oxford cohort were genotyped on the Illumina 660Q-Quad, Illumina custom Human 1.2M-Duo array, Illumina Hap1M and Illumina Hap550 arrays. Samples from the previous GWAS BonnV1 were genotyped on the HumanCoreExome, PsychArray and HumanOmniExpress arrays. Samples from the new GWAS cohort BonnV2 cohort were genotyped on the Illumina Global Screening Array (V2) and samples from the UK Biobank cohort were genotyped on the Affymetrix UK BiLEVE Axiom array, and the Affymetrix UK Biobank Axiom array. Samples and SNPs with missingness >3% and HWE $P<1x10^{-6}$ were excluded. For Bonn V2 samples with phenotypic and genotypic sex mismatch samples were removed. GWAS data was imputed against two panels (Haplotype Reference Consortium(29) and merged 1000 Genomes Project (phase 3) plus UK10K (30, 31) to match the imputation strategy used for the UK Biobank samples which were imputed by UK Biobank(14). The dataset for BEACON is available in dbGAP and the dataset of the Cambridge cohort is available via EDAM.(26) The other datasets analysed during this study by Schröder et al. are not publicly available due to non-conformity with consent forms but were made available from the corresponding author on reasonable request.

Ovarian cancer GWAS data were accessed by the IEU Open GWAS database using IEU Open GWAS ID: ieu-a-1120. This includes 25,509 cases of ovarian cancer and 40,941 controls from the Ovarian Cancer Association Consortium (OCAC), previously published .(32, 33) Individuals were of European ancestry. Information on cancer diagnosis was not provided in the study. Cases were defined by histotype as follows: serous borderline (1,954), mucinous borderline (1,149), low grade serous ovarian cancer (LGSOC) (1,012), high grade serous ovarian cancer (HGSOC) (13,037), endometrioid ovarian cancer (ENOC) (2,810), clear cell odontogenic carcinoma () (1,366) and other epithelial ovarian cancer (EOC) (2,764).Genotyping data from six OCAC genotyping projects were used and genotyped on a custom Illumina Oncoarray. Genotyping QC has been previously described.(33, 34) Samples were excluded if they had a genotyping call rate <95%, if they had excessively low or high heterozygosity, if they were not female or if they were duplicates (cryptic or intended). Samples with concordance >0.86 were flagged as duplicates, and samples with concordance between 0.74 and 0.86 were flagged as relatives. The comparison was performed among all the OncoArray samples and among all the previously genotyped samples. Data from the 1000 genomes project reference panel was used to impute genotypes for 11,403,952 common variants (MAF>1%).

*Replication datasets*

GWAS data from UK Biobank and FinnGen (accessed via the IEU Open GWAS) were used as replication datasets, where available. UK Biobank cancer cases were recorded according to the International Classification of Diseases (ICD9, ICD10) with data completed to April 2019.(35) Cases were diagnosed by the type of cancer and behaviour of cancer tumour as recorded in national cancer registries. UK Biobank samples were genotyped using the Affymetrix UK BiLEVE Axiom array and Affymetrix UK Biobank Axiom® array and imputed to Haplotypes Reference Consortium (HRC) and UK10K reference panels.(14) Cancer GWAS were performed in individuals of European ancestry, and restrictions were made to include a maximal set of unrelated individuals based on kinship coefficients provided by UK Biobank.(35, 36) Further details on genotyping, QC and GWAS analysis performed by UK Biobank are provided elsewhere.(36) Following initial QC, principal components analysis (PCA) was performed and samples were excluded if missing rate on autosomes > 0.02, were not in a set of unrelated individuals and there was a sex-mismatch between inferred sex and self-reported sex. SNPs were excluded where there was a missing rate >0.015, MAF<0.01 and in regions of long-range linkage disequilibrium (LD). FinnGen cancer cases were defined by ICD-10 codes in the care register for health care, and in causes of deaths and the Finnish Cancer Registry records. Controls were defined as not having any other cancer incidence recorded. In the FinnGen study, 154,714 individuals were genotyped using a custom Axiom FinnGen1 array, while 70,023 additional individuals were derived from legacy collections genotyped with non-custom genotyping arrays. Further details on genotyping and QC is available elsewhere.(37) The genotyping array consists of 736,145 probes interrogating 655,973 variants. In addition to the core imputation 'backbone', 116,402 rare (MAF < 0.5%) coding variants (97,665 missense and 20,432 predicted loss of function) that had been identified in 15 000 Finnish exomes were directly genotyped. Along with variants for the KIR and HLA haplotypes (10,800), known pathogenic ClinVar variants (14,900), pharmacogenomic markers (4,600) and 57,008 additional markers that were of special interest for the FinnGen partners. QC criteria included sample-wise missingness rate >0.05%, heterozygosity rate greater than population average ±4 standard deviations (SD), sample contamination detection with PI-HAT >0.1 to more than 14 samples, PCA outliers in the first 2 dimensions ±4SD. Sex-check F-value ≤0.3 was used for females and ≥0.8 for expected male samples. Variant-wise QC was done by applying Hardy-Weinberg equilibrium test p-value cut-off of $<10^{-6}$, call-rate of >98% and minimum allele count of 1. This retained 224 737 samples.

All primary MR analyses were repeated using summary statistics from the GWAS of each cancer outcome in UK Biobank and/or FinnGen, where available via the IEU Open GWAS.(38) For some cancers, it was not possible to perform such replication, due to an unavailability of published GWAS summary statistics in UK Biobank or FinnGen, or where primary MR analysis included UK Biobank or FinnGen data. This was the case for CRC, where the CRC GWAS meta-analysis used in the main MR analysis contained UK

Biobank, therefore replication in UK Biobank was not appropriate. Additionally, the primary analysis for endometrial cancer contains UK Biobank cases in the meta-analyses, therefore replication in UK Biobank could not be performed. There was no available endometrial cancer GWAS replication dataset in FinnGen. Similarly, there was no available ovarian cancer GWAS in FinnGen hence this replication could not be performed. The primary analysis for oesophageal cancer contains UK Biobank samples and therefore the replication in UK Biobank could not be performed.

Replication using a UK Biobank lung cancer GWAS (IEU Open GWAS ID ieu-b-4954) provided 2,671 overall lung cancer cases and 372,016 controls.(35) (35)Lung cancer GWAS data obtained from FinnGen was accessed using IEU Open GWAS ID: finn-b-C3_BRONCHUS_LUNG_EXALLC, which provided a replication dataset 1,681 cases of malignant neoplasm of bronchus and lung and 173,993 controls.

A breast cancer GWAS including 13,879 breast cancer cases and 198,523 controls from UK Biobank was accessed using IEU Open GWAS ID: ieu-b-4810 and used in replication analyses. Breast cancer GWAS data from FinnGen was accessed using the IEU Open GWAS ID: finn-b-C3_BREAST_EXALLC and provided 8,401 cases and 99,321 controls in the replication analysis. Colorectal cancer GWAS data from FinnGen was accessed using IEU Open GWAS ID: finn-b-C3_COLORECTAL_EXALLC and included 3022 cases and 174,006 controls used in the replication analysis. Pancreatic cancer GWAS data from FinnGen was accessed using IEU Open GWAS ID: finn-b-C3_PANCREAS_EXALLC and included 605 cases and 174,006 controls used in the replication analysis. Pancreatic cancer GWAS data from UK Biobank was accessed using IEU Open GWAS ID: ebi-a-GCST90018893 and included 1196 cases and 475049 controls which were used in replication. Prostate cancer GWAS data from UK Biobank was accessed using IEU Open GWAS ID: ieu-b-4809 and included 9132 cases and 173493 male controls which were used in replication. Prostate cancer GWAS data from FinnGen was accessed using IEU Open GWAS ID: finn-b-C3_PROSTATE_EXALLC and consists of 6311 cases and 74685 controls which were used in replication analysis. Oesophageal cancer GWAS data from FinnGen was accessed using IEU Open GWAS ID: finn-b-C3_OESOPHAGUS_EXALLC and consists of 232 cases and 174006 controls which were used in replication analysis. Ovarian cancer GWAS data from UK Biobank was accessed using IEU Open GWAS ID: ieu-b-4963 and consists of 1218 cases and 198523 controls used in replication analysis. Further details are provided in Supplementary Table 4.

*Reverse two-sample Mendelian randomization datasets*

As sensitivity analysis, MR analyses were performed in the reverse direction where there was evidence of a causal effect in the main MR analysis, where each cancer was the exposure and the microbial traits were the outcomes. For most cancers, the same cancer GWAS was used as the exposure in the reverse MR analysis that was previously used as the outcome in the main MR analysis. However, there were some exceptions for breast cancer, colorectal cancer and pancreatic cancer where the largest and most recent publically available GWAS data was used, where the data in the main MR analysis is not publically available.

Summary-level breast cancer GWAS data from BCAC were obtained from the IEU Open GWAS database using ID: ieu-a-1126. This consisted of 122977 European breast cancer cases and 105974 European controls from BCAC.(39) Women were included if they had a diagnosis of a primary invasive breast cancer without known metastases. The

GWAS genotyped 61,282 female cases from 68 studies in BCAC and Discovery, Biology and Risk of Inherited Variants in Breast Cancer Consortium (DRIVE) using the Oncoarray, and genotypes for 21 million variants were imputed using 1000 genomes project phase 3 reference panel. Filtering on MAF<0.5% and an imputation quality score <0.3 was performed in addition to adjusting for principal components. Results were meta-analysed (using a fixed effect) with 46,785 cases and 42,892 controls from iCOGS and 11 other breast cancer GWAS (14,910 cases and 17,588 controls).

Colorectal cancer GWAS data were accessed by the IEU Open GWAS database using ID: ebi-a-GCST012879. This consisted of 19948 cases and 12124 controls used in the reverse MR analysis. Huyghe et al. performed whole genome sequencing in 1,439 CRC cases and 720 controls of European ancestry at low sequencing depth (3.8–8.6×) and then imputed variants using HRC into 34,860 cases and 29,051 controls of European (91.7%) and East Asian ancestry (8.3%) from 30 existing GWAS forming the stage 1 meta-analysis and was used to inform the design of a custom Illumina Oncoarray.(40) 12,007 cases and 12,000 controls of European ancestry were genotyped on the custom array and combined with an additional 11,255 cases and 26296 controls resulting in 23,262 cases and 38,296 controls (stage 2 meta-analysis). A combined stage 1 and stage 2 meta-analysis was performed totalling 58,131 cases and 67,347 controls. It is unclear why the dataset available in the IEU Open GWAS database is only a subset of this total meta-analysis. No information was provided by Huyghe et al. on how CRC was diagnosed.(40)

Pancreatic cancer GWAS summary data consisting of the genome-wide significantly associated SNPs and effect estimates were downloaded from the supplementary material by Klein et al.(18) In this pancreatic cancer GWAS data there were 9040 cases of pancreatic cancer and 12,496 controls. GWAS data was similarly obtained from the PanScan I, PanScan II, PanScan III and PanC4 studies and the genotyping, QC and imputation process is the same as described above for the data used in the primary analysis. Incident primary pancreatic adenocarcinoma cases were identified by self-report with subsequent medical record review, and/or linkage with a cancer registry. Cases were defined as primary adenocarcinoma of the exocrine pancreas (ICD-O-3 code C250–C259). Non-exocrine pancreatic tumours (histology type 8150, 8151, 8153, 8155 and 8240) were excluded.

### **Statistical analyses**

*Two-sample Mendelian randomization*

We used the strongest SNPs associated with each microbial trait across both mGWASs (i.e., those meeting study- and genome-wide p-value thresholds) and estimated the F-statistic for each instrument to check validity of the first MR assumption. Summary-level data (i.e., the SNP name [rsid], effect allele, other allele, effect allele frequency, beta coefficient, standard error, p-value and sample size) for each of the SNPs associated with the 41 microbial traits across both mGWASs were extracted from the respective mGWAS data and each cancer GWAS. Where the SNP was not available in the outcome cancer GWAS, we searched for a proxy SNP using LDlink using the LDproxy_batch() function in R, using the following parameters: pop = "CEU", r2d = "r2", append = TRUE, genome_build = "grch37", api_root = https://ldlink.nih.gov/LDlinkRest”. We selected proxies with the largest $r^2$ values and smallest distance away from the original SNP. SNP rsids were extracted from biomaRt and then we searched for the proxy SNP in the original microbiome GWAS and the outcome data. In some cases, we were unable to find an appropriate proxy SNP that was present in the original mGWAS and the outcome cancer GWAS, see Supplementary Table 5.

Given that, for the majority of instances, there was only one SNP associated with each microbial trait, the Wald ratio method was used as the main analyses, which estimates the effect of the exposure (each microbial trait) on the outcome (each cancer outcome) by dividing the SNP-outcome association by the SNP-exposure association.(41, 42) Where there was more than one SNP associated with the microbial trait, the inverse-variance weighted (IVW) random effects method was used, which combines Wald ratios together in an IVW meta-analysis, weighted by the inverse variance of the SNP-outcome association, adjusting for heterogeneity. Cochran's Q test for heterogeneity was used in this calculation to formally estimate the heterogeneity across individual SNPs.(43, 44)

The exposure and outcome datasets were harmonized such that the effect of each SNP on the exposure and outcome was relative to the same effect allele. For ambiguously coded SNPs (i.e., "palindromic" SNPs where the effect/other allele were either an A/T or G/C combination), we used the effect allele frequency (EAF) where available to resolve strand ambiguity, where possible. Non-inferable SNPs (i.e., "palindromic" SNPs with a MAF > 0.42) were removed from the analysis. If there was no allele frequency available, we undertook analyses assuming that the alleles were all on the forward strand, after comparing the exposure and outcome data sources.

The proportion of variance explained ($R^2$) in each microbial trait by each SNP and the strength of the instrument (assessed through the F-statistic) were calculated, see Supplementary Methods. For each binary microbial trait, the proportion of variance explained of the liability to presence (versus absence) was obtained using the "*get_r_from_lor*" function in the *TwoSampleMR* package using the beta (i.e., log odds ratio), EAF, number of cases, number of controls and the prevalence of the exposure as input. The required parameter for this function describing prevalence of each microbial trait was obtained from FGFP alone (as published by Hughes et al.(35)), as prevalence estimates of each microbial trait were not available in the two other cohorts. For continuous microbial traits (i.e., abundances) the $R^2$ was calculated using the following formula using the "*steiger_filtering*" function in the *TwoSampleMR* package:

$$R2 = \frac{2\beta^2(eaf)(1 - eaf)}{(2\beta^2 eaf(1 - eaf)) + ((SE(\beta))^2(2N)eaf(1 - eaf))}$$

where $\beta$ and SE reflect the beta and standard error of effect estimates representing the SD change in relative abundance of bacteria, N represents the sample size of the mGWAS and EAF represents the effect allele frequency. Where the EAF was not available (i.e. oesophageal cancer GWAS), the $R^2$ was calculated using the "*add_rsq*" function in the *TwoSampleMR* package, which uses the following formula:

$$\frac{\left(\frac{\beta}{SE}\right)\hat{}2}{\left(\frac{\beta}{SE}\right)^2 + (n-1)}$$

The F-statistic (i.e., an indicator of weak instrument bias) was then calculated as follows:

$$F = (R^2(N - 1 - k))/((1 - R^2)k)$$

where $R^2$ is the proportion of variance explained in the microbial trait by each SNP, $N$ is the sample size of the mGWAS, and $k$ is the number of SNPs included in the instrument. The F-statistic was generated for each SNP separately (therefore, k = 1 in the above calculation). Effect estimates obtained from two-sample MR analyses represent the OR for cancer risk for each rank normalized SD higher abundance of each continuous microbial trait (MiBioGen microbial traits and the Hughes traits denoted with "RNT") and the OR for cancer risk for an approximate doubling of the genetic liability to presence (versus absence) of each binary microbial trait (those denoted with "HB").

In this study, to obtain 80% power to detect real causal effects, using an alpha level of 0.05 and an $R^2$ of 0.01, a proportion of 0.2 cases to controls in the study would require a sample size of 131862 individuals to detect an odds ratio of 1.2.

**Sensitivity analyses**

Assessing the evidence of a causal relationship between the gut microbiome and cancer risk, there are three MR assumptions that must be met: (1) the relevance assumption (the genetic instruments used to proxy the exposure are strongly associated with that exposure), (2) the independence assumption (there is no confounding between the genetic instrument(s) used to proxy the exposure and the outcome) and (3) the exclusion restriction assumption (the genetic instruments used to proxy the exposure have no effect on the outcome other than through the exposure).(41, 42, 45) To ensure homogeneity across exposure and outcome datasets and thus lowering the likelihood of violations of the second MR assumption, we used independent SNPs associated with our exposures of interest and data that had adjusted for genetic principal components and were of European ancestry. Microbial traits for which there was evidence of a causal effect on any cancer outcome were then followed up with a series of analyses that assessed the robustness of our findings to violations of these assumptions.

In addition to testing for instrument strength (i.e., via the F-statistic) and for possible reverse causality (i.e., through the Steiger test), the below sections describe the sensitivity analyses conducted to test the robustness of results found in main analyses. Namely, the creation of European-only data from the MiBioGen consortium(5) genetic colocalisation analyses, the manual exploration of pleiotropy, two-sample MR using a lenient p-value threshold to select genetic instruments, the Steiger test and filtering, and formal reverse MR analyses.

*European-only mGWAS data from MiBioGen*

To evaluate potential bias through population stratification, sensitivity analyses were performed on a newly generated set of MiBioGen summary statistics, subsetting the cohorts included in the meta-analysis to those of European ancestry. The meta-analysis followed the same pipeline as detailed on the MiBioGen GWAS publication (46, 47) (https://github.com/alexa-kur/miQTL_cookbook), where summary statistics of the bacterial traits were generated per cohort through an adjusted spearman correlation. The summary statistics of each cohort were then harmonised and combined through and inverse variance weighted (IVW) meta-analysis. The authors of MiBioGen supplied the necessary scripts and data to restrict analyses to a total of 17 European cohorts and ~14,000 individuals. Following removal of non-European or multi-ancestry cohorts, all 450 bacterial traits analysed in the original publication were regenerated using Java 1.8.0_74 and R 4.4.1.

*Colocalisation*

Colocalisation analyses were conducted using the 'coloc' R package (version 5.2.3).(48) The prior probabilities of the SNP being associated with the exposure, the outcome or both traits were specified as $1x10^{-04}$ and $1x10^{-05}$, respectively, as previously specified.(45) The continuous exposures (i.e., the relative abundance of microbial traits) were specified as continuous using the 'quant' option, while the Hughes binary traits (presence vs. absence of microbial traits) were specified as binary using the 'cc' option. The outcome (i.e., overall cancer risk) was binary using the 'cc' option. Bayes factor computation was used to generate 5 posterior probabilities (H0-H4) characterised by the following outcomes: (H0) neither trait has a genetic association in the region; (H1) only the gut microbiome has a genetic association in the region; (H2) only overall cancer risk has a genetic association in the region; (H3) both traits are associated but have different causal variants and (H4) both traits are associated and have the same causal variant. Whilst the commonly used posterior probability threshold across hypotheses is >0.80, we compared relative probabilities to assess evidence that the genetic variant was more predominantly associated with the exposure than with the outcome (H1 vs. H2) and that there was a shared (not different) causal variant in the region between each microbial trait and cancer risk (H4 vs. H3). Genetic variants ± 1 Mb of the lead SNP associated with any microbial trait for which there was evidence for a causal impact on overall cancer risk in our main MR analyses were extracted from Flemish Gut Flora Project (FGFP), and MiBioGen datasets and each cancer GWAS. Regional association plots for the microbial traits from the Hughes et al.(1) GWAS were generated using the locuscompare() function in the locuscomparer R package (Version 1.0.0), while the regional association plots for the microbial traits from the MiBioGen consortium(5) were generated using code available on GitHub.(49)

*Manual exploration of pleiotropy*

Summary-level GWAS data collated by the IEU Open GWAS(9, 38) were searched in a phenome-wide association analysis approach to identify whether any SNP used as an instrument for those microbial traits had been reported to be associated with cancer or any other trait that could be an independent cause of cancer. A lenient p-value threshold of $1x10^{-05}$ was set as the multiple testing threshold at which we defined evidence for an association between the SNP and any disease or trait in the IEU Open GWAS.

These analyses were performed in R using the phewas function in the R package *ieugwasr* (version 1.0.2), which searched for each SNP in the IEU Open GWAS database.(50) This was important to assess whether any causal relationships found between microbial traits and cancer were affected by horizontal pleiotropy. If some or all of the genetic instruments that are robustly associated with the exposure are also associated with other traits (or the outcome itself), independently of the exposure, then the MR estimate may reflect the effect of these traits on cancer and not the effect of the exposure of interest (i.e., horizontal pleiotropy). However, this analysis is limited in its ability to distinguish this from vertical pleiotropy (i.e., the causal mechanism by which the exposure influences the outcome). Since there were only singular instruments for each microbial trait, pleiotropy-robust methods that usually require multiple genetic variants were not possible to apply within this context.

*Two-sample MR using a lenient p-value threshold to select genetic instruments*

To allow the use of more formal pleiotropy-robust methods that require multiple genetic instruments, a more lenient *p*-value threshold of $1x10^{-05}$ was used to expand the number of SNPs associated with each of the microbial traits in the main analyses, reflecting the p-value threshold commonly used in the literature.(51, 52) However, a p-value of $1x10^{-05}$ is likely to induce weak instrument bias and violate several MR assumptions. To counter this, and to reduce heterogeneity in the effect of each SNP on the exposure, we selected SNPs that are directionally consistent across the comprising cohorts in the Hughes and MiBioGen mGWASs respectively(1, 5). SNPs were selected based on a *p*-value threshold of $1x10^{-05}$ and clumped (r-squared = 0.001, kb = 10000, P-value significance level = 1) to obtain independent SNPs.

When using the MiBioGen mGWAS data in these analyses, we used information from only the European-ancestry cohorts to reduce heterogeneity in the effect of each SNP on the exposure that may have arisen from differences in genetic architecture between ancestral groups.

Where multiple SNPs associated with each microbial trait were identified, the IVW method was used as was done in the primary analysis. The IVW method meta-analyses effect estimates across all SNPs weighted by the inverse variance of the SNP-outcome association using fixed effects.(45) Then, to test for horizontal pleiotropy using a larger number of SNPs, the weighted median,(53) weighted mode(54), and MR-Egger(55) regression methods were applied and consistency of effect estimates were compared to those obtained from the IVW method.

The weighted median requires that only half the SNPs are valid instruments (i.e., exhibiting no horizontal pleiotropy, no confounders of the instrument-outcome association and a robust association with the exposure) for the causal effect estimate to be unbiased.(53) The mode-based estimator clusters the SNPs into groups based on similarity of causal effects and returns the causal effect estimate based on the cluster that has the largest number of SNPs.(54) The weighted mode introduces an extra element like the IVW and weighted median estimators, weighting the contribution of each SNP to the clustering by the inverse variance of its outcome effect. The weighted mode estimator assumes that the most frequent estimates come from valid instruments.

The MR-Egger is sensitive to invalid SNPs, especially where there are only few SNPs.(55) The MR-Egger method relaxes the "no horizontal pleiotropy" assumption by allowing a non-zero intercept in the relationship between multiple SNP-outcome and SNP-exposure associations.(55) The presence of directional horizontal pleiotropy is tested formally by the intercept. A causal effect between the exposure and the outcome is indicated by the slope of the MR-Egger regression between multiple SNP-outcome and SNP-exposure associations and is considered unbiased assuming any horizontal pleiotropic effects are not correlated with the SNP-exposure effects (i.e., strength of the instrument). Visual inspection of MR forest plots, scatter plots, leave-one SNP out plots and funnel plots created using the *TwoSampleMR* R package (version 0.6.8) were used to determine any violations of the third MR assumption(44).

Steiger test and filtering

The Steiger test is used to provide evidence for the most plausible direction of causality given the instrument strength of the genetic instruments associated with the exposure in relation to the exposure and outcome. The Steiger test assumes that a valid genetic instrument should explain more variance in the exposure than the outcome and identifies those genetic variants that do not satisfy this criterion (i.e., where the genetic variants explain more variation in the outcome and thus could indicate reverse causation, where the SNPs influence the outcome which, in turn, influences the exposure). Where the Steiger test provided such evidence, we performed Steiger filtering (i.e., removing these SNPs) and repeated analyses (i.e., where there were multiple SNPs available in the instrument set).

*Reverse two-sample Mendelian randomization*

Where there was evidence of a causal effect in the main analyses, MR analyses were performed in the reverse direction, where each cancer was the exposure and the microbial traits were the outcomes. In this analysis, SNPs associated with each cancer phenotype at the genome-wide significance threshold ($P<5x10^{-8}$) were obtained from the largest and most recent GWAS meta-analysis of each cancer and then clumped (r-squared = 0.001, kb = 10000, P-value significance level = 1) to obtain independent SNPs. Summary statistics for cancer-related SNPs were extracted from the Hughes et al. and MiBioGen mGWASs,(1, 5) respectively, for the relevant microbial traits. Further details of the cancer GWASs used can be found above in the "Data" section of the Supplementary Methods and Supplementary Table 4.

MR analyses were undertaken the same as described in the previous section, using *TwoSample* MR package (version 0.6.8) using the IVW(56), weighted median(53), weighted mode(53) and MR-Egger(55) methods. Violations of the third MR assumption were visually assessed by funnel, forest, scatter and leave-one-out plots, and tests of heterogeneity of effects between the SNPs using Cochran's Q statistic(44, 57, 58).

### SUPPLEMENTARY FIGURES

**Supplementary Figure 1. Colocalisation results for lead SNP (rs13207588) associated with G.Parabacteroides.RNT and lung cancer**

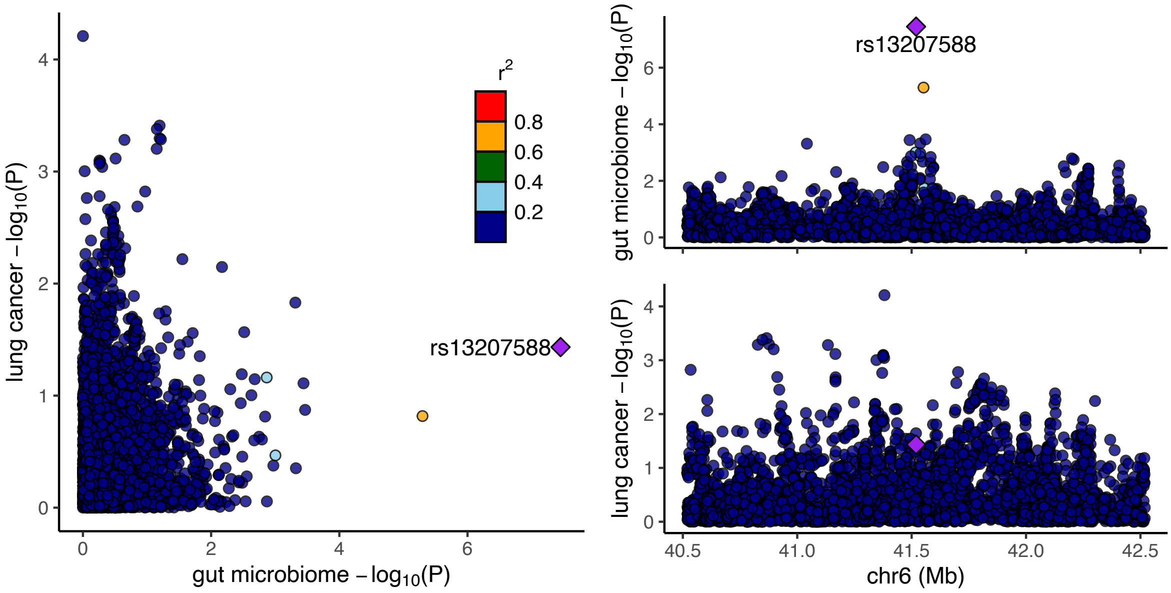


*G = genus; GWAS = genome-wide association study; RNT= reverse normal transformed; SNP = single nucleotide polymorphism. Regional association plots, generated from LocusCompareR, showing the −log10(P-value) where the lead SNP (rs13207588) associated with the relative abundance of bacteria in the genus Parabacteroides (G.Parabacteroides.RNT) is represented by a purple diamond. These plots were created using the Flemish Gut Flora Project (FGFP) and Trans-disciplinary Research Into Cancer of the Lung (TRICL) (accessed by IEU Open GWAS using ID: ieu-a-987) full summary-level GWAS data for microbial traits and lung cancer, respectively.*

**Supplementary Figure 2. Colocalisation results for lead SNP (rs11788336) associated with G.unclassified.P.Firmicutes.RNT and prostate cancer**

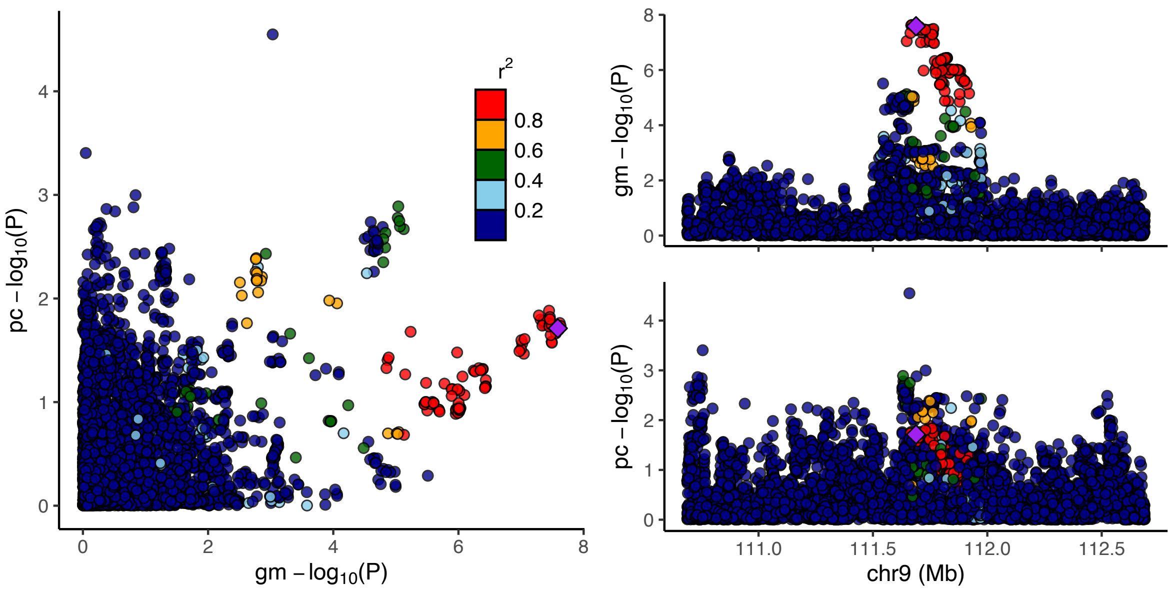


*G = genus; gm = gut microbiome; GWAS = genome-wide association study; P = phylum; pc = prostate cancer; RNT = reverse normal transformed; SNP = single nucleotide polymorphism. Regional association plots, generated from LocusCompareR showing the −log10(P-value) where the lead SNP (rs11788336) associated with the relative abundance of an unclassified group of bacteria in the Firmicutes phylum (G.unclassified.P.Firmicutes.RNT is represented by a purple diamond. These plots were created using the Flemish Gut Flora Project (FGFP) and the Prostate Cancer Association Group to Investigate Cancer Associated Alterations in the genome (PRACTICAL) (accessed by IEU Open GWAS using ID: ebi-a-GCST006085) full summary-level GWAS data for microbial traits and oesophageal cancer, respectively.*

**Supplementary Figure 3. Colocalisation results for lead SNP (rs35866622) associated with genus.Ruminococcustorquesgroup.id.14377 and oesophageal cancer**

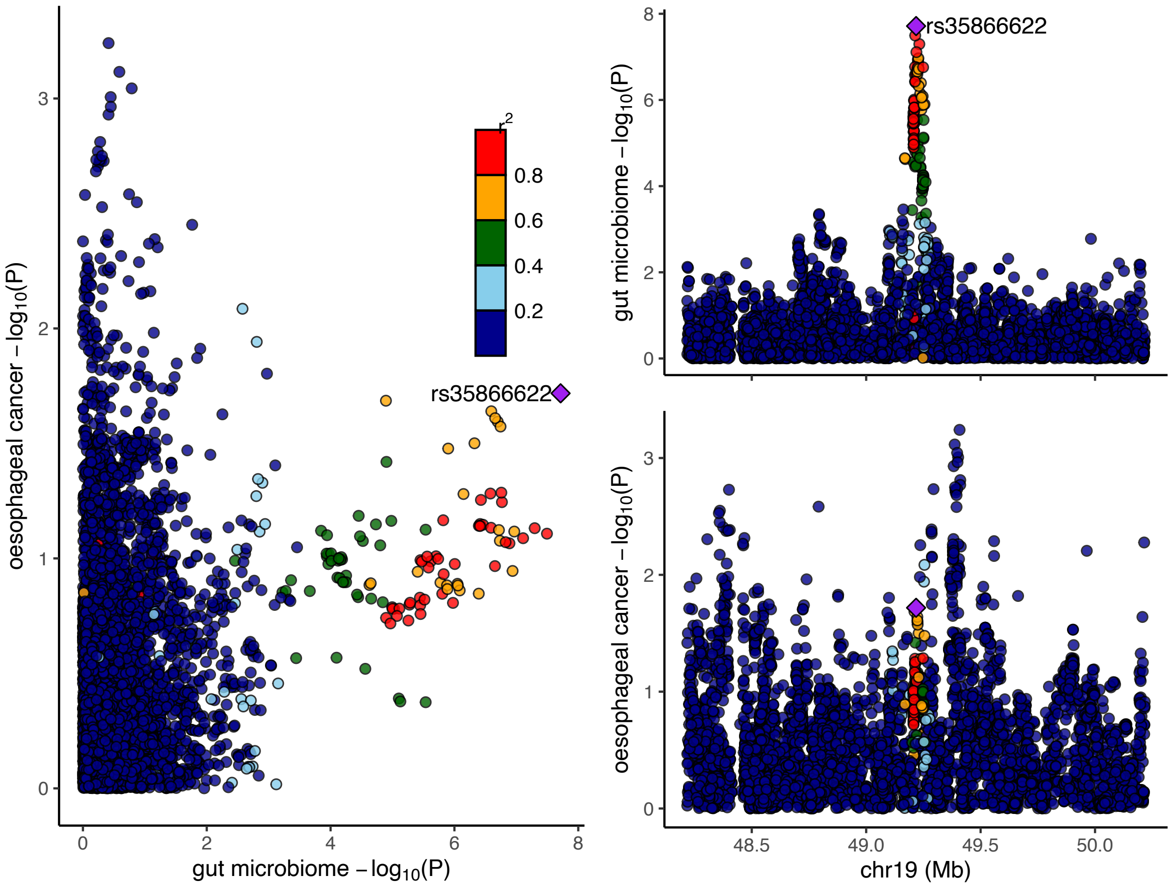


*GWAS = genome-wide association study; SNP = single nucleotide polymorphism. Regional association plots, from LocusCompareR showing the −log10(P-value) where the lead SNP (rs35866622) associated with the relative abundance of bacteria in the genus Ruminococcus Torques group (genus.Ruminococcustorquesgroup.id.14377) is represented by a purple diamond. These plots were created using the MiBioGen consortium and the Schröder et al. (2023) full summary-level GWAS data for microbial traits and oesophageal cancer, respectively.*

**Supplementary Figure 4. Colocalisation results for lead SNP (rs11098863) associated with genus.Enterorhabdus.id.820 and breast cancer**

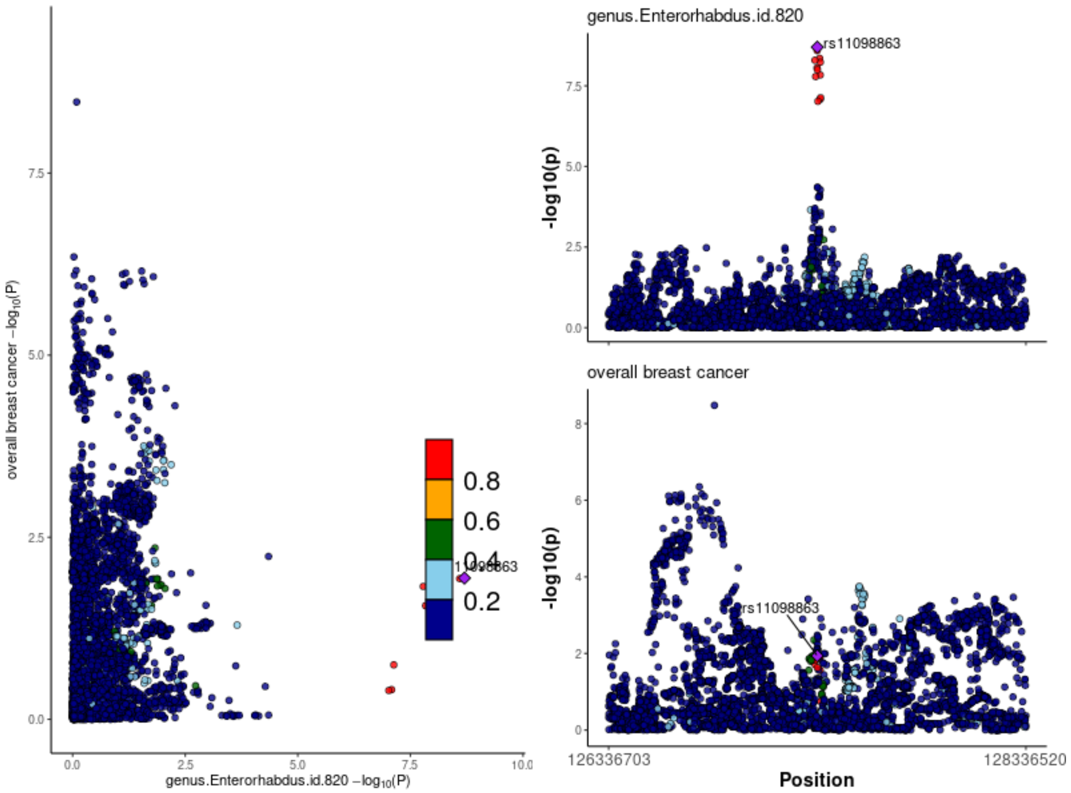


*GWAS = genome-wide association study; SNP = single nucleotide polymorphism. Regional association plots, generated from code available on GitHub(49), showing the −log10(P-value) where the lead SNP (rs11098863) associated with the relative abundance of bacteria in the genus Enterorhabdus (genus.Enterorhabdus.id.820) is represented by a purple diamond. These plots were created using the MiBioGen consortium and the Breast Cancer Association Consortium (BCAC) full summary-level GWAS data for microbial traits and breast cancer, respectively.*

**Supplementary Figure 5. Colocalisation results for lead SNP (rs4428215) associated with family.Oxalobacteraceae.id.2966 and breast cancer**

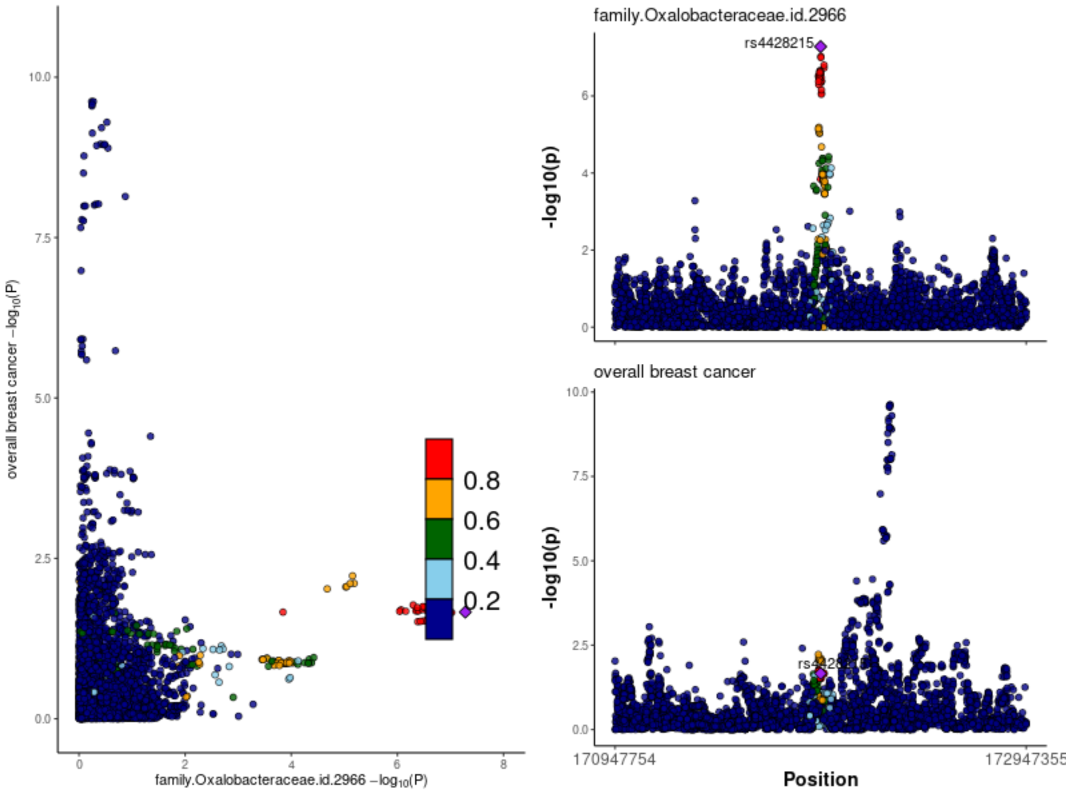


*GWAS = genome-wide association study; SNP = single nucleotide polymorphism. Regional association plots, generated from code available on GitHub(49), showing the −log10(P-value) where the lead SNP (rs4428215) associated with the relative abundance of bacteria in the family Oxalobacteraceae (family.Oxalobacteraceae.id.2966) is represented by a purple diamond. These plots were created using the MiBioGen consortium and the Breast Cancer Association Consortium (BCAC) full summary-level GWAS data for microbial traits and breast cancer, respectively.*

**Supplementary Figure 6. Colocalisation results for lead SNP (rs7221249) associated with genus.Erysipelatoclostridium.id.11381 and pancreatic cancer**

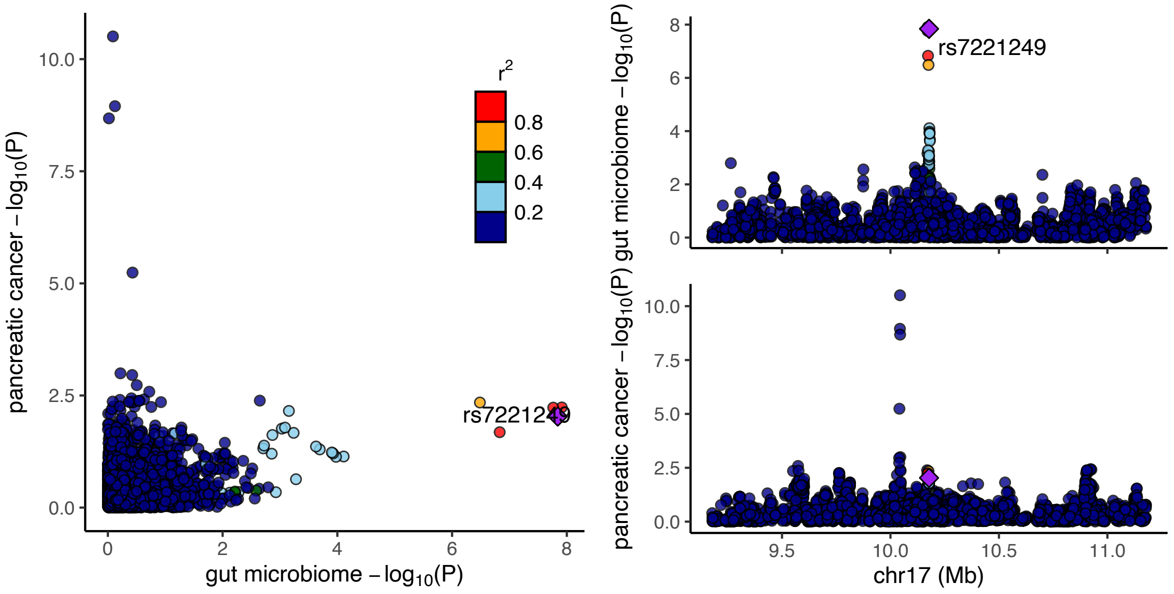


*GWAS = genome-wide association study; SNP = single nucleotide polymorphism. Regional association plots, generated from code available on GitHub(49), showing the −log10(P-value) where the lead SNP (rs7221249) associated with the relative abundance of bacteria in the genus Erysipelatoclostridium (genus.Erysipelatoclostridium.id.11381) is represented by a purple diamond. These plots were created using the MiBioGen consortium and the Klein et al. (2018) Pan Scan 3 + C4 full summary-level GWAS data for microbial traits and pancreatic cancer, respectively.*

**Supplementary Figure 7. Colocalisation results for lead SNP (rs13207588) associated with G.Parabacteroides.RNT and prostate cancer**

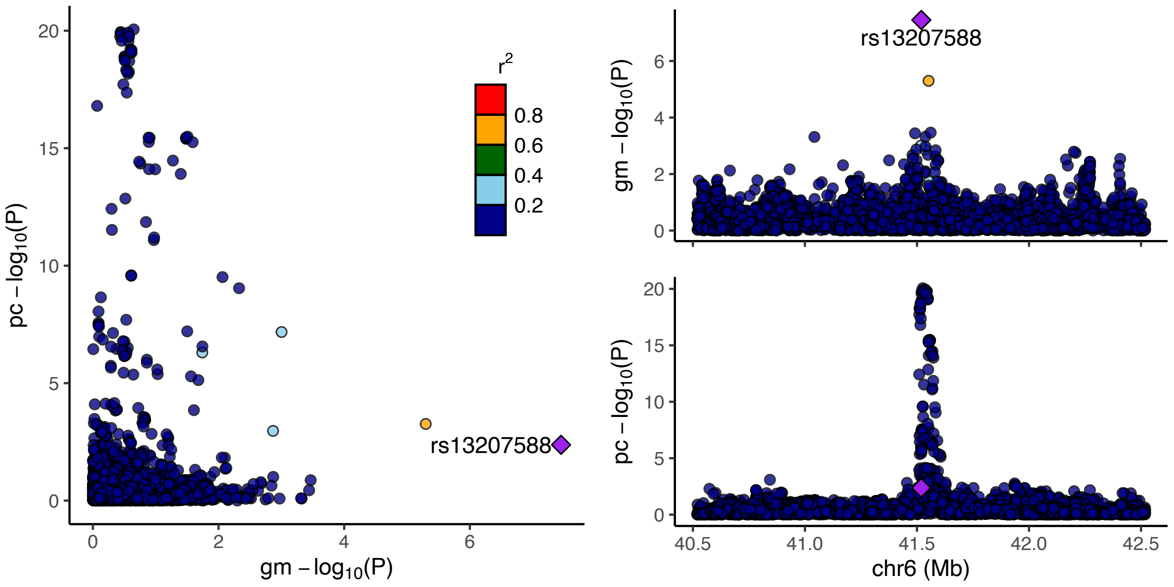


*G = genus; GWAS = genome-wide association study; RNT = reverse normal transformed; SNP = single nucleotide polymorphism. Regional association plots, generated from LocusCompareR showing the −log10(P-value) where the lead SNP (rs13207588) associated with the relative abundance of bacteria in the genus Parabacteroides (G.Parabacteroides.RNT) is represented by a purple diamond. These plots were created using the Flemish Gut Flora Project (FGFP) and the Prostate Cancer Association Group to Investigate Cancer Associated Alterations in the genome (PRACTICAL) (accessed by IEU Open GWAS using ID: ebi-a-GCST006085) full summary-level GWAS data for microbial traits and oesophageal cancer, respectively.*

**Supplementary Figure 8. Colocalisation results for lead SNP (rs4428215) associated with family.Oxalobacteraceae.id.2966 and colorectal cancer**

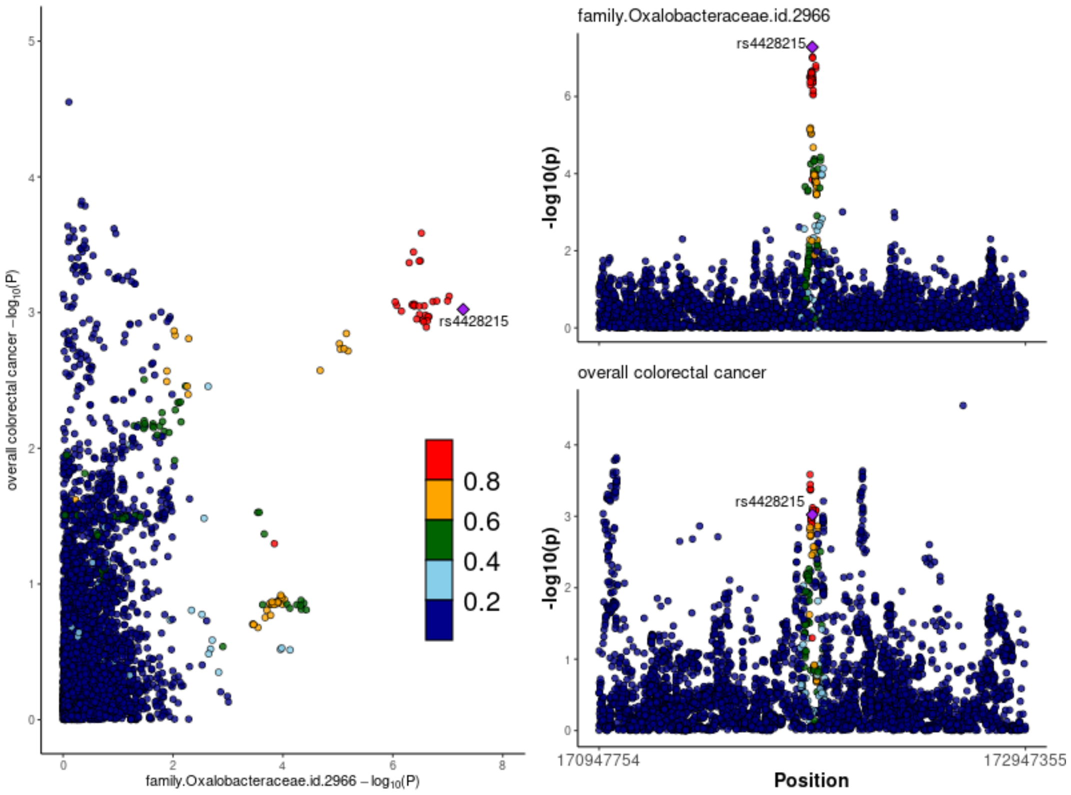


*GWAS = genome-wide association study; SNP = single nucleotide polymorphi. Regional association plots, generated from code available on GitHub(49), showing the −log10(P-value) where the lead SNP (rs4428215) associated with the relative abundance of bacteria in the family Oxalobacteraceae (family.Oxalobacteraceae.id.2966) is represented by a purple diamond. These plots were created using the MiBioGen consortium and the Genetic and Epidemiology of Colorectal Cancer Consortium (GECCO) (downloaded from [https://www.ebi.ac.uk/gwas/studies/GCST90255675](https://www.ebi.ac.uk/gwas/studies/GCST90255675)) full summary-level GWAS data for microbial traits and colorectal cancer, respectively.*

**Supplementary Figure 9. Colocalisation results for lead SNP (rs35866622) associated with genus.Ruminococcustorquesgroup.id.14377 and colorectal cancer**

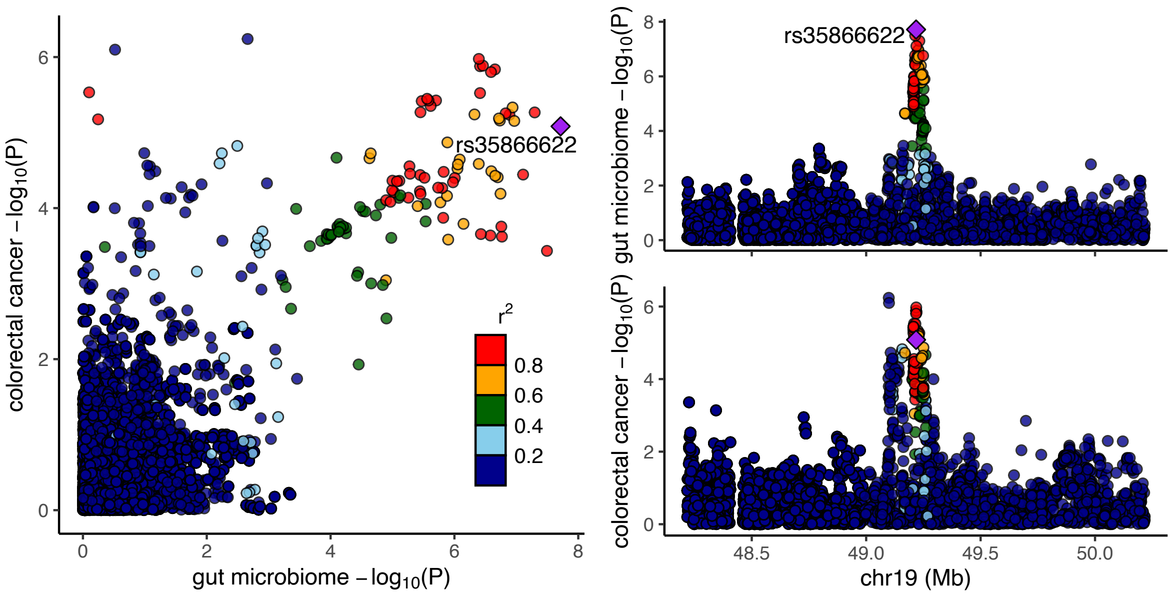


*GWAS = genome-wide association study; SNP = single nucleotide polymorphism. Regional association plots, generated from code available on GitHub(49), showing the −log10(P-value) where the lead SNP (rs35866622) associated with the relative abundance of bacteria in the genus Ruminococcus Torques group (genus.Ruminococcustorquesgroup.id.14377) is represented by a purple diamond. These plots were created using the MiBioGen consortium and the Genetic and Epidemiology of Colorectal Cancer Consortium (GECCO) (downloaded from [https://www.ebi.ac.uk/gwas/studies/GCST90255675](https://www.ebi.ac.uk/gwas/studies/GCST90255675)) full summary-level GWAS data for microbial traits and colorectal cancer, respectively.*

**Supplementary Figure 10. Colocalisation results for lead SNP (rs7570971) associated with phylum.Actinobacteria.id.400 and colorectal cancer**

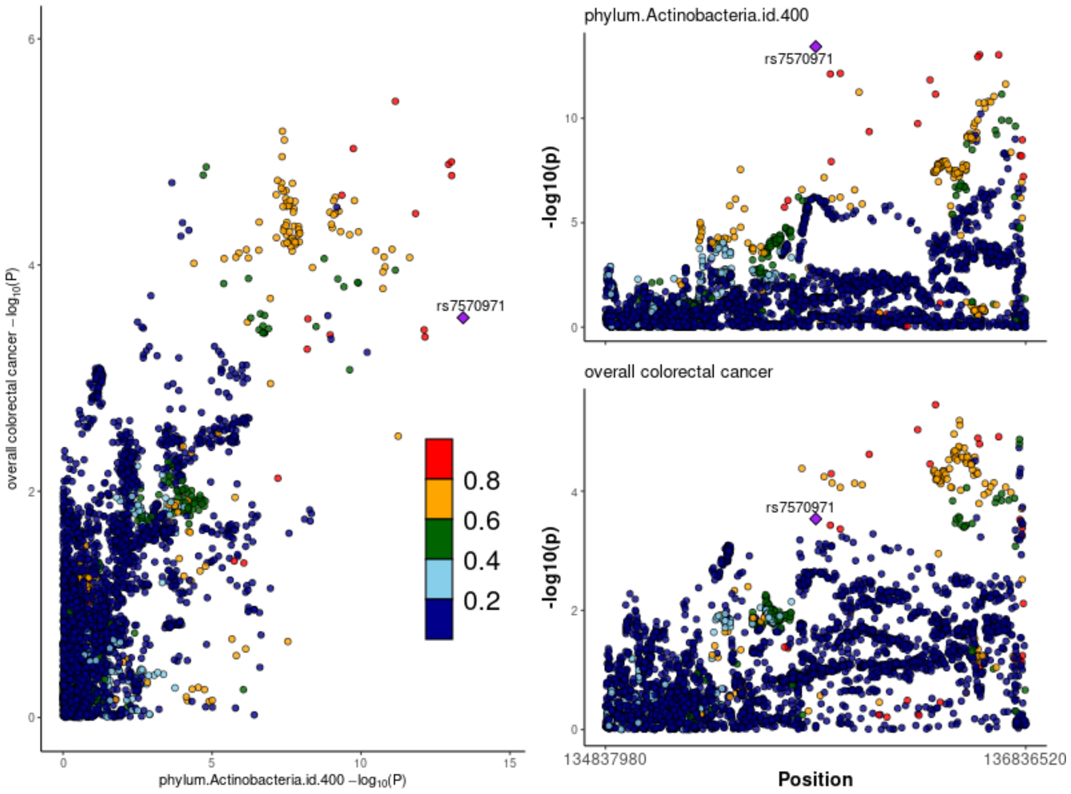


*GWAS = genome-wide association study; SNP = single nucleotide polymorphism. Regional association plots, generated from code available on GitHub(49), showing the −log10(P-value) where the lead SNP (rs7570971) associated with the relative abundance of bacteria in the phylum Actinobacteria (phylum.Actinobacteria.id.400) is represented by a purple diamond. These plots were created using the MiBioGen consortium and the Genetic and Epidemiology of Colorectal Cancer Consortium (GECCO) (downloaded from https://www.ebi.ac.uk/gwas/studies/GCST90255675) full summary-level GWAS data for microbial traits and colorectal cancer, respectively.*

**Supplementary Figure 11. Colocalisation results for lead SNP (rs182549) associated with class.Actinobacteria.id.419 and colorectal cancer**

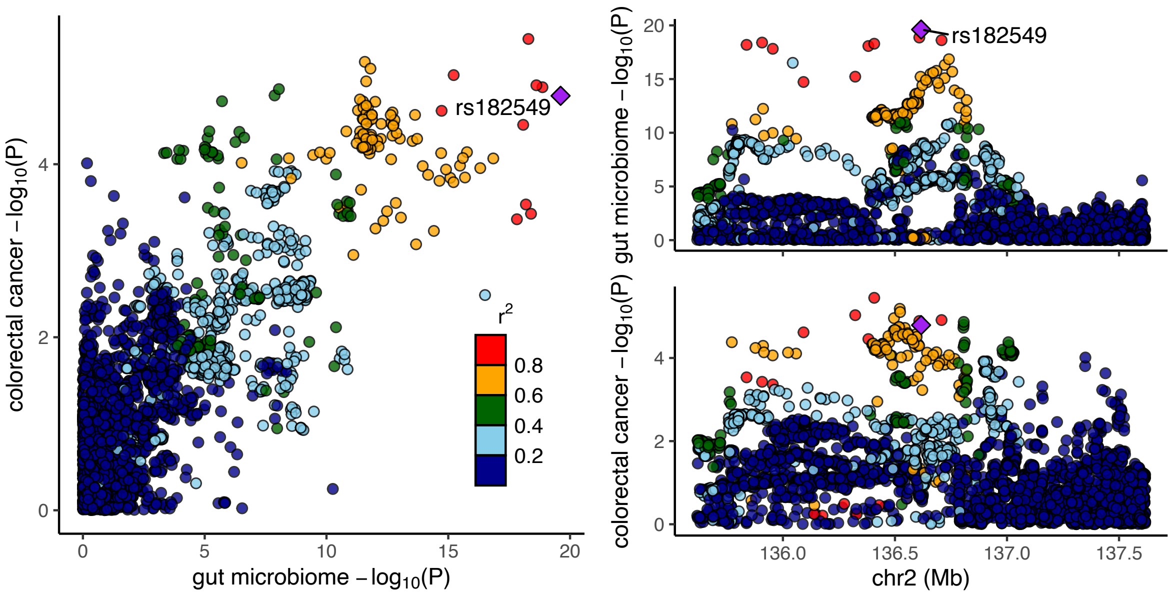


*GWAS = genome-wide association study; SNP = single nucleotide polymorphism. Regional association plots, generated from code available on GitHub(49), showing the −log10(P-value) where the lead SNP (rs182549) associated with the relative abundance of bacteria in the class Actinobacteria (class.Actinobacteria.id.419) is represented by a purple diamond. These plots were created using the MiBioGen consortium and the Genetic and Epidemiology of Colorectal Cancer Consortium (GECCO) (downloaded from [https://www.ebi.ac.uk/gwas/studies/GCST90255675](https://www.ebi.ac.uk/gwas/studies/GCST90255675)) full summary-level GWAS data for microbial traits and colorectal cancer, respectively.*

**Supplementary Figure 12. Colocalisation results for lead SNP (rs10805326) associated with genus.Intestinibacter.id.11345 and lung cancer**

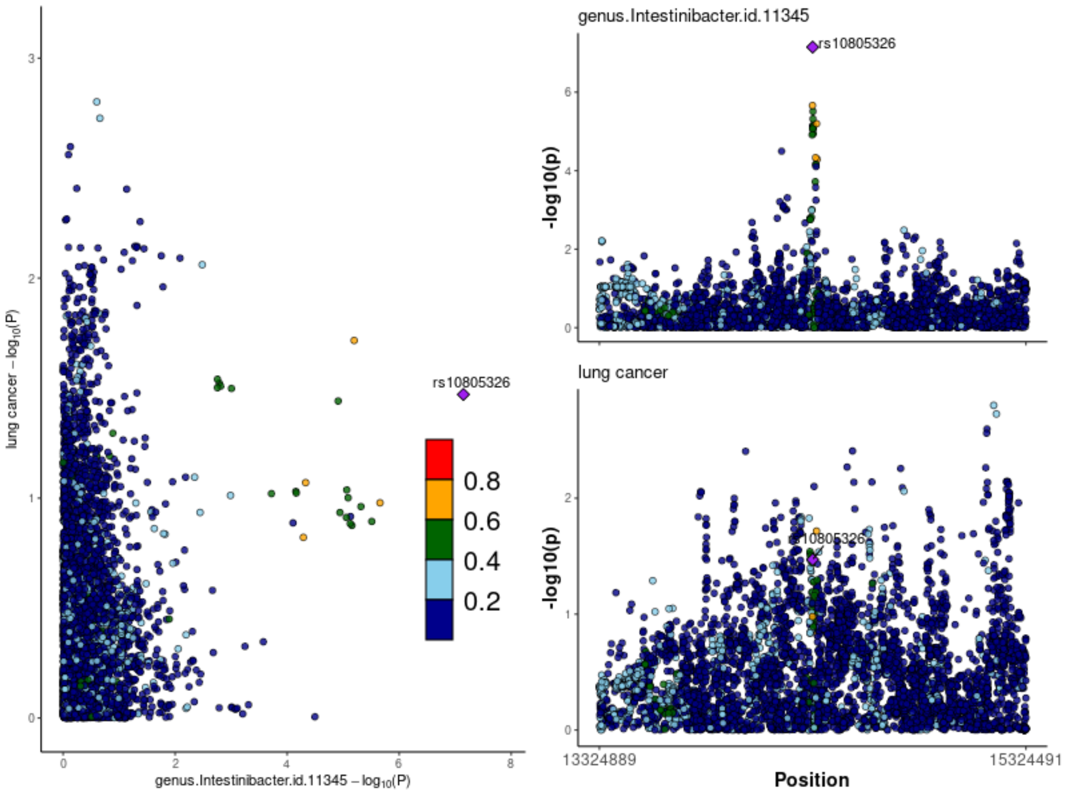


*GWAS = genome-wide association study; SNP = single nucleotide polymorphism. Regional association plots, generated from code available on GitHub(49), showing the −log10(P-value) where the lead SNP (*rs10805326*) associated with the relative abundance of bacteria in the genus* Intestinibacter (genus.Intestinibacter.id.11345*) is represented by a purple diamond. These plots were created using the MiBioGen consortium and Trans-disciplinary Research Into Cancer of the Lung (TRICL) (accessed by IEU Open GWAS using ID: ieu-a-987) full summary-level GWAS data for microbial traits and lung cancer, respectively.*

**Supplementary Figure 13. Colocalisation results for lead SNP (rs11110281) associated with genus.Streptococcus.id.1853 and lung cancer**

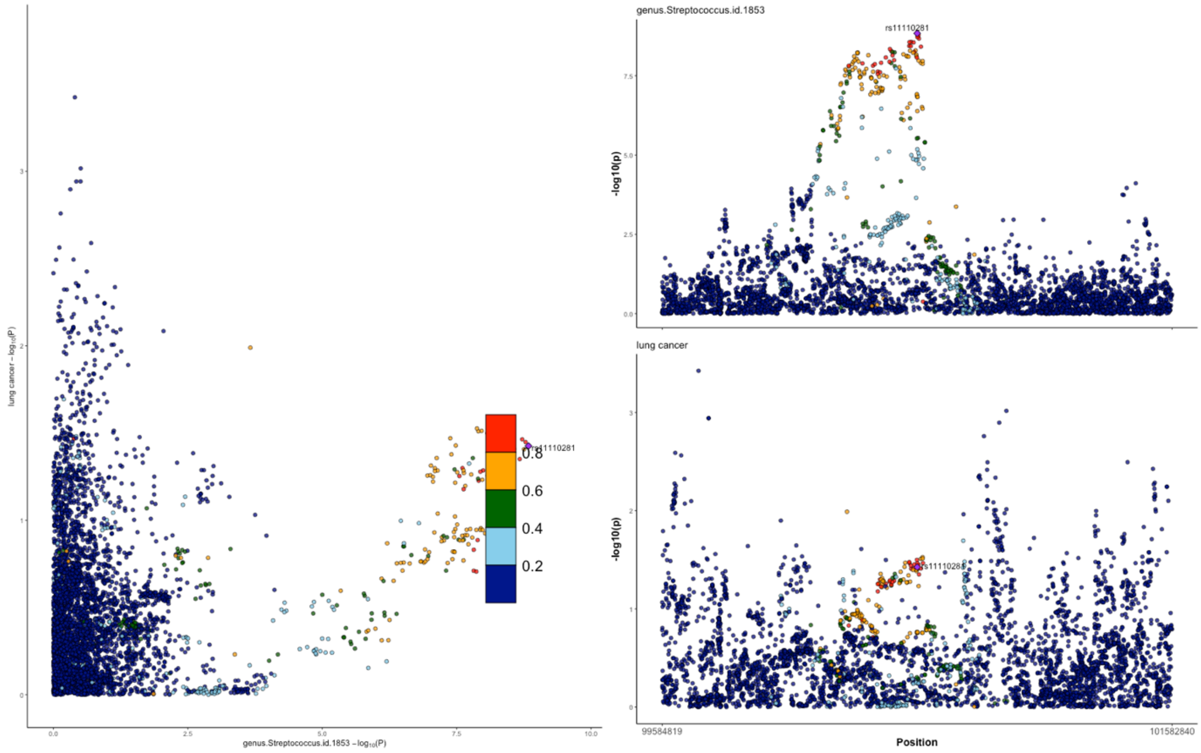


*GWAS = genome-wide association study; SNP = single nucleotide polymorphism. Regional association plots, generated from code available on GitHub(49), showing the −log10(P-value) where the lead SNP (rs11110281) associated with the relative abundance of bacteria in the genus* Streptococcus (genus.Streptococcus.id.1853*) is represented by a purple diamond. These plots were created using the MiBioGen consortium and Trans-disciplinary Research Into Cancer of the Lung (TRICL) (accessed by IEU Open GWAS using ID: ieu-a-987) full summary-level GWAS data for microbial traits and lung cancer, respectively.*

**Supplementary Figure 14. Colocalisation results for lead SNP (rs11110281) associated with family.Streptococcaceae.id.1850 and lung cancer**

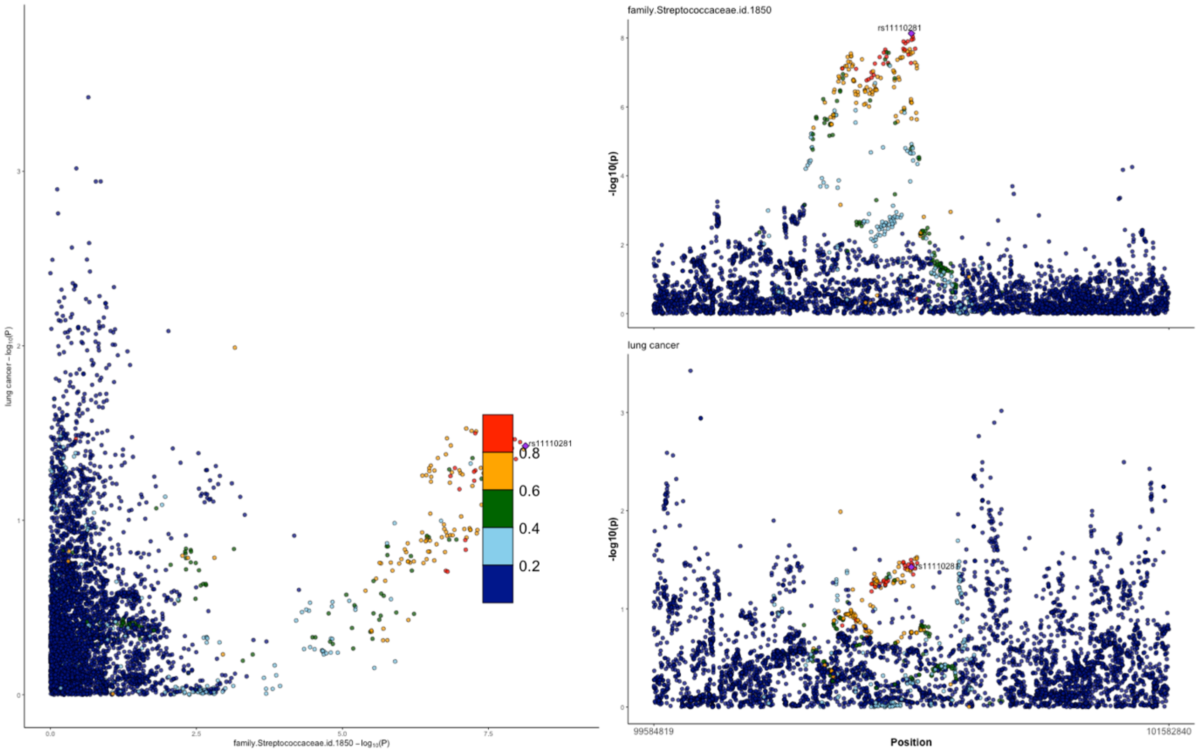


*GWAS = genome-wide association study; SNP = single nucleotide polymorphism. Regional association plots, generated from code available on GitHub(49), showing the −log10(P-value) where the lead SNP (rs11110281) associated with the relative abundance of bacteria in the family* Streptococcaceae (family.Streptococcaceae.id.1850*) is represented by a purple diamond. These plots were created using the MiBioGen consortium and Trans-disciplinary Research Into Cancer of the Lung (TRICL) (accessed by IEU Open GWAS using ID: ieu-a-987) full summary-level GWAS data for microbial traits and lung cancer, respectively.*

**Supplementary Figure 15. Colocalisation results for lead SNP (rs61841503) associated with family.Peptostreptococcaceae.id.2042 and pancreatic cancer**

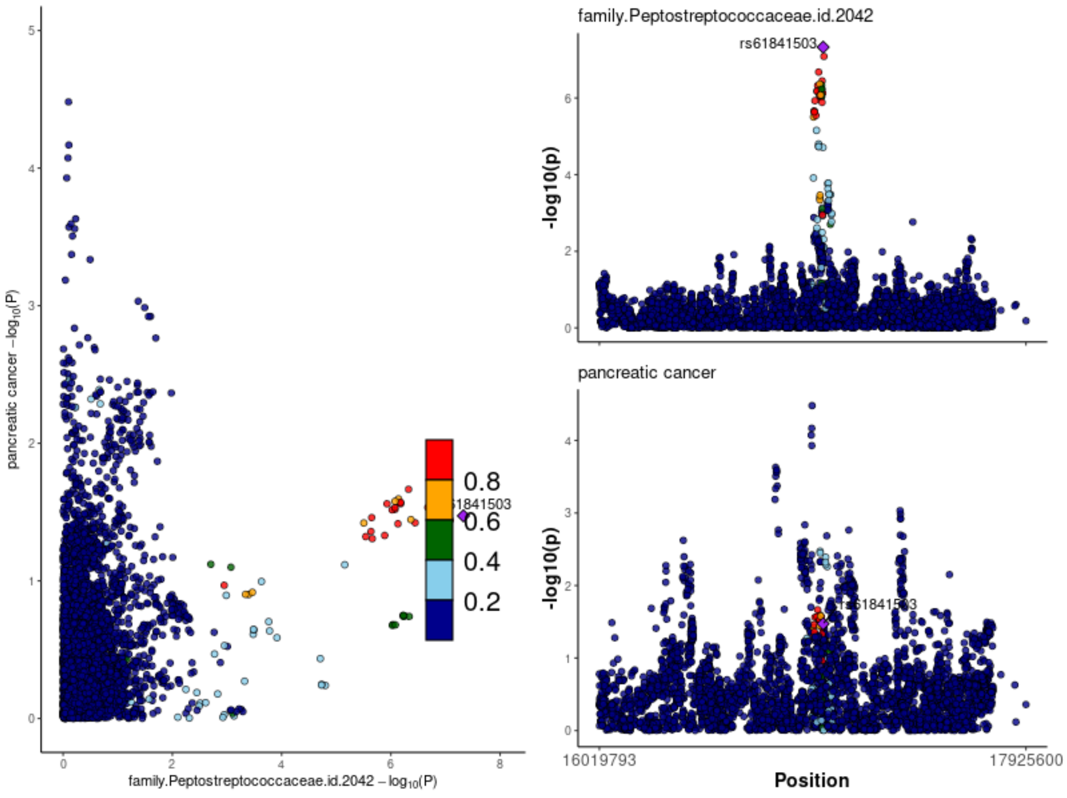


*GWAS = genome-wide association study; SNP = single nucleotide polymorphism. Regional association plots, generated from code available on GitHub(49), showing the −log10(P-value) where the lead SNP (rs61841503) associated with the relative abundance of bacteria in the family* Peptostreptococcaceae *(family.Peptostreptococcaceae.id.2042) is represented by a purple diamond. These plots were created using the MiBioGen consortium and the Klein et al. (2018) Pan Scan 3 + C4 full summary-level GWAS data for microbial traits and pancreatic cancer, respectively.*

**Supplementary Figure 16. Colocalisation results for lead SNP (rs61841503) associated with genus.Romboutsia.id.11347 and pancreatic cancer**

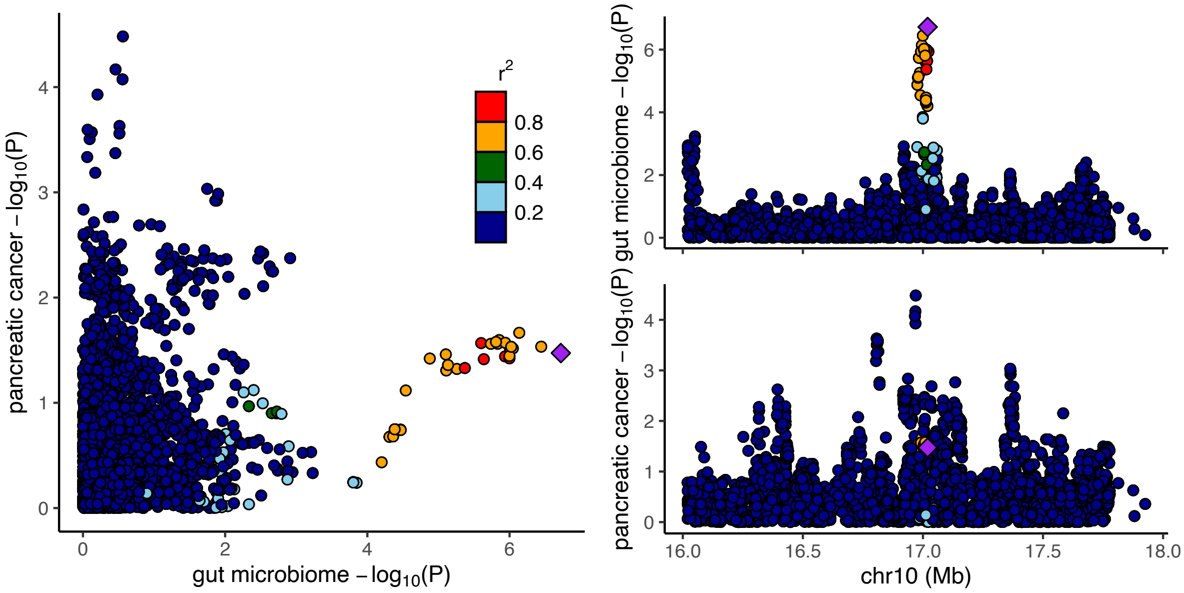


*SNP = single nucleotide polymorphism. Chr = chromosome. Regional association plots, generated from code available on GitHub(49), showing the −log10(P-value) where the lead SNP* (rs61841503*) associated with the relative abundance of bacteria in the genus Romboutsia (genus.Romboutsia.id.11347) is represented by a purple diamond. These plots were created using the MiBioGen consortium and the Klein et al. (2018) Pan Scan 3 + C4 full summary-level GWAS data for microbial traits and pancreatic cancer, respectively.*

**Supplementary Figure 17. Colocalisation results for lead SNP (rs35980751) associated with G.unclassified.F.Porphyromonadaceae.RNT and prostate cancer**

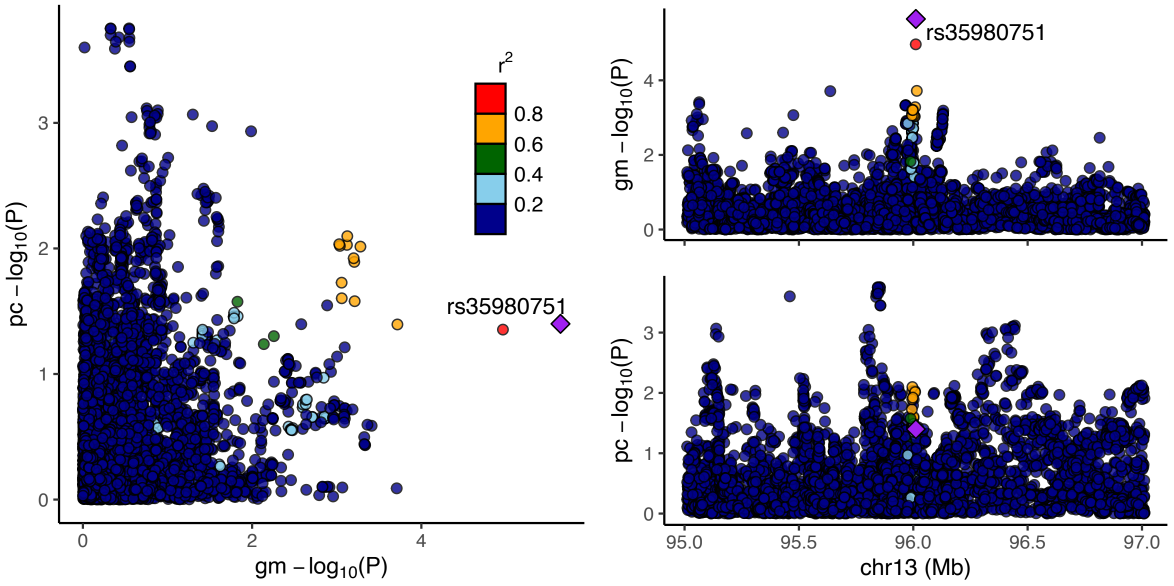


*F = family; G = genus; gm = gut microbiome; GWAS = genome-wide association study; pc = prostate cancer; RNT = reverse normal transformed; SNP = single nucleotide polymorphism. Regional association plots, generated from LocusCompareR showing the −log10(P-value) where the lead SNP (rs35980751) associated with the relative abundance of an unclassified group of bacteria in the Porphyromonadaceae family (G.unclassified.F.Porphyromonadaceae.RNT) is represented by a purple diamond. These plots were created using the Flemish Gut Flora Project (FGFP) and the Prostate Cancer Association Group to Investigate Cancer Associated Alterations in the genome (PRACTICAL) (accessed by IEU Open GWAS using ID: ebi-a-GCST006085) full summary-level GWAS data for microbial traits and oesophageal cancer, respectively.*

**Supplementary Figure 18. Colocalisation results for lead SNP (rs602075) associated with genus.Allisonella.id.2174 and prostate cancer**

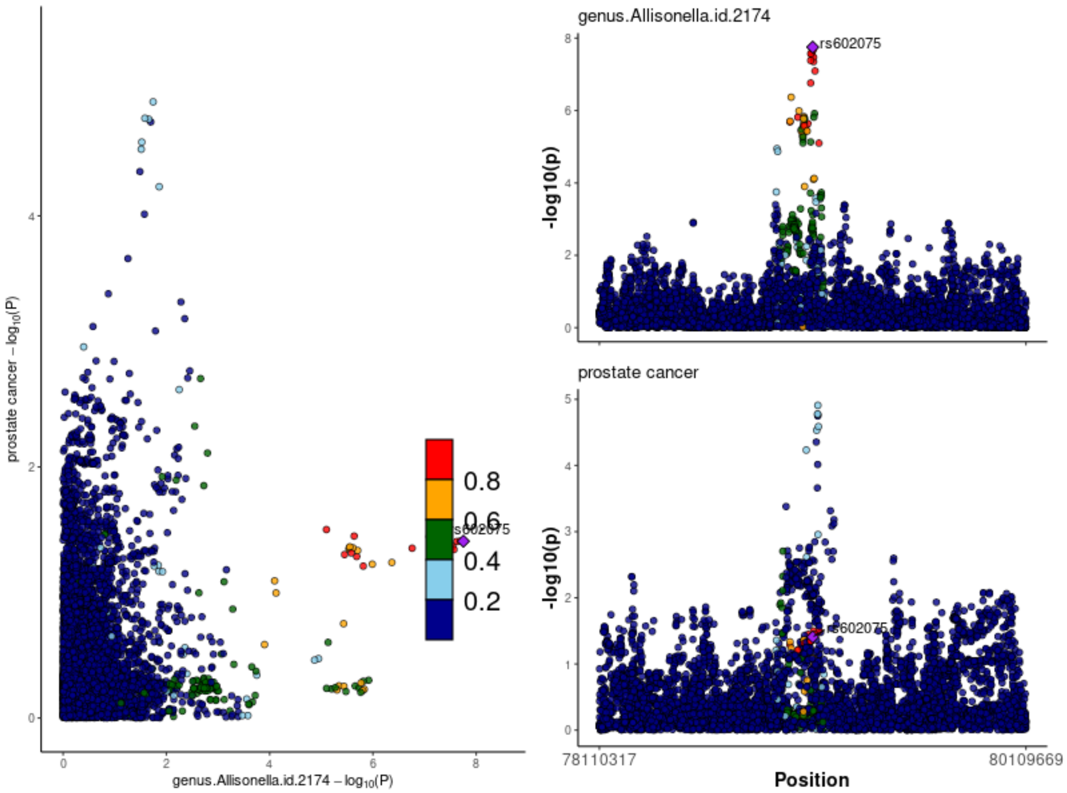


*GWAS = genome-wide association study; SNP = single nucleotide polymorphism. Regional association plots, generated from code available on GitHub(49), showing the −log10(P-value) where the lead SNP (rs602075) associated with the relative abundance of bacteria in the genus Allisonella (genus.Allisonella.id.2174) is represented by a purple diamond. These plots were created using the MiBioGen consortium and the Prostate Cancer Association Group to Investigate Cancer Associated Alterations in the genome (PRACTICAL) (accessed by IEU Open GWAS using ID: ebi-a-GCST006085) full summary-level GWAS data for microbial traits and prostate cancer, respectively.*

**Supplementary Figure 19. Colocalisation results for lead SNP (rs34656657) associated with G.Unclassified.P.Firmicutes.HB and ovarian cancer**

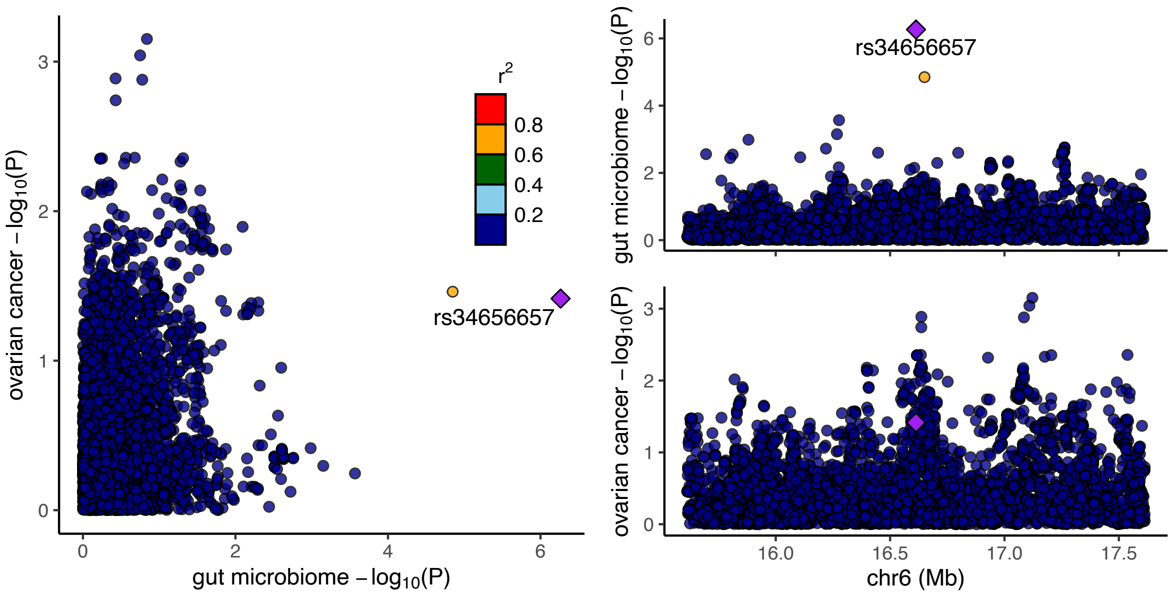


*G = genus; GWAS = genome-wide association study; P = phylum; HB = hurdle binary; SNP = single nucleotide polymorphism. Regional association plots, generated from LocusCompareR showing the −log10(P-value) where the lead SNP (rs34656657) associated with the presence (vs. absence) of an unclassified group of bacteria in the Firmicutes phylum (*G.Unclassified.P.Firmicutes.HB*) is represented by a purple diamond. These plots were created using the Flemish Gut Flora Project (FGFP) and the Ovarian Cancer Association Consortium (accessed by IEU Open GWAS using ID: ieu-a-1120) full summary-level GWAS data for microbial traits and oesophageal cancer, respectively.*

**Supplementary Figure 20. Colocalisation results for lead SNP (rs4428215) associated with family.Oxalobacteraceae.id.2966 and endometrial cancer**

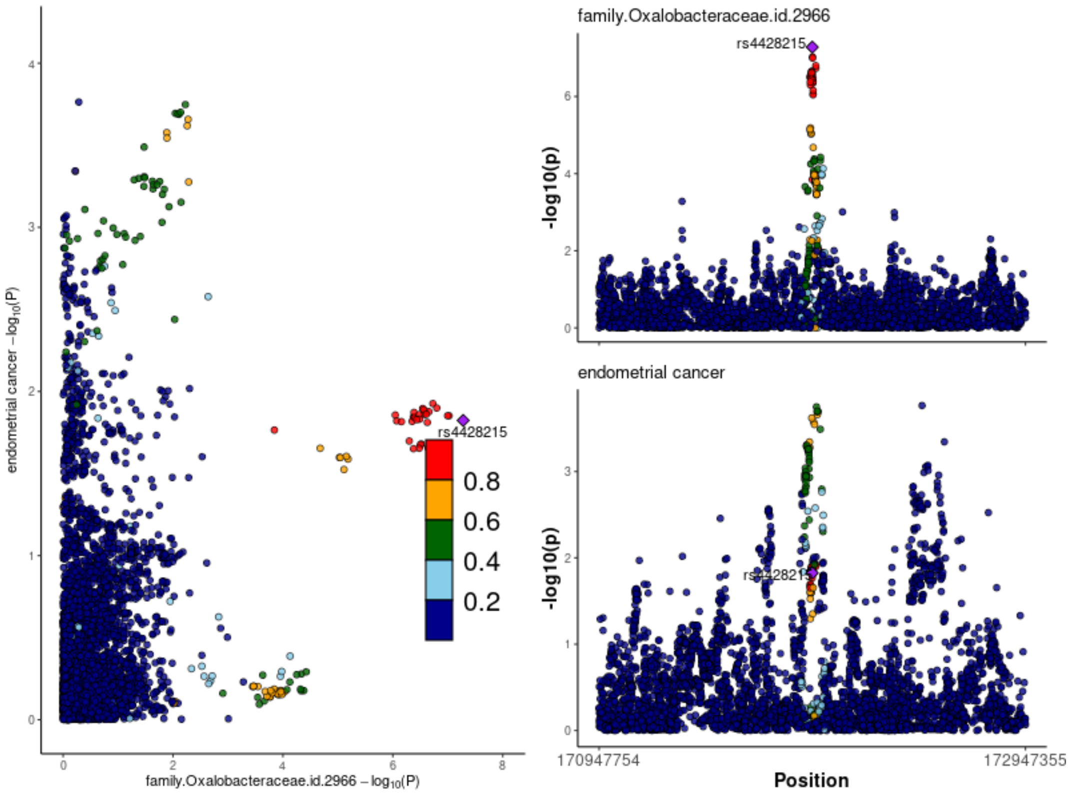


*GWAS = genome-wide association study; SNP = single nucleotide polymorphism. Regional association plots, generated from code available on GitHub(49), showing the −log10(P-value) where the lead SNP* (rs4428215*) associated with the relative abundance of bacteria in the family Oxalobacteraceae (family.Oxalobacteraceae.id.2966) is represented by a purple diamond. These plots were created using the MiBioGen consortium and the O'Mara et al. (2018) (accessed by IEU Open GWAS using ID: ebi-a-GCST006464) full summary-level GWAS data for microbial traits and endometrial cancer, respectively.*

**Supplementary Figure 21. Colocalisation results for lead SNP (rs11110281) associated with family.Streptococcaceae.id.1850 and endometrial cancer**

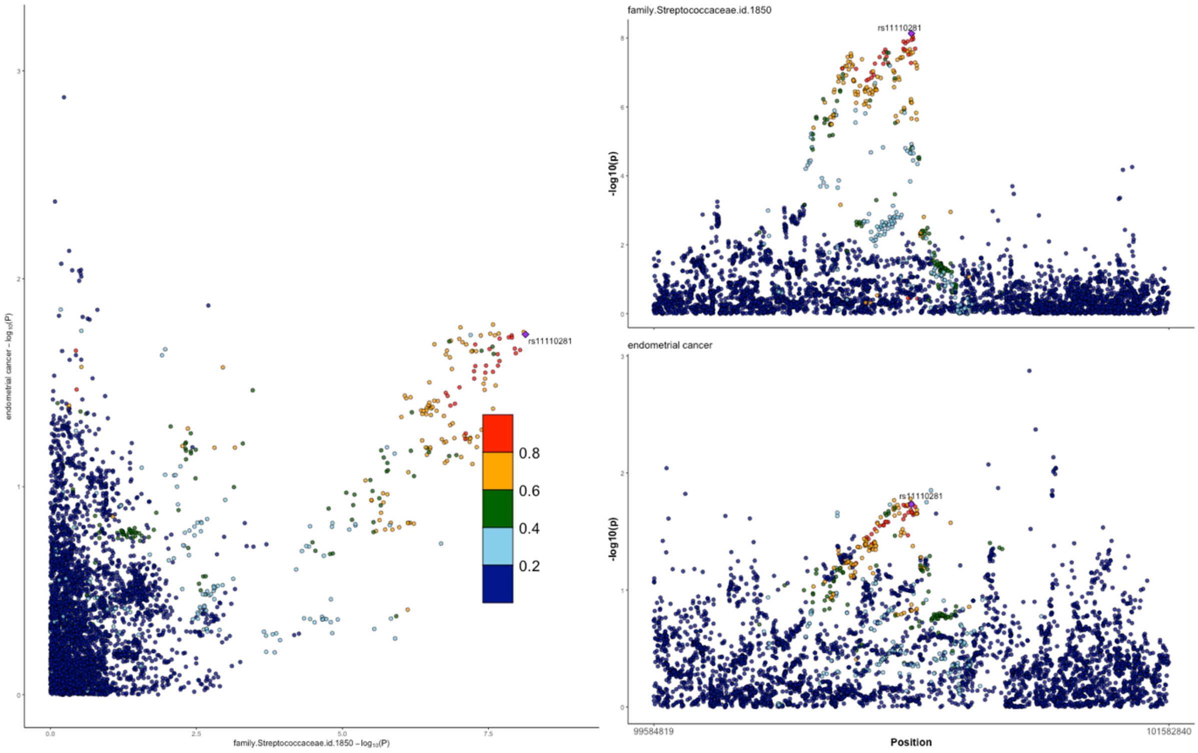


*GWAS = genome-wide association study; SNP = single nucleotide polymorphism. Regional association plots, generated from code available on GitHub(49), showing the −log10(P-value) where the lead SNP (rs11110281) associated with the relative abundance of bacteria in the family Streptococcaceae (family.Streptococcaceae.id.1850) is represented by a purple diamond. These plots were created using the MiBioGen consortium and the O'Mara et al. (2018) (accessed by IEU Open GWAS using ID: ebi-a-GCST006464) full summary-level GWAS data for microbial traits and endometrial cancer, respectively.*

**Supplementary Figure 22. Colocalisation results for lead SNP (rs11110281) associated with genus.Streptococcus.id.1853 and endometrial cancer**

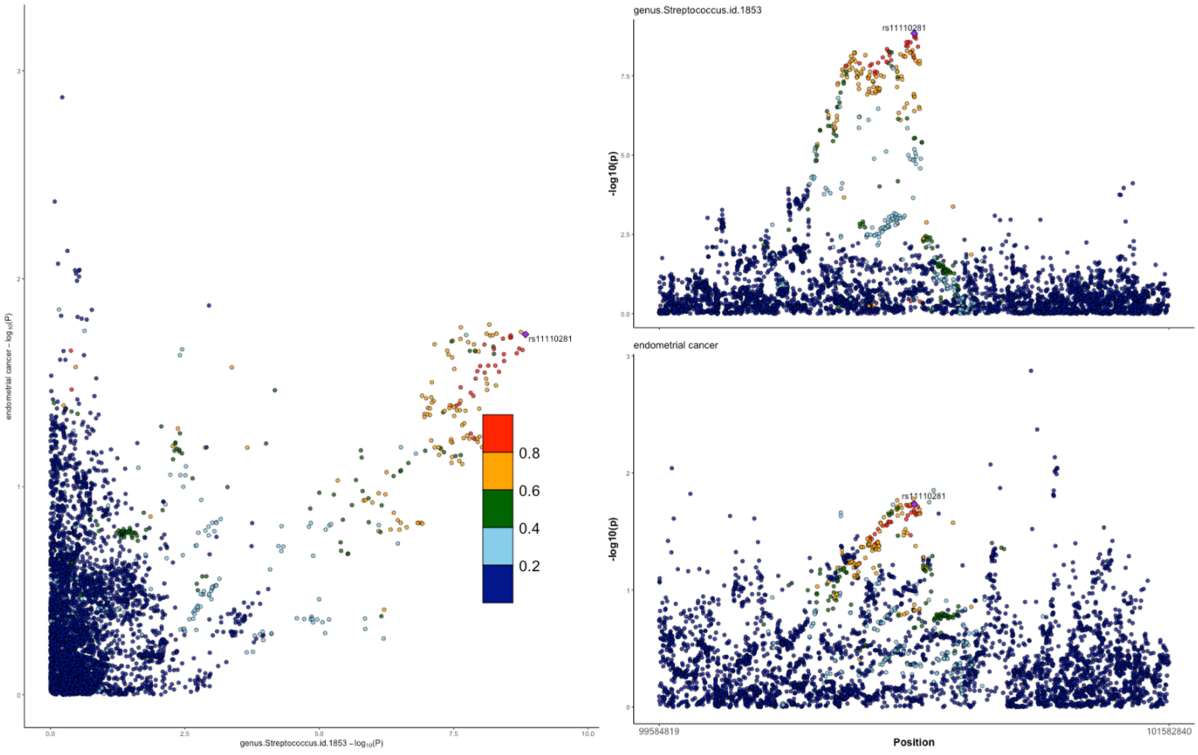


*GWAS = genome-wide association study; SNP = single nucleotide polymorphism. Regional association plots, generated from code available on GitHub(49), showing the −log10(P-value) where the lead SNP (rs11110281) associated with the relative abundance of bacteria in the genus Streptococcus (genus.Streptococcus.id.1853) is represented by a purple diamond. These plots were created using the MiBioGen consortium and the O'Mara et al. (2018) (accessed by IEU Open GWAS using ID: ebi-a-GCST006464) full summary-level GWAS data for microbial traits and endometrial cancer, respectively.*

**Supplementary Figure 23. Colocalisation results for lead SNP (rs11098863) associated with genus.Enterorhabdus.id.820 and endometrial cancer**

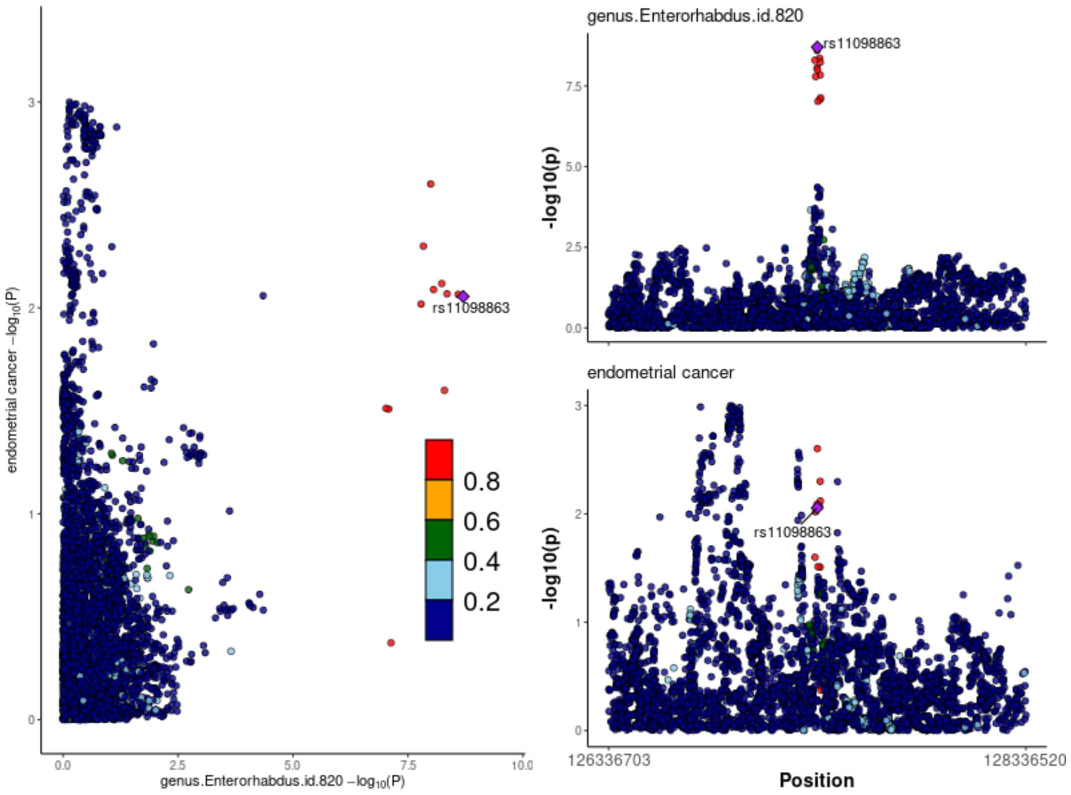


*GWAS = genome-wide association study; SNP = single nucleotide polymorphism. Regional association plots, generated from code available on GitHub(49), showing the −log10(P-value) where the lead SNP (rs11098863) associated with the relative abundance of bacteria in the genus Enterorhabdus (genus.Enterorhabdus.id.820) is represented by a purple diamond. These plots were created using the MiBioGen consortium and the O'Mara et al. (2018) (accessed by IEU Open GWAS using ID: ebi-a-GCST006464) full summary-level GWAS data for microbial traits and endometrial cancer, respectively.*

**Supplementary Figure 24: MR results for effect of G.unclassified.P.Firmicutes.HB on ovarian cancer**

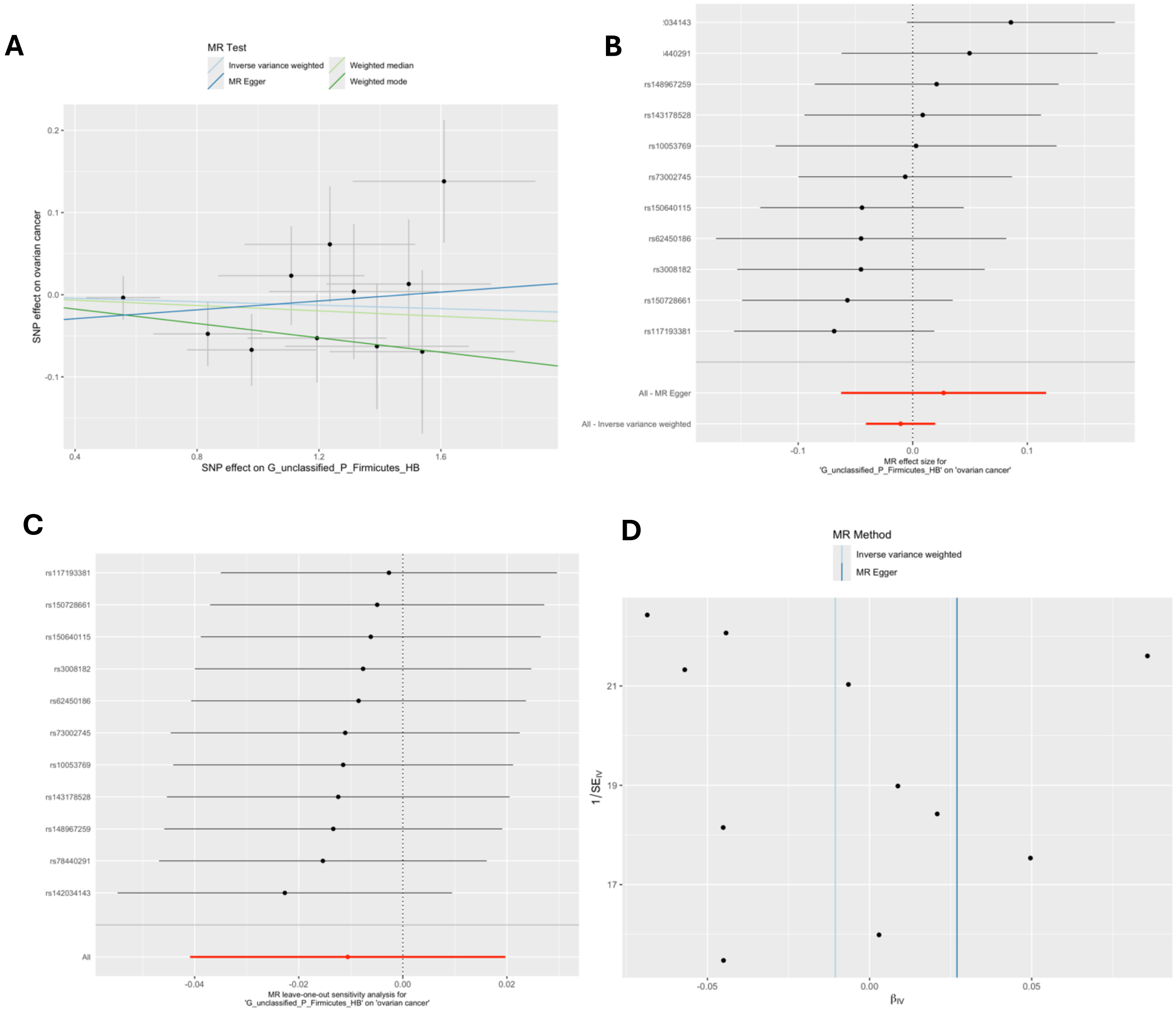


*G = genus; HB = hurdle binary; MR = Mendelian randomization; P = phylum; RNT = reverse normal transformed; SNP = single nucleotide polymorphism. These plots show the results of MR analyses to test the causal effect of presence (vs. absence) of bacteria within the genus of unclassified Firmicutes phylum (G.unclassified.P.Firmicutes.HB) on ovarian cancer using all directionally consistent SNPs associated with G.unclassified.P.Firmicutes.HB at a lenient P-value threshold ($P<1x10^{-5}$) in the Hughes et al. mGWAS(1). A) Scatter plot comparing four MR methods; B) forest plot comparing the effect of individual SNP-level effect estimates derived using the Wald ratio method and in combination with the inverse variance weighted (IVW) and MR-Egger estimates; C) leave-one-out analysis to check if any one SNP is driving pleiotropy or asymmetry in the estimate; and D) funnel plot comparing the effect estimate and precision of each SNP-level Wald ratio estimate.*

**Supplementary Figure 25: MR results for effect of G.unclassified.F.Porphyromonadaceae.RNT on prostate cancer**

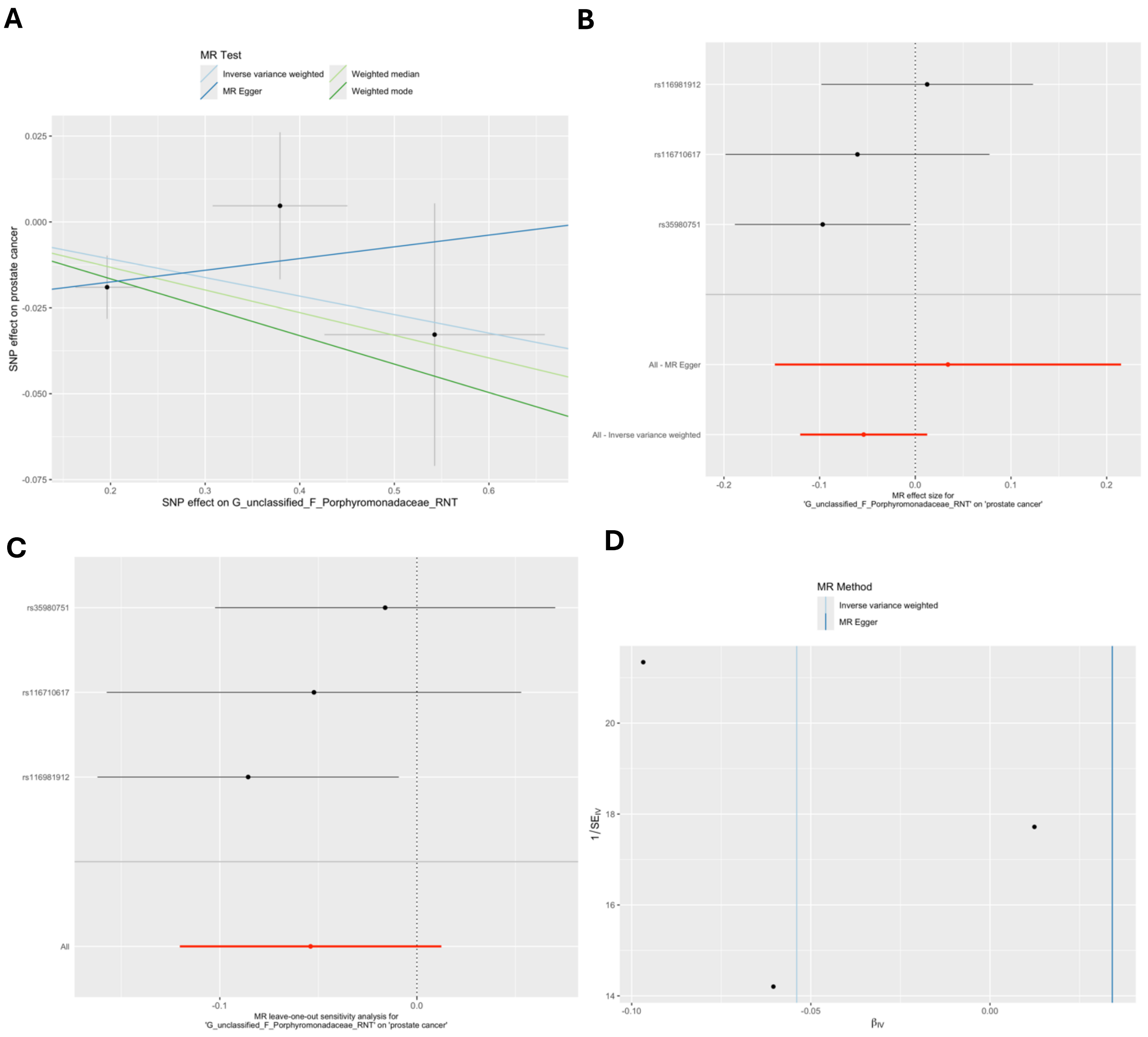


*MR = Mendelian randomization; SNP = single nucleotide polymorphism; G = genus; RNT = reverse normal transformed. These plots show the results of MR analyses to test the causal effect of a higher relative abundance of bacteria within the genus of unclassified family* Porphyromonadaceae (G.unclassified.F.Porphyromonadaceae.RNT*) on prostate cancer using all directionally consistent SNPs associated with* G.unclassified.F.Porphyromonadaceae.RNT*) at a lenient P-value threshold ($P<1x10^{-5}$) in the Hughes et al. mGWAS(1). A) Scatter plot comparing four MR methods; B) forest plot comparing the effect of individual SNP-level effect estimates derived using the Wald ratio method and in combination with the inverse variance weighted (IVW) and MR-Egger estimates; C) leave-one-out analysis to check if any one SNP is driving pleiotropy or asymmetry in the estimate; and D) funnel plot comparing the effect estimate and precision of each SNP-level Wald ratio estimate.*

### Supplementary Figure 26: MR results for effect of family.Oxalobacteraceae.id.2966 on breast cancer

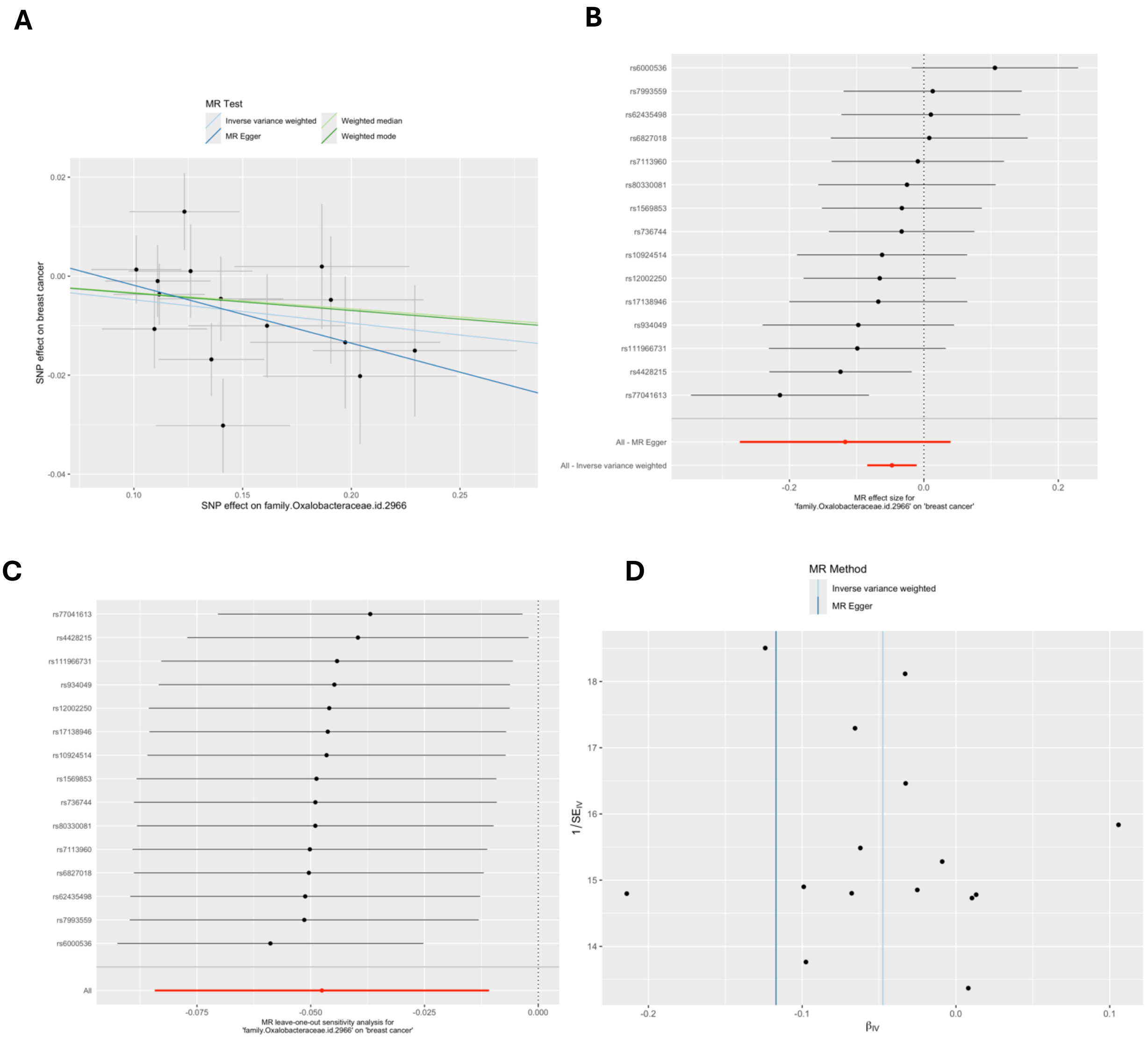


*MR = Mendelian randomization; SNP = single nucleotide polymorphism. These plots show the results of MR analyses to test the causal effect of a higher relative abundance of bacteria within the Oxalobacteraceae family (family.Oxalobacteraceae.id.2966) on breast cancer using all SNPs associated with family.Oxalobacteraceae.id.2966 at a lenient P-value threshold ($P<1x10^{-5}$) in the European-only cohorts in the MiBioGen consortium. A) Scatter plot comparing four MR methods; B) forest plot comparing the effect of individual SNP-level effect estimates derived using the Wald ratio method and in combination with the inverse variance weighted (IVW) and MR-Egger estimates; C) leave-one-out analysis to check if any one SNP is driving pleiotropy or asymmetry in the estimate; and D) funnel plot comparing the effect estimate and precision of each SNP-level Wald ratio estimate.*

**Supplementary Figure 27: MR results for effect of class.Actinobacteria.id.419 on colorectal cancer**

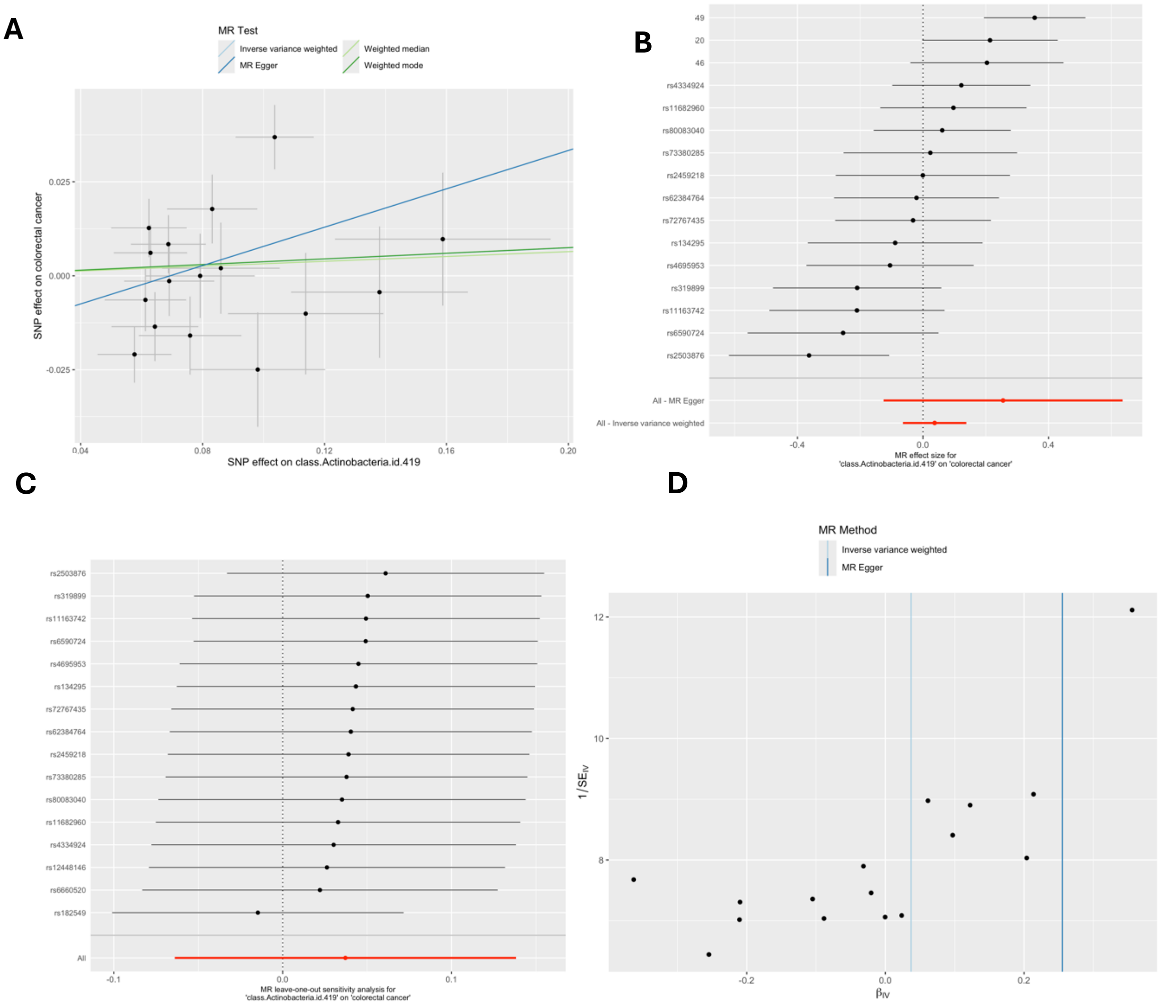


*MR = Mendelian randomization; SNP = single nucleotide polymorphism. These plots show the results of MR analyses to test the causal effect of a higher relative abundance of bacteria within* class Actinobacteria *(class.Actinobacteria.id.419) on colorectal cancer using all SNPs associated with class.Actinobacteria.id.419 at a lenient P-value threshold (P<1x10$^{-5}$) in the European-only cohorts in the MiBioGen consortium. A) Scatter plot comparing four MR methods; B) forest plot comparing the effect of individual SNP-level effect estimates derived using the Wald ratio method and in combination with the inverse variance weighted (IVW) and MR-Egger estimates; C) leave-one-out analysis to check if any one SNP is driving pleiotropy or asymmetry in the estimate; and D) funnel plot comparing the effect estimate and precision of each SNP-level Wald ratio estimate.*

### Supplementary Figure 28: MR results for effect of genus.Erysipelatoclostridium.id.11381 on pancreatic cancer

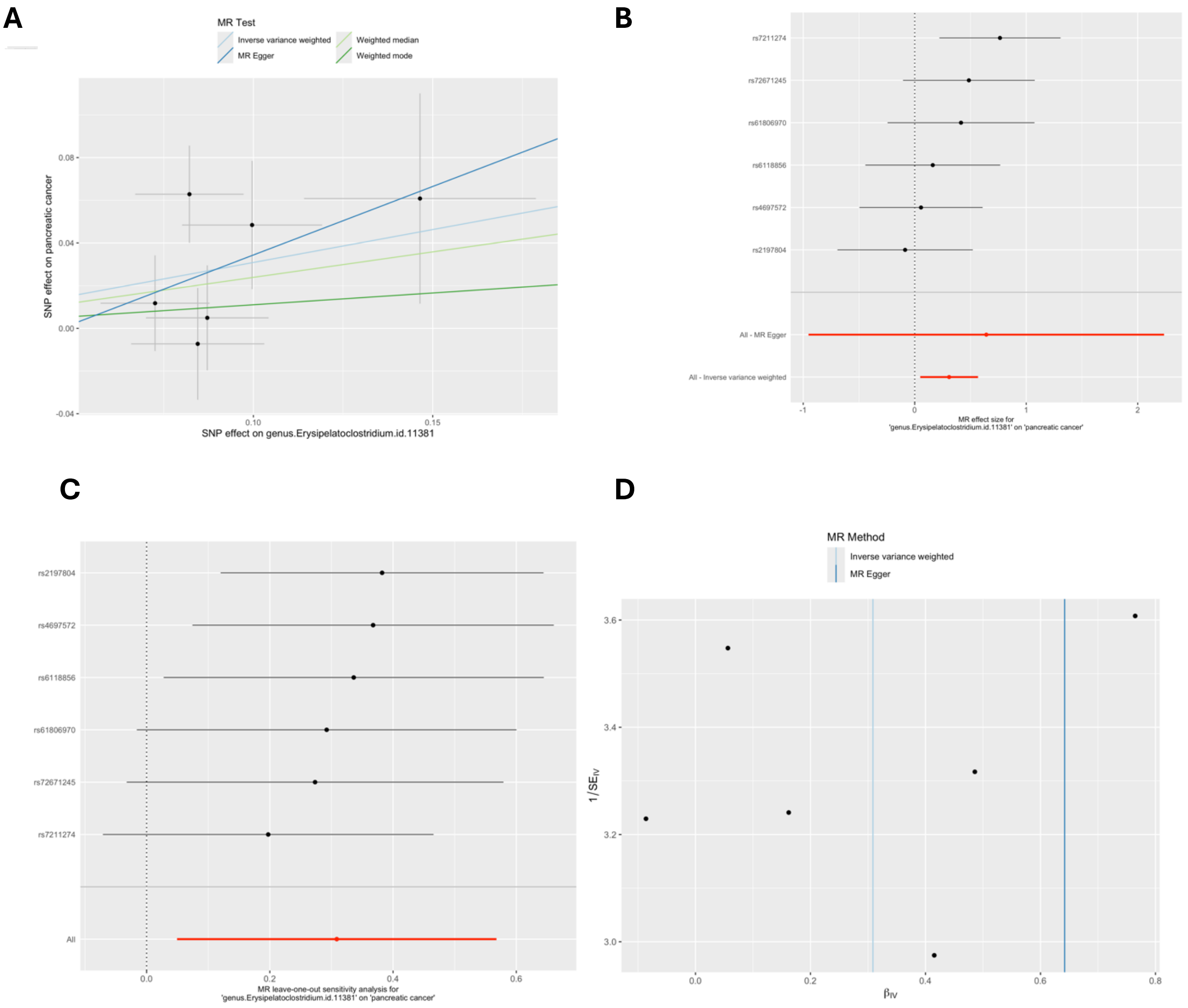


*MR = Mendelian randomization; SNP = single nucleotide polymorphism. These plots show the results of MR analyses to test the causal effect of a higher relative abundance of bacteria within* the genus *Erysipelatoclostridium (genus.Erysipelatoclostridium.id.11381) on pancreatic cancer using all SNPs associated with genus.Erysipelatoclostridium.id.11381 at a lenient P-value threshold ($P<1x10^{-5}$) in the European-only cohorts in the MiBioGen consortium. A) Scatter plot comparing four MR methods; B) forest plot comparing the effect of individual SNP-level effect estimates derived using the Wald ratio method and in combination with the inverse variance weighted (IVW) and MR-Egger estimates; C) leave-one-out analysis to check if any one SNP is driving pleiotropy or asymmetry in the estimate; and D) funnel plot comparing the effect estimate and precision of each SNP-level Wald ratio estimate.*

### Supplementary Figure 29: MR results for effect of lung cancer on genus.Intestinibacter.id.11345

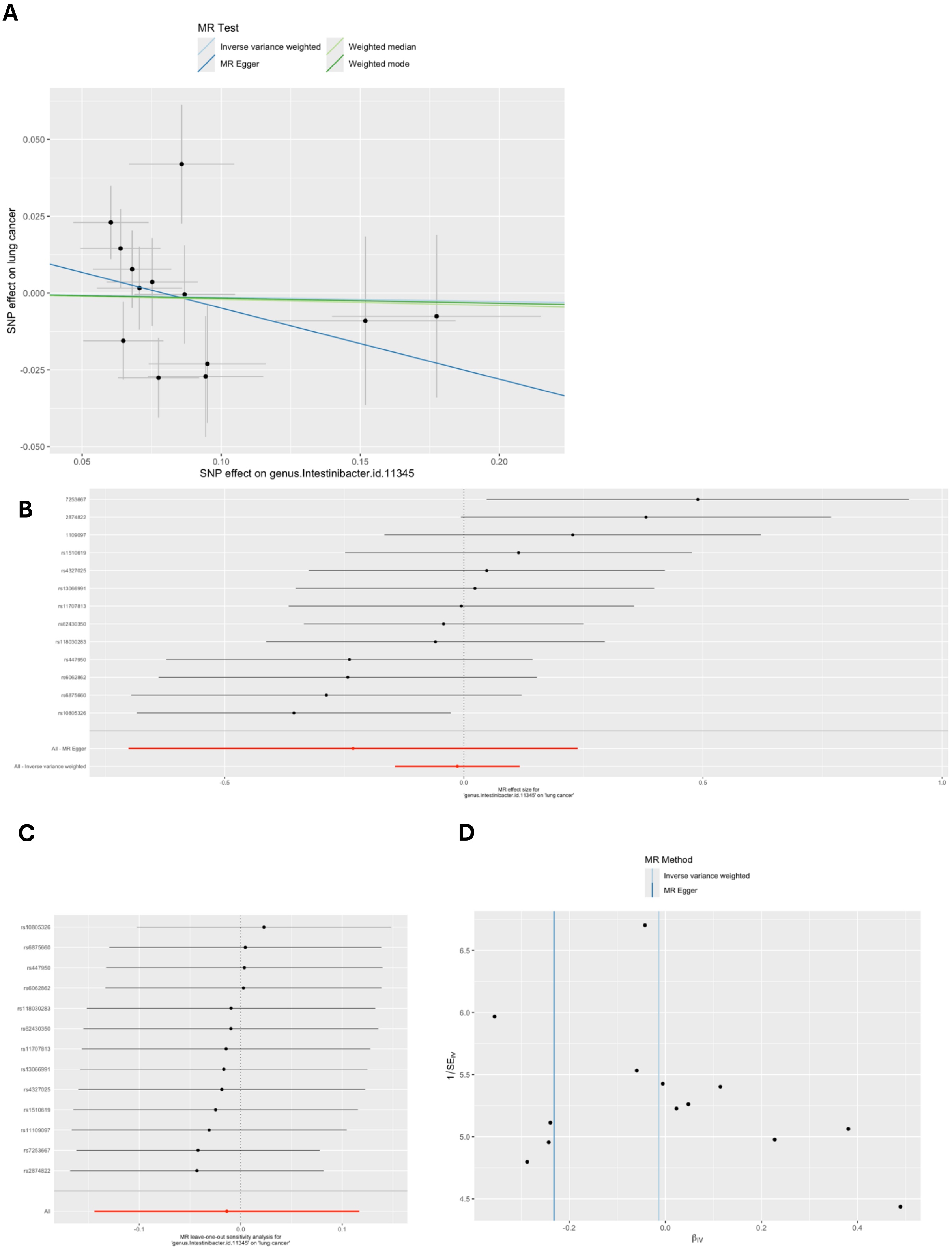


*MR = Mendelian randomization; SNP = single nucleotide polymorphism. These plots show the results of MR analyses to test the causal effect of a higher relative abundance of bacteria within* the genus

Intestinibacter *(genus.Intestinibacter.id.11345) on lung cancer using all SNPs associated with genus.Intestinibacter.id.11345 at a lenient P-value threshold ($P<1x10^{-5}$) in the European-only cohorts in the MiBioGen consortium. A) Scatter plot comparing four MR methods; B) forest plot comparing the effect of individual SNP-level effect estimates derived using the Wald ratio method and in combination with the inverse variance weighted (IVW) and MR-Egger estimates; C) leave-one-out analysis to check if any one SNP is driving pleiotropy or asymmetry in the estimate; and D) funnel plot comparing the effect estimate and precision of each SNP-level Wald ratio estimate.*

**Supplementary Figure 30: MR results for effect of lung cancer on genus.Streptococcus.id.1853**

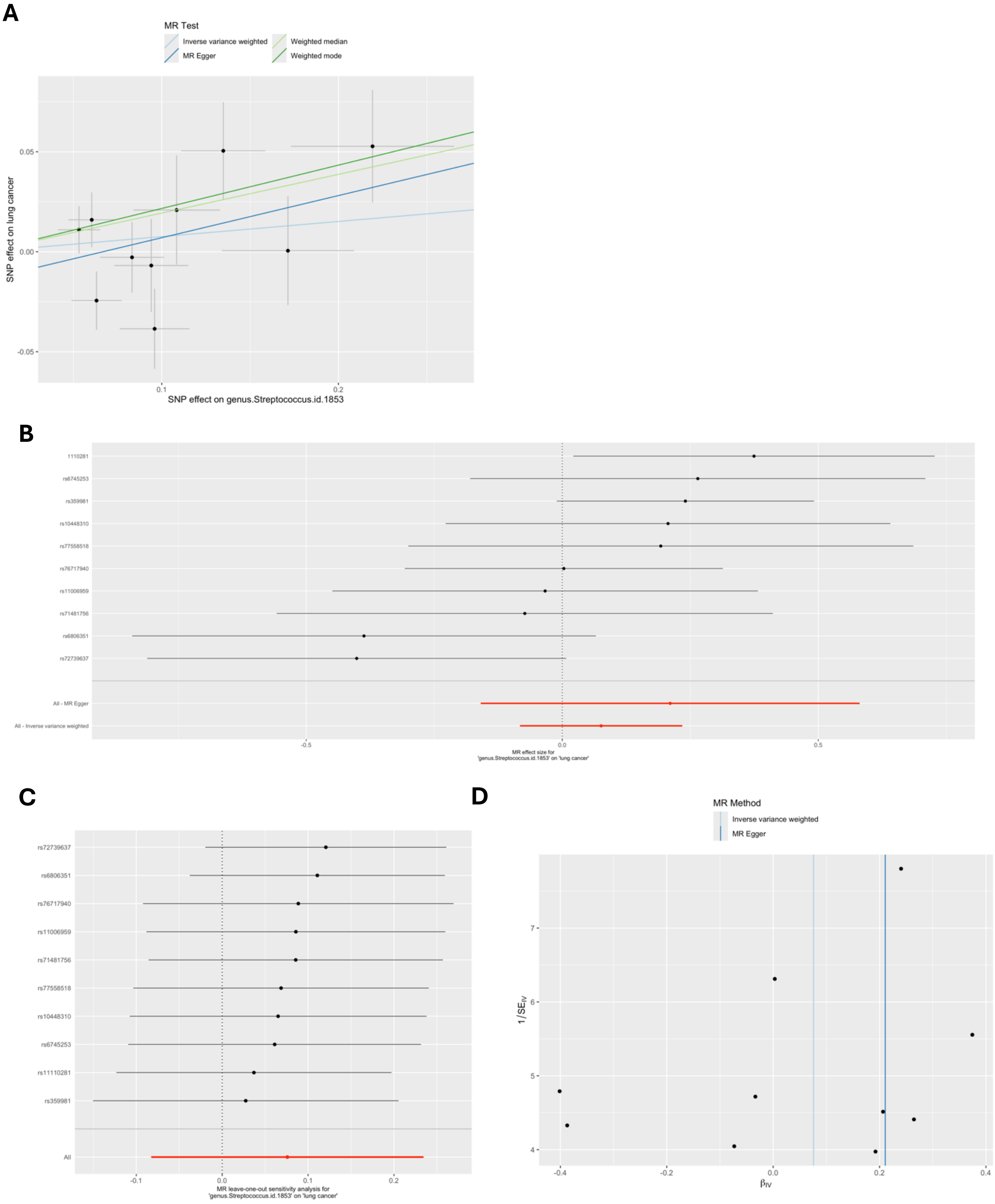


*MR = Mendelian randomization; SNP = single nucleotide polymorphism. These plots show the results of MR analyses to test the causal effect of a higher relative abundance of bacteria within* the genus *Streptococcus (genus.Streptococcus.id.1853) on lung cancer using all SNPs associated with*

*genus.Streptococcus.id.1853 at a lenient P-value threshold ($P<1x10^{-5}$) in the European-only cohorts in the MiBioGen consortium. A) Scatter plot comparing four MR methods; B) forest plot comparing the effect of individual SNP-level effect estimates derived using the Wald ratio method and in combination with the inverse variance weighted (IVW) and MR-Egger estimates; C) leave-one-out analysis to check if any one SNP is driving pleiotropy or asymmetry in the estimate; and D) funnel plot comparing the effect estimate and precision of each SNP-level Wald ratio estimate.*

**Supplementary Figure 31: MR results for effect of Ruminococcustorquesgroup.id.14377 on colorectal cancer**

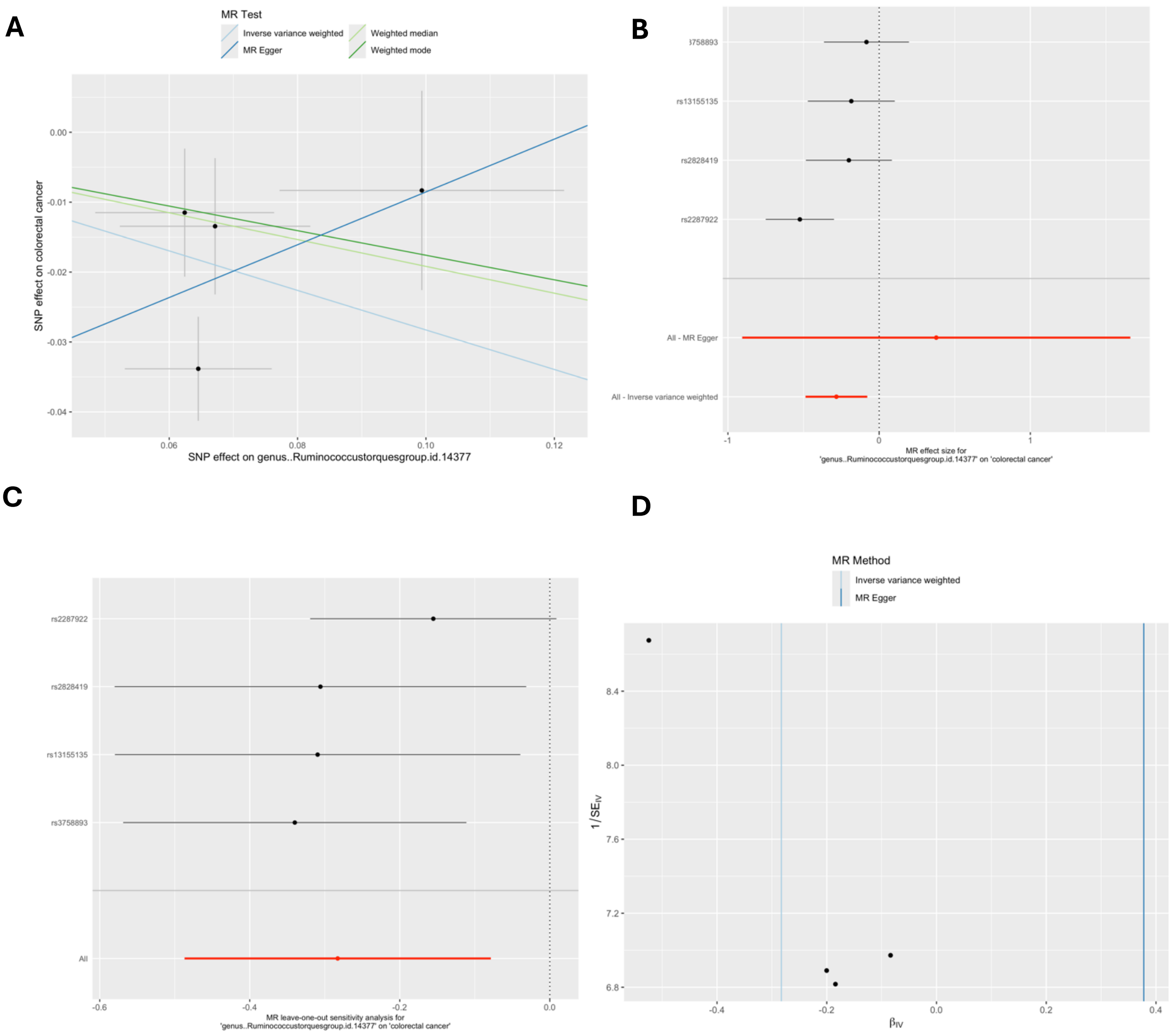


*MR = Mendelian randomization; SNP = single nucleotide polymorphism. These plots show the results of MR analyses to test the causal effect of a higher relative abundance of bacteria within* Ruminococcus torques group (Ruminococcustorquesgroup.id.14377*) on colorectal cancer using all SNPs associated with* Ruminococcustorquesgroup.id.14377 *at a lenient P-value threshold (P<1x10$^{-5}$) in the European-only cohorts in the MiBioGen consortium. A) Scatter plot comparing four MR methods; B) forest plot comparing the effect of individual SNP-level effect estimates derived using the Wald ratio method and in combination with the inverse variance weighted (IVW) and MR-Egger estimates; C) leave-one-out analysis to check if any one SNP is driving pleiotropy or asymmetry in the estimate; and D) funnel plot comparing the effect estimate and precision of each SNP-level Wald ratio estimate.*

**Supplementary Figure 32: MR results for effect of family.Oxalobacteraceae.id.2966 on colorectal cancer**

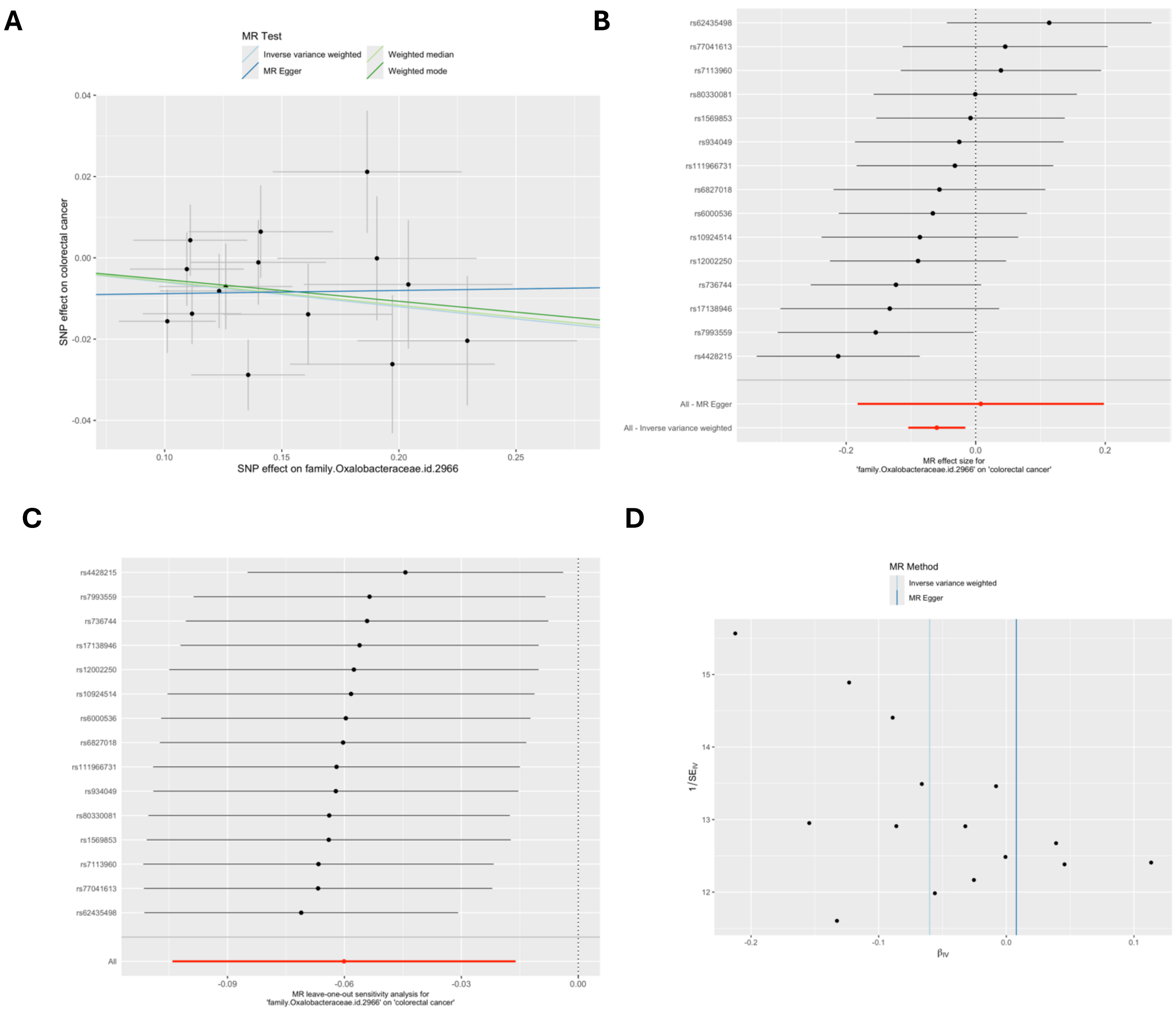


*MR = Mendelian randomization; SNP = single nucleotide polymorphism. These plots show the results of MR analyses to test the causal effect of a higher relative abundance of bacteria within the Oxalobacteraceae family* (family.Oxalobacteraceae.id.2966) on colorectal cancer using all SNPs associated with family.Oxalobacteraceae.id.2966 *at a lenient P-value threshold (P<1x10$^{-5}$) in the European-only cohorts in the MiBioGen consortium. A) Scatter plot comparing four MR methods; B) forest plot comparing the effect of individual SNP-level effect estimates derived using the Wald ratio method and in combination with the inverse variance weighted (IVW) and MR-Egger estimates; C) leave-one-out analysis to check if any one SNP is driving pleiotropy or asymmetry in the estimate; and D) funnel plot comparing the effect estimate and precision of each SNP-level Wald ratio estimate.*

### Supplementary Figure 33: MR results for effect of phylum.Actinobacteria.id.400 on colorectal cancer

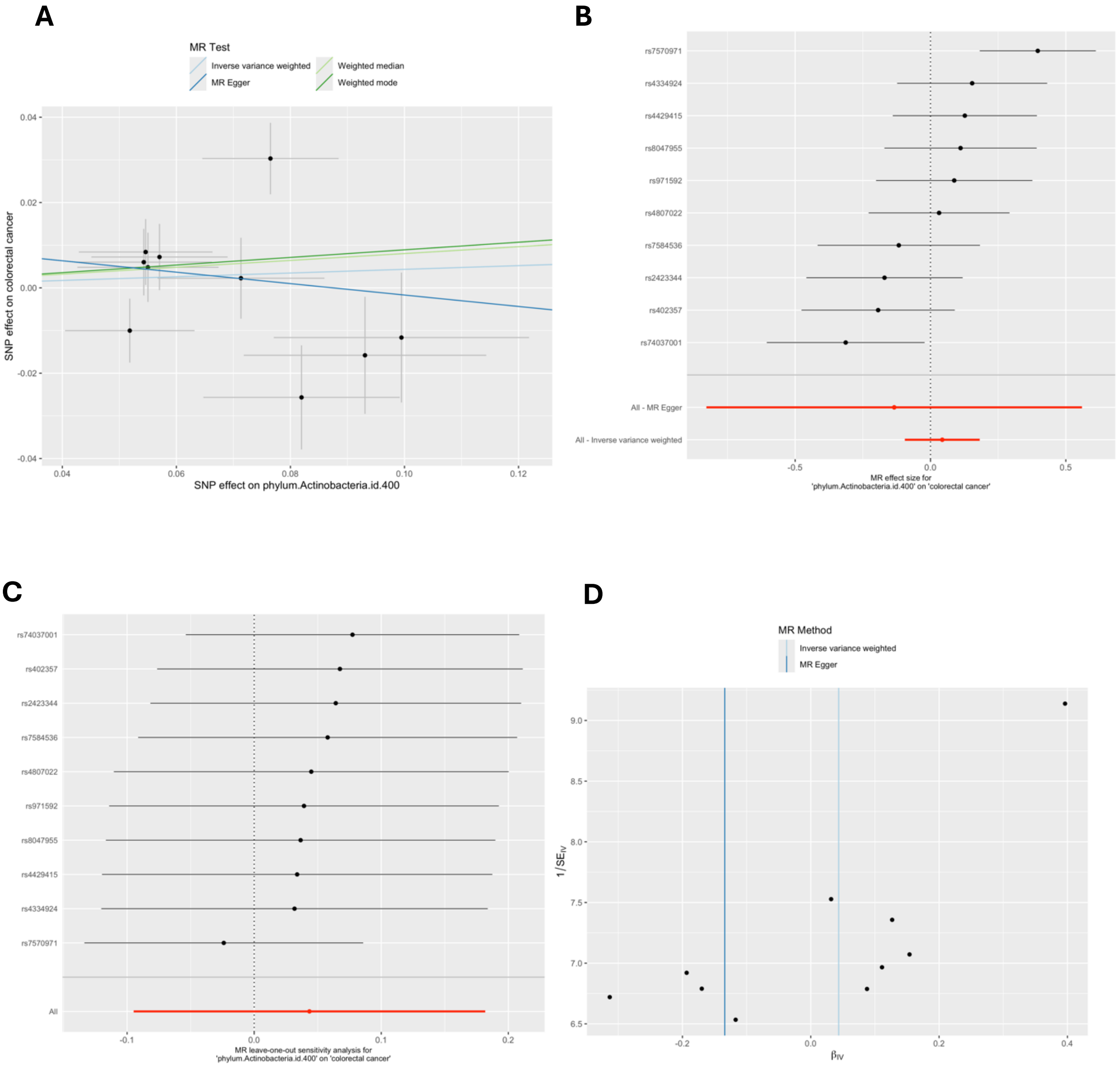


*MR = Mendelian randomization; SNP = single nucleotide polymorphism. These plots show the results of MR analyses to test the causal effect of a higher relative abundance of bacteria within the Actinobacteria phylum (phylum.Actinobacteria.id.400) on colorectal cancer using all SNPs associated with phylum.Actinobacteria.id.400 at a lenient P-value threshold ($P<1x10^{-5}$) in the European-only cohorts in the MiBioGen consortium. A) Scatter plot comparing four MR methods; B) forest plot comparing the effect of individual SNP-level effect estimates derived using the Wald ratio method and in combination with the inverse variance weighted (IVW) and MR-Egger estimates; C) leave-one-out analysis to check if any one SNP is driving pleiotropy or asymmetry in the estimate; and D) funnel plot comparing the effect estimate and precision of each SNP-level Wald ratio estimate.*

### Supplementary Figure 34: MR results for effect of family.Streptococcaceae.id.1850 on lung cancer

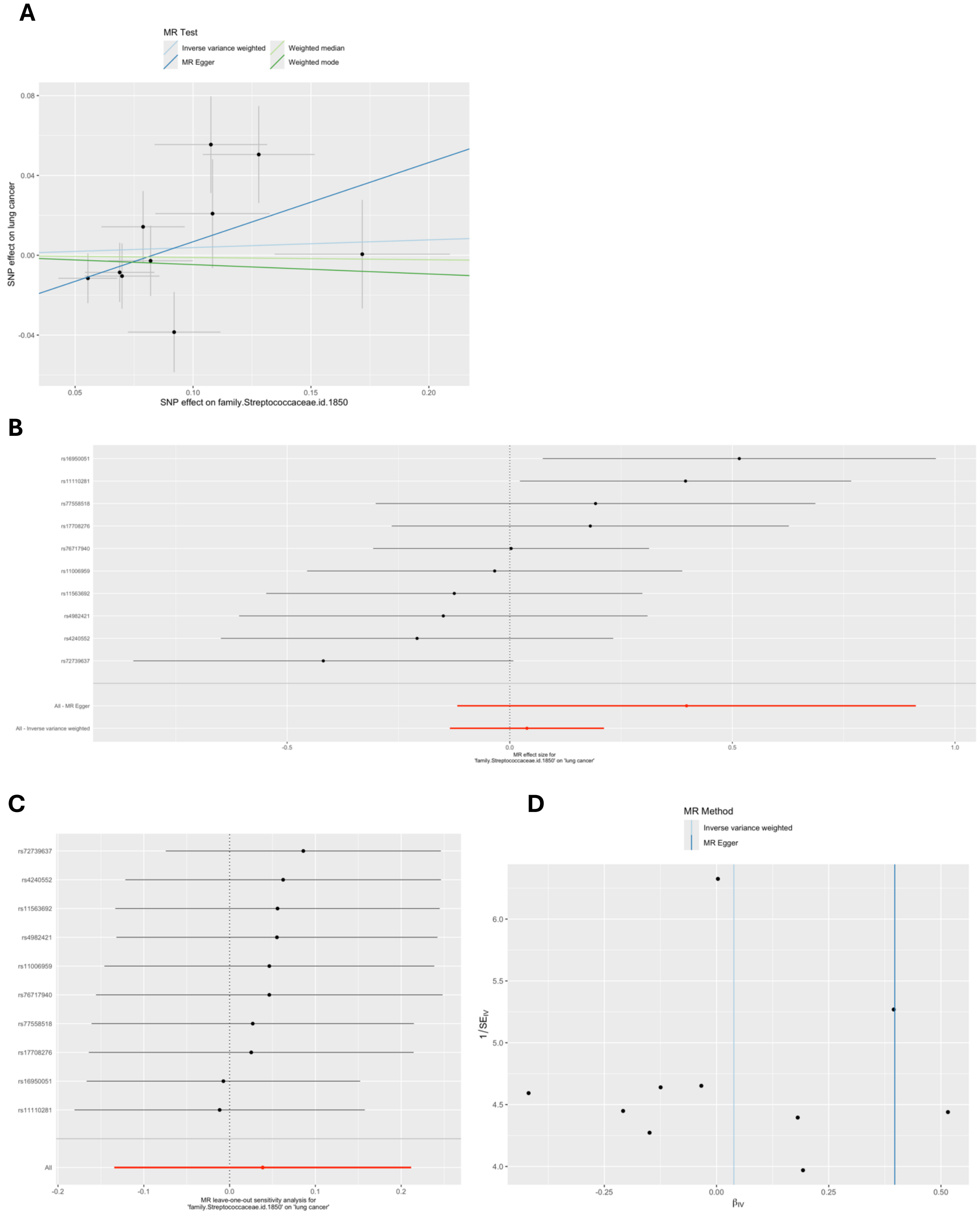

*MR = Mendelian randomization; SNP = single nucleotide polymorphism. These plots show the results of MR analyses to test the causal effect of a higher relative abundance of bacteria within the Streptococcaceae family (family.Streptococcaceae.id.1850) on lung cancer using all SNPs associated with family.Streptococcaceae.id.1850 at a lenient P-value threshold ($P<1x10^{-5}$) in the European-only cohorts in the MiBioGen consortium. A) Scatter plot comparing four MR methods; B) forest plot comparing the effect of individual SNP-level effect estimates derived using the Wald ratio method and in combination with the inverse variance weighted (IVW) and MR-Egger estimates; C) leave-one-out analysis to check if any one SNP is driving pleiotropy or asymmetry in the estimate; and D) funnel plot comparing the effect estimate and precision of each SNP-level Wald ratio estimate.*

### Supplementary Figure 35: MR results for effect of genus.Romboutsia.id.11347 on pancreatic cancer

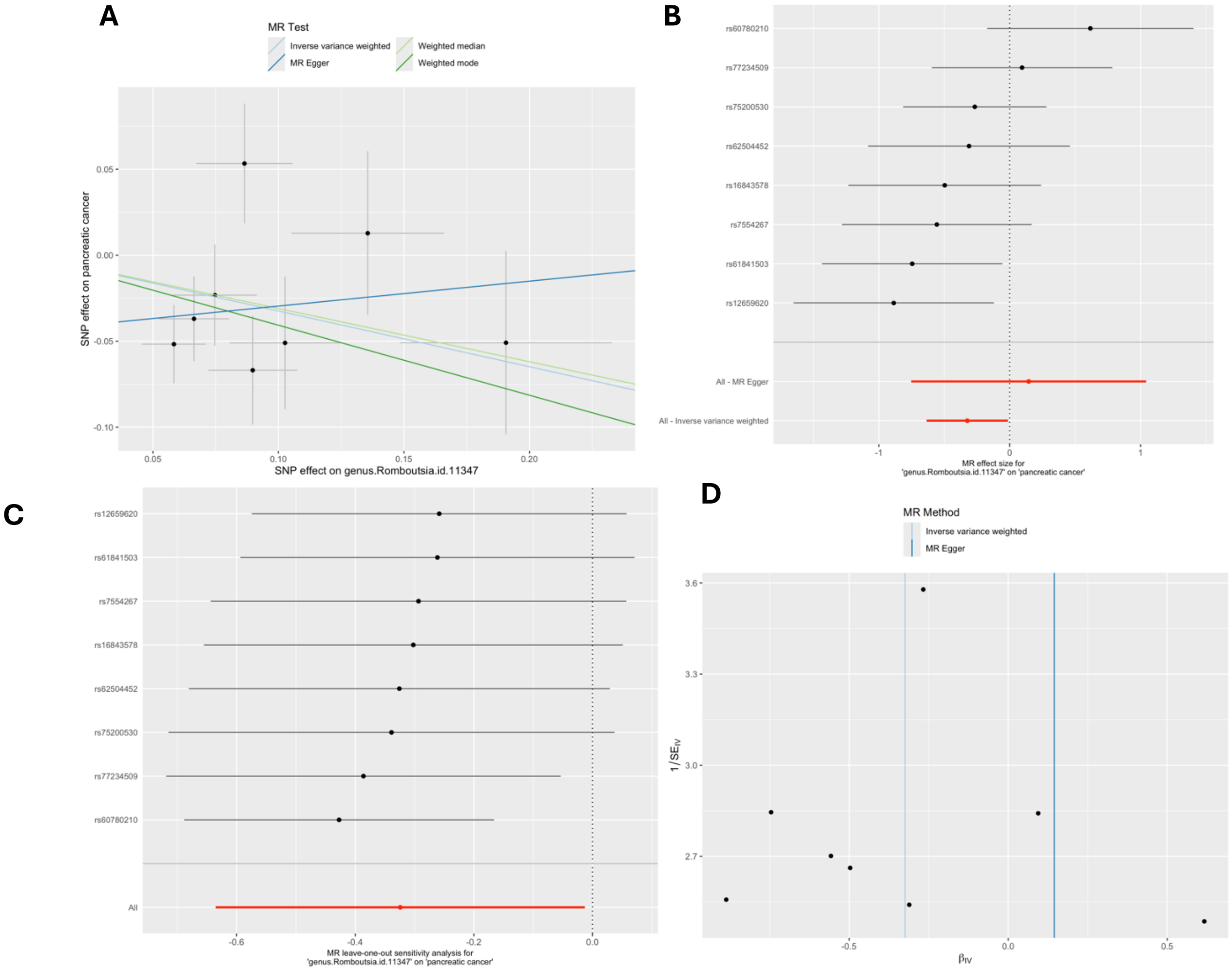


*MR = Mendelian randomization; SNP = single nucleotide polymorphism. These plots show the results of MR analyses to test the causal effect of a higher relative abundance of bacteria within* the genus *Romboutsia (*genus.Romboutsia.id.11347*) on pancreatic cancer using all SNPs associated with* genus.Romboutsia.id.11347 *at a lenient P-value threshold (P<1x10$^{-5}$) in the European-only cohorts in the MiBioGen consortium. A) Scatter plot comparing four MR methods; B) forest plot comparing the effect of individual SNP-level effect estimates derived using the Wald ratio method and in combination with the inverse variance weighted (IVW) and MR-Egger estimates; C) leave-one-out analysis to check if any one SNP is driving pleiotropy or asymmetry in the estimate; and D) funnel plot comparing the effect estimate and precision of each SNP-level Wald ratio estimate.*

### Supplementary Figure 36: MR results for effect of family.Peptostreptococcaceae.id.2042 on pancreatic cancer

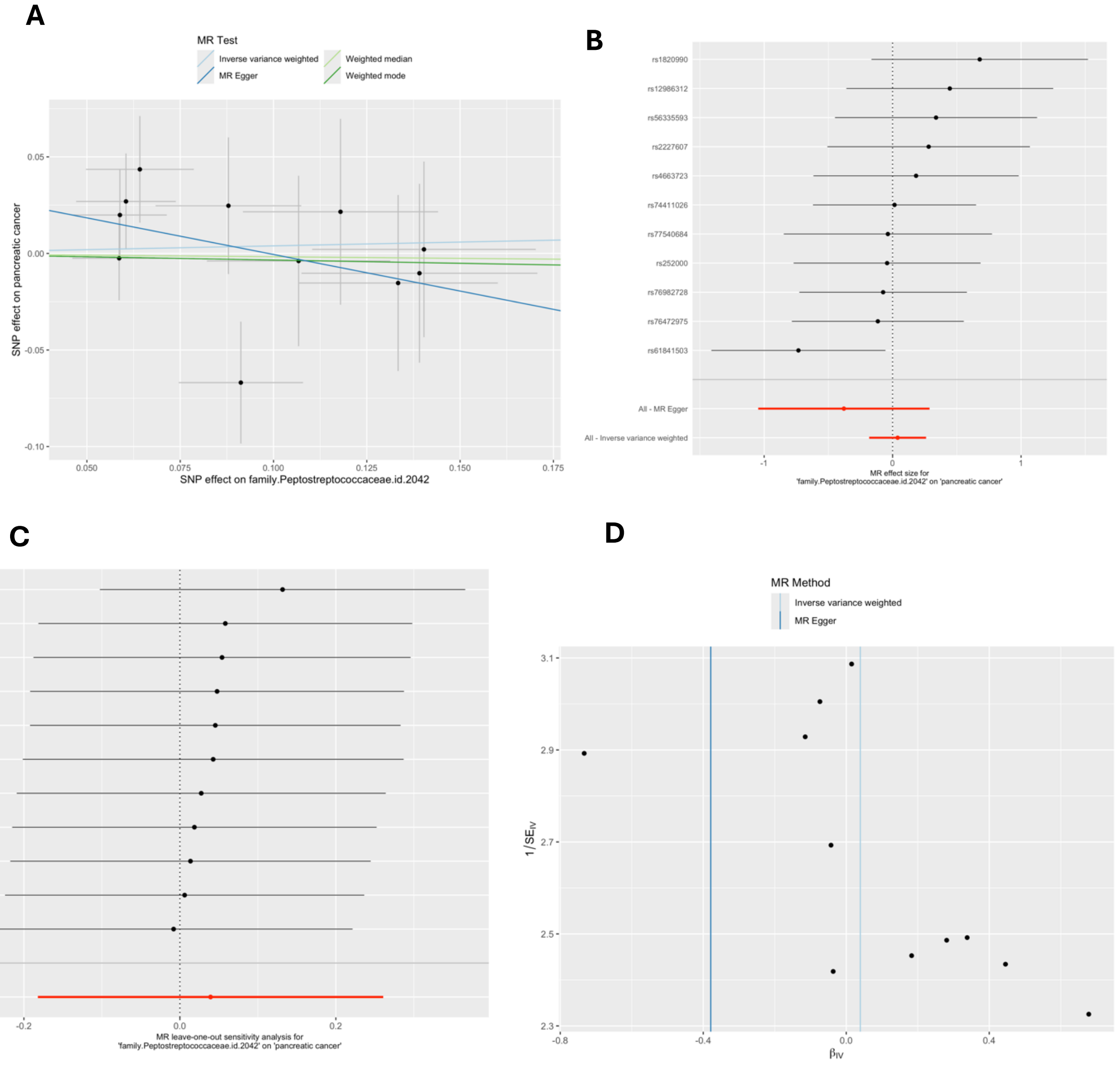


*MR = Mendelian randomization; SNP = single nucleotide polymorphism. These plots show the results of MR analyses to test the causal effect of a higher relative abundance of bacteria within* the family Peptostreptococcaceae (family.Peptostreptococcaceae.id.2042*) on pancreatic cancer using all SNPs associated with* family.Peptostreptococcaceae.id.20422174 *at a lenient P-value threshold ($P<1x10^{-5}$) in the European-only cohorts in the MiBioGen consortium. A) Scatter plot comparing four MR methods; B) forest plot comparing the effect of individual SNP-level effect estimates derived using the Wald ratio method and in combination with the inverse variance weighted (IVW) and MR-Egger estimates; C) leave-one-out analysis to check if any one SNP is driving pleiotropy or asymmetry in the estimate; and D) funnel plot comparing the effect estimate and precision of each SNP-level Wald ratio estimate.*

**Supplementary Figure 37: MR results for effect of family.Oxalobacteraceae.id.2966 on endometrial cancer**

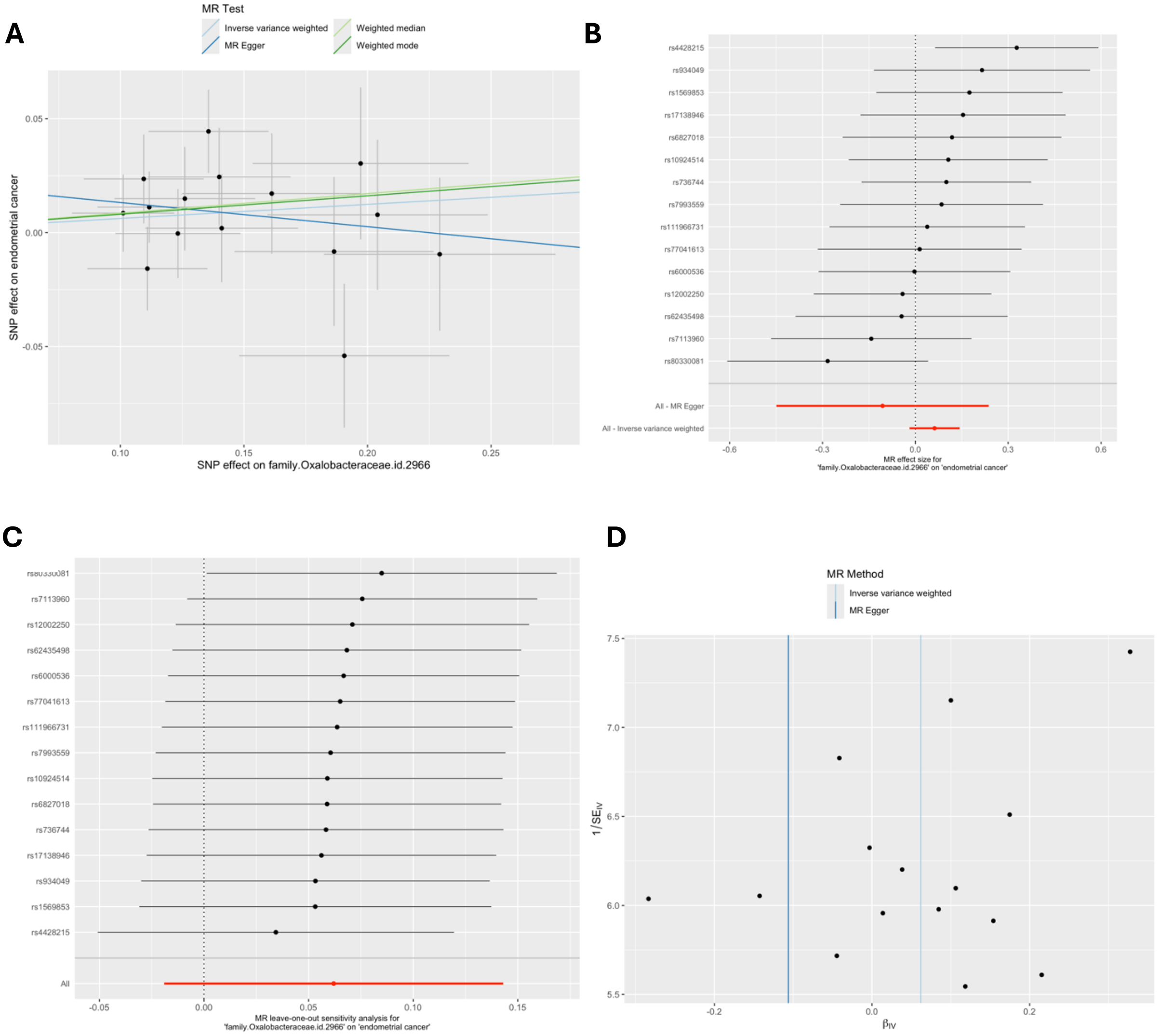


*MR = Mendelian randomization; SNP = single nucleotide polymorphism. These plots show the results of MR analyses to test the causal effect of a higher relative abundance of bacteria within the family* Oxalobacteraceae (family.Oxalobacteraceae.id.2966*) on endometrial cancer using all SNPs associated with* family.Oxalobacteraceae.id.2966 *at a lenient P-value threshold (P<1x10$^{-5}$) in the European-only cohorts in the MiBioGen consortium. A) Scatter plot comparing four MR methods; B) forest plot comparing the effect of individual SNP-level effect estimates derived using the Wald ratio method and in combination with the inverse variance weighted (IVW) and MR-Egger estimates; C) leave-one-out analysis to check if any one SNP is driving pleiotropy or asymmetry in the estimate; and D) funnel plot comparing the effect estimate and precision of each SNP-level Wald ratio estimate.*

### Supplementary Figure 38: MR results for effect of genus.Enterorhabdus.id.820 on endometrial cancer

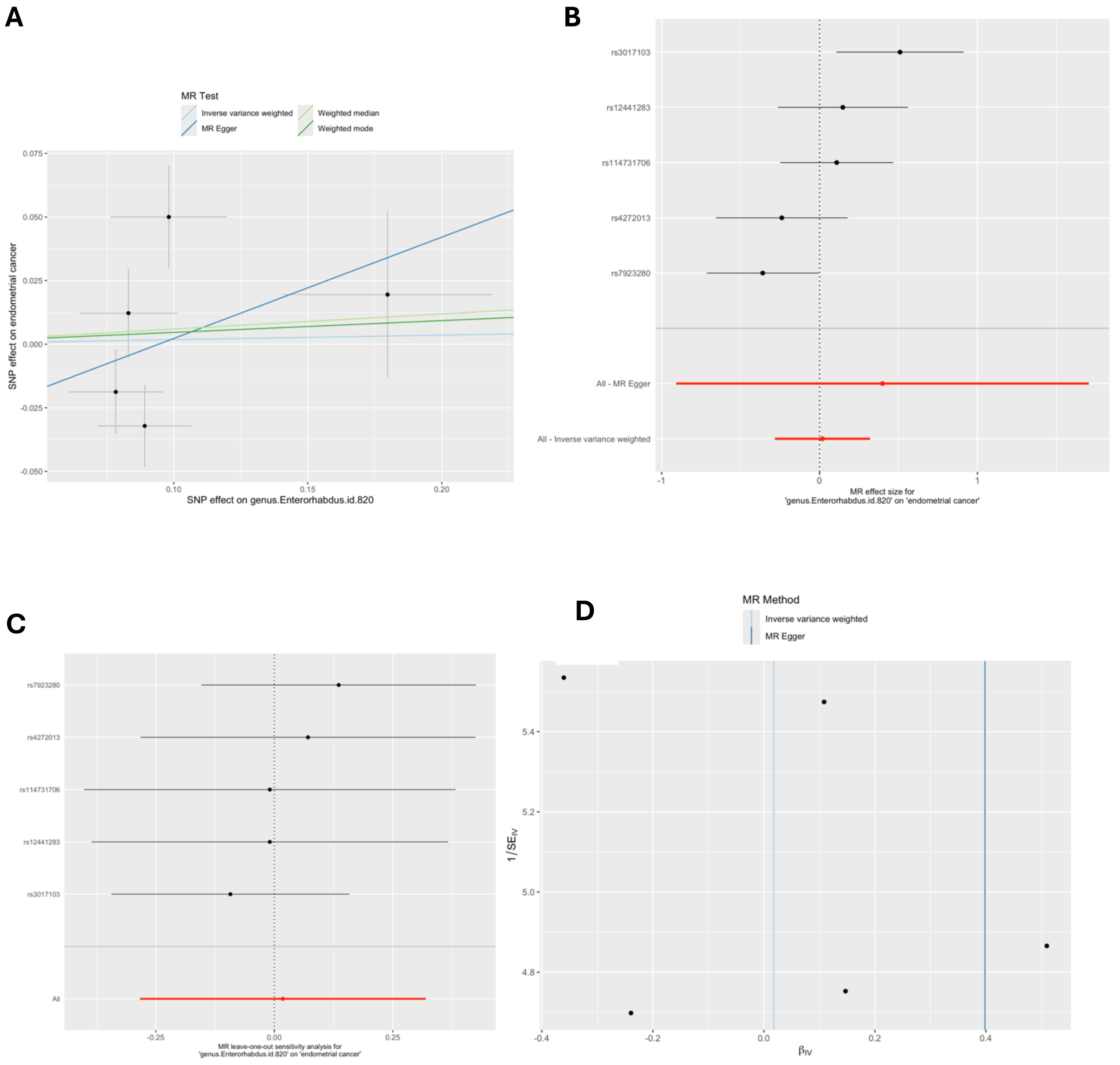


*MR = Mendelian randomization; SNP = single nucleotide polymorphism. These plots show the results of MR analyses to test the causal effect of a higher relative abundance of bacteria within the genus Enterorhabdus (genus.Enterorhabdus.id.820) on endometrial cancer using all SNPs associated with genus.Enterorhabdus.id.820 at a lenient P-value threshold ($P<1x10^{-5}$) in the European-only cohorts in the MiBioGen consortium. A) Scatter plot comparing four MR methods; B) forest plot comparing the effect of individual SNP-level effect estimates derived using the Wald ratio method and in combination with the inverse variance weighted (IVW) and MR-Egger estimates; C) leave-one-out analysis to check if any one SNP is driving pleiotropy or asymmetry in the estimate; and D) funnel plot comparing the effect estimate and precision of each SNP-level Wald ratio estimate.*

### Supplementary Figure 39: MR results for effect of family.Streptococcaceae.id.1850 on endometrial cancer

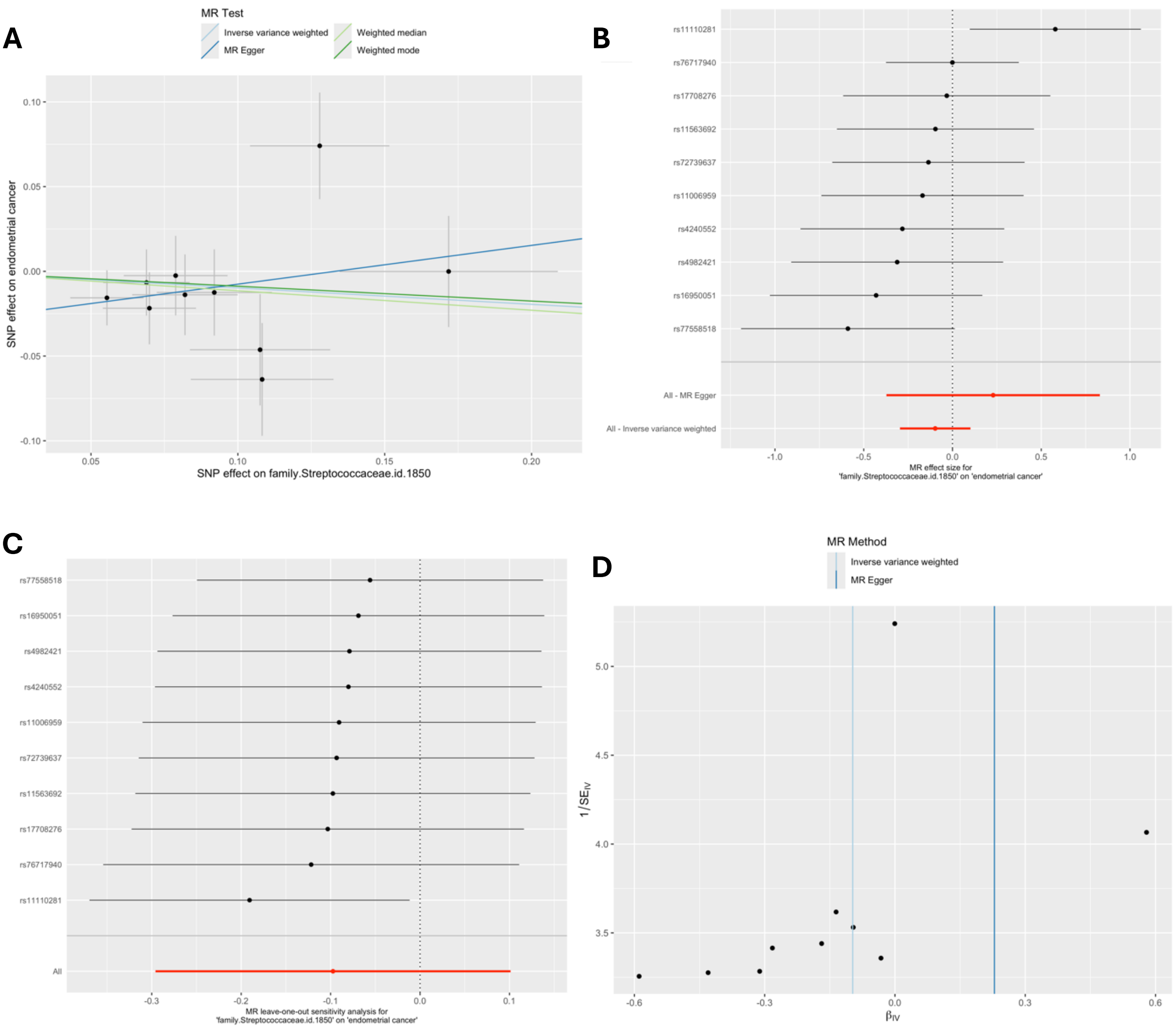


*MR = Mendelian randomization; SNP = single nucleotide polymorphism. These plots show the results of MR analyses to test the causal effect of a higher relative abundance of bacteria within the family* Streptococcaceae (family.Streptococcaceae.id.1850*) on endometrial cancer using all SNPs associated with* family.Streptococcaceae.id.1850 *at a lenient P-value threshold (P<1x10$^{-5}$) in the European-only cohorts in the MiBioGen consortium. A) Scatter plot comparing four MR methods; B) forest plot comparing the effect of individual SNP-level effect estimates derived using the Wald ratio method and in combination with the inverse variance weighted (IVW) and MR-Egger estimates; C) leave-one-out analysis to check if any one SNP is driving pleiotropy or asymmetry in the estimate; and D) funnel plot comparing the effect estimate and precision of each SNP-level Wald ratio estimate.*

**Supplementary Figure 40: MR results for effect of genus.Streptococcus.id.1853 on endometrial cancer**

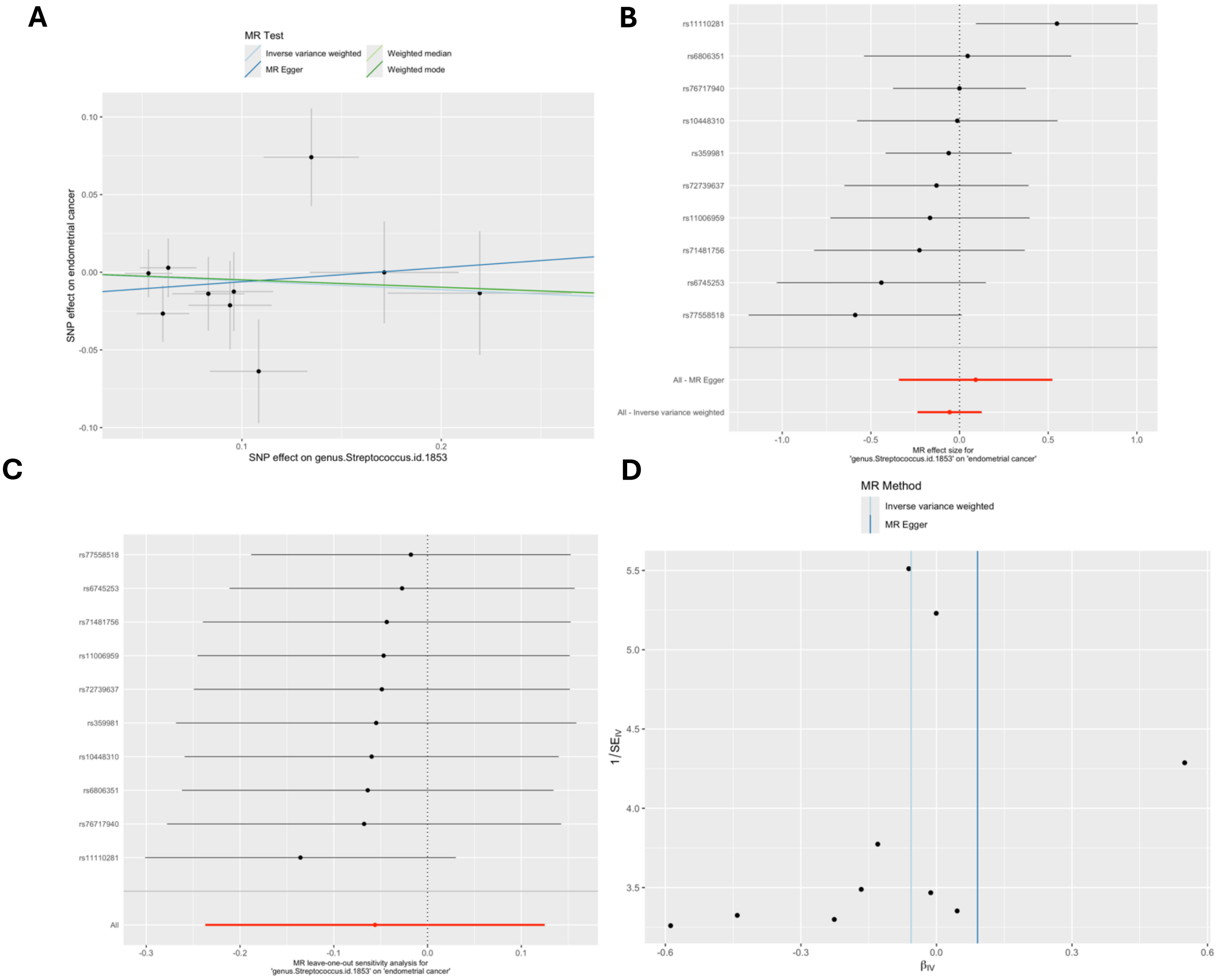


*MR = Mendelian randomization; SNP = single nucleotide polymorphism. These plots show the results of MR analyses to test the causal effect of a higher relative abundance of bacteria within the genus* Streptococcus (genus.Streptococcus.id.1853*) on endometrial cancer using all SNPs associated with* genus.Streptococcus.id.1853 *at a lenient P-value threshold (P<1x10$^{-5}$) in the European-only cohorts in the MiBioGen consortium. A) Scatter plot comparing four MR methods; B) forest plot comparing the effect of individual SNP-level effect estimates derived using the Wald ratio method and in combination with the inverse variance weighted (IVW) and MR-Egger estimates; C) leave-one-out analysis to check if any one SNP is driving pleiotropy or asymmetry in the estimate; and D) funnel plot comparing the effect estimate and precision of each SNP-level Wald ratio estimate.*

**Supplementary Figure 41: MR results for effect of genus.Ruminococcustorquesgroup.id.14377 on oesophageal cancer**

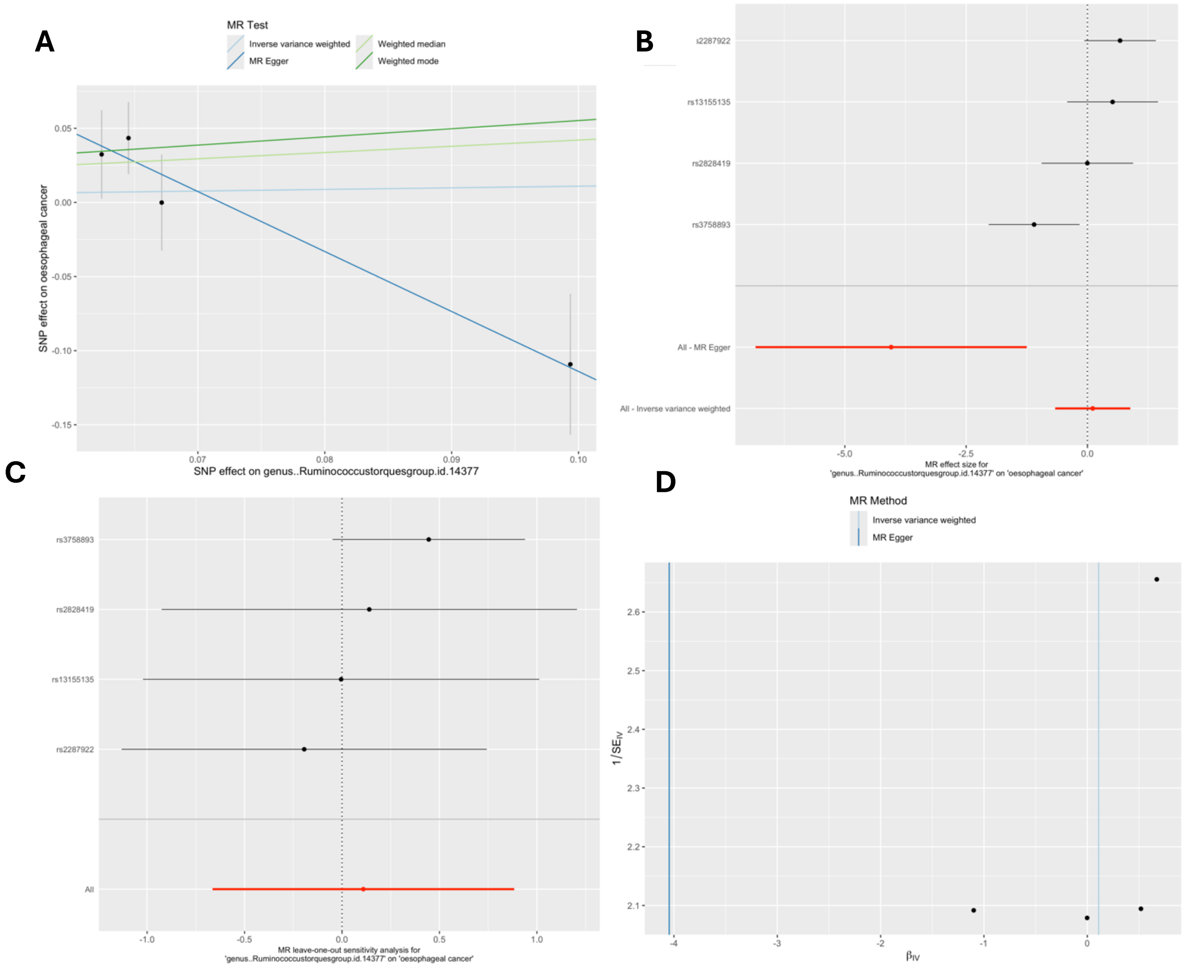


*MR = Mendelian randomization; SNP = single nucleotide polymorphism. These plots show the results of MR analyses to test the causal effect of a higher relative abundance of bacteria within the genus* Streptococcus (genus..Ruminococcustorquesgroup.id.14377*) on oesophageal cancer using all SNPs associated with* genus..Ruminococcustorquesgroup.id.14377 *at a lenient P-value threshold (P<1x10*$^{-5}$*) in the European-only cohorts in the MiBioGen consortium. A) Scatter plot comparing four MR methods; B) forest plot comparing the effect of individual SNP-level effect estimates derived using the Wald ratio method and in combination with the inverse variance weighted (IVW) and MR-Egger estimates; C) leave-one-out analysis to check if any one SNP is driving pleiotropy or asymmetry in the estimate; and D) funnel plot comparing the effect estimate and precision of each SNP-level Wald ratio estimate.*

### Supplementary Figure 42: Reverse MR results for effect of pancreatic cancer on genus.Romboutsia.id.11347

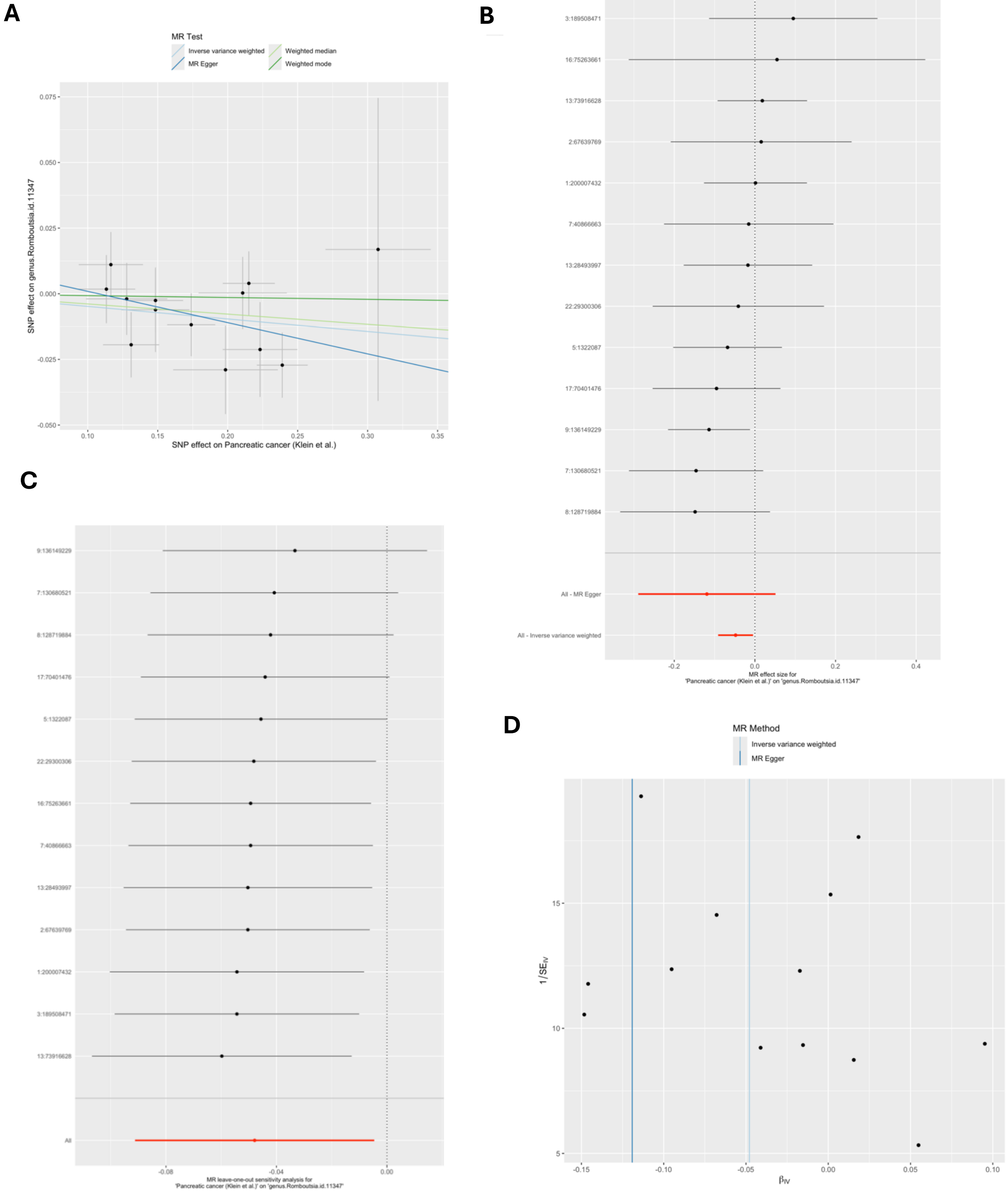


*MR = Mendelian randomization; SNP = single nucleotide polymorphism. These plots show the results of reverse MR analyses to test the causal effect of pancreatic cancer on a higher relative abundance of bacteria within the genus* Romboutsia *(on* genus.Romboutsia.id.11347*) using all SNPs associated with* pancreatic cancer *at a genome-wide significant P-value threshold (P<5x10$^{-8}$) in the Klein et al (2018)*

*pancreatic cancer GWAS. A) Scatter plot comparing four MR methods; B) forest plot comparing the effect of individual SNP-level effect estimates derived using the Wald ratio method and in combination with the inverse variance weighted (IVW) and MR-Egger estimates; C) leave-one-out analysis to check if any one SNP is driving pleiotropy or asymmetry in the estimate; and D) funnel plot comparing the effect estimate and precision of each SNP-level Wald ratio estimate.*

### Supplementary Figure 43: Reverse MR results for effect of pancreatic cancer on family.Peptostreptococcaceae.id.2042

A

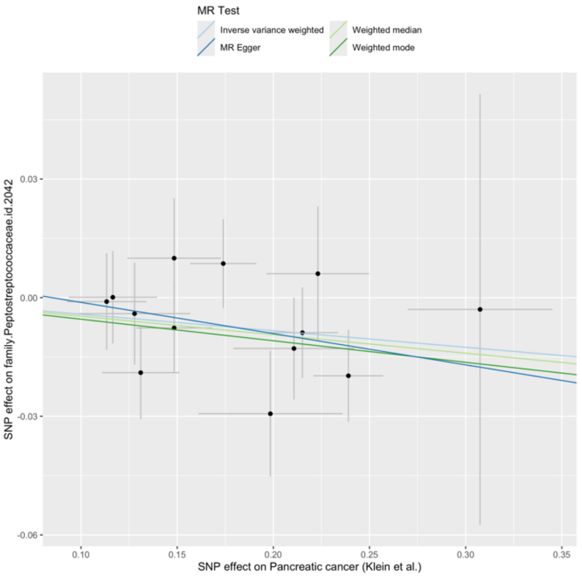


B

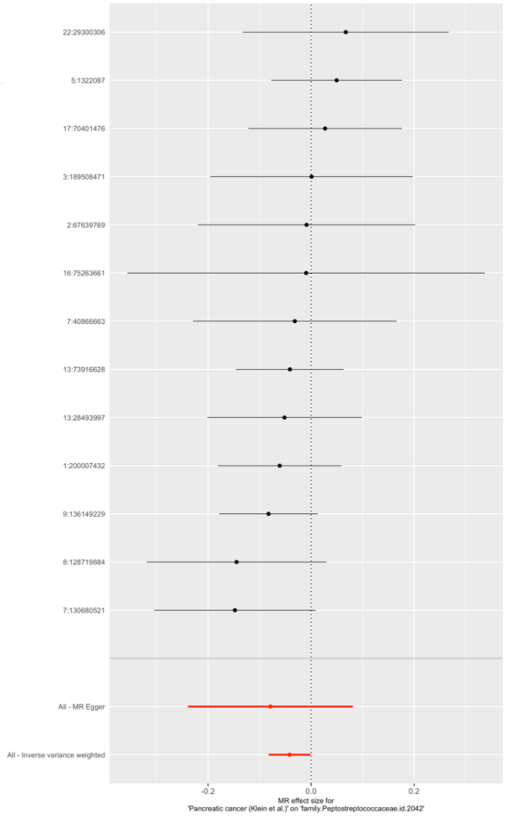


C

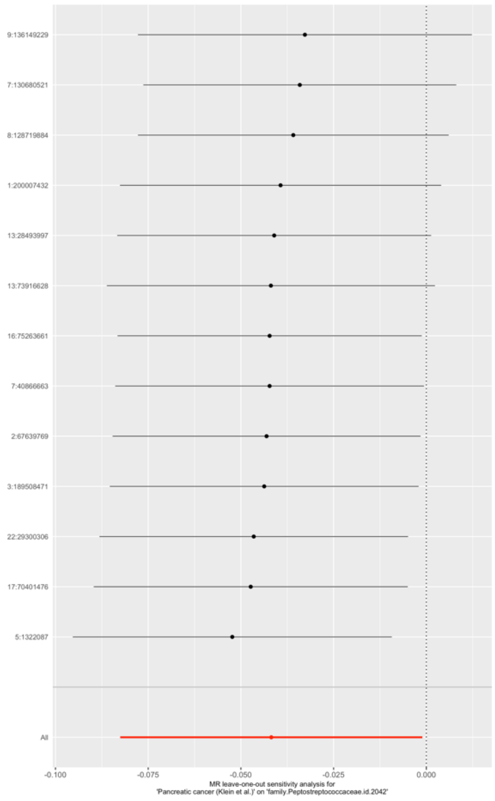


D

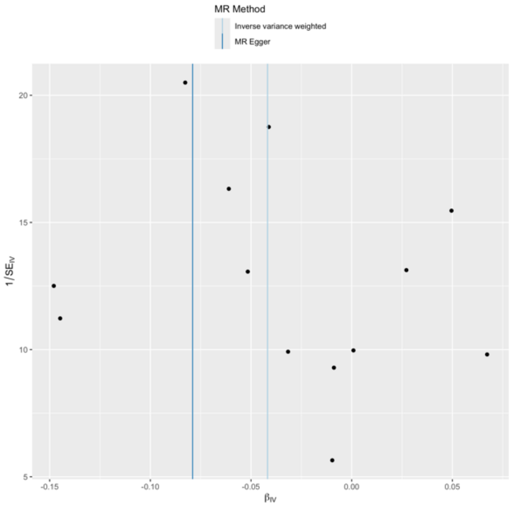

*MR = Mendelian randomization; SNP = single nucleotide polymorphism. These plots show the results of reverse MR analyses to test the causal effect of pancreatic cancer on a higher relative abundance of bacteria within the family* Peptostreptococcaceae *(*on family.Peptostreptococcaceae.id.2042*) using all SNPs associated with* pancreatic cancer *at a genome-wide significant P-value threshold (P<5x10$^{-8}$) in the Klein et al (2018) pancreatic cancer GWAS. A) Scatter plot comparing four MR methods; B) forest plot comparing the effect of individual SNP-level effect estimates derived using the Wald ratio method and in combination with the inverse variance weighted (IVW) and MR-Egger estimates; C) leave-one-out analysis to check if any one SNP is driving pleiotropy or asymmetry in the estimate; and D) funnel plot comparing the effect estimate and precision of each SNP-level Wald ratio estimate.*

**Supplementary Figure 44: Reverse MR results for effect of pancreatic cancer on genus.Erysipelatoclostridium.id.11381**

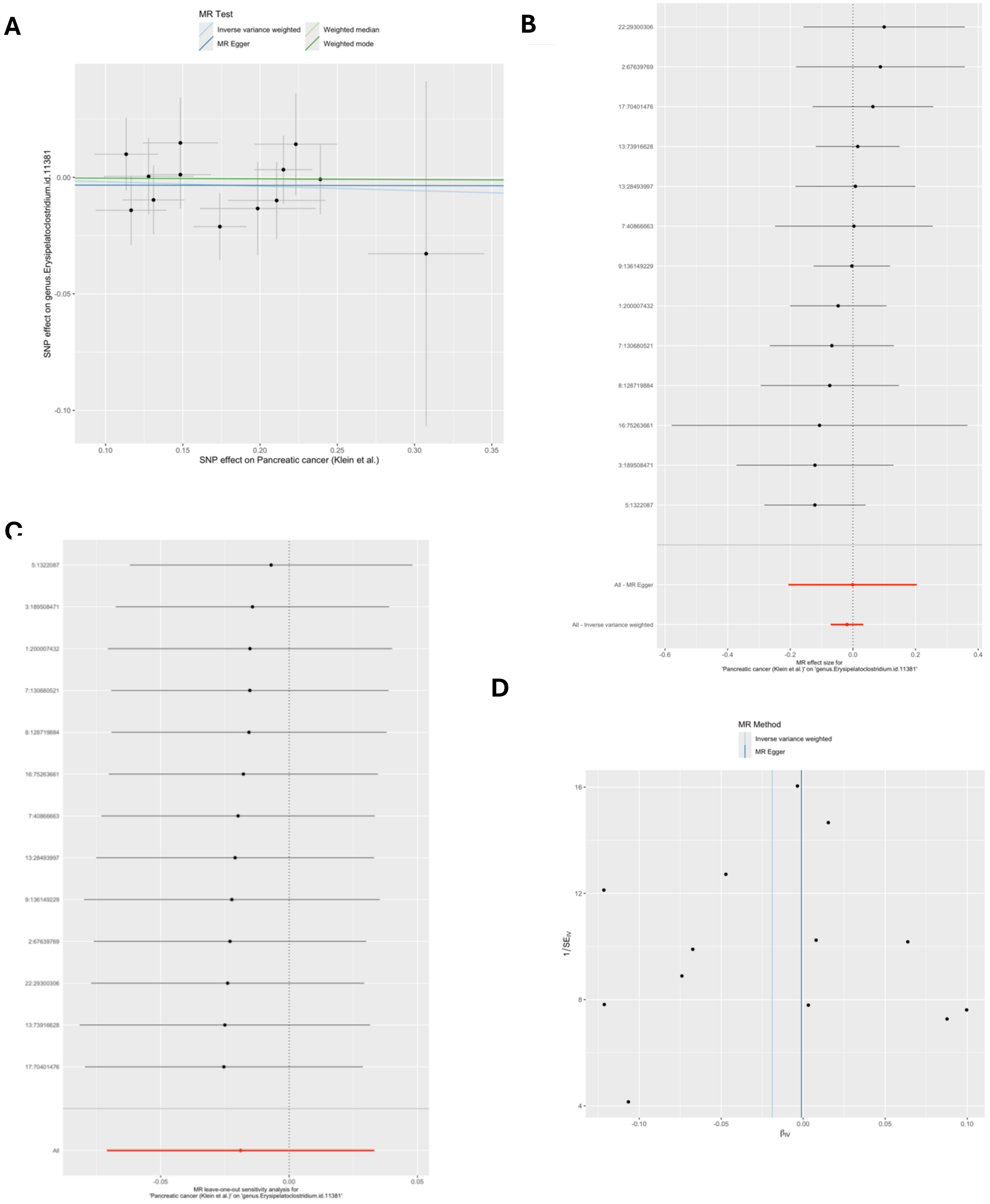


*MR = Mendelian randomization; SNP = single nucleotide polymorphism. These plots show the results of reverse MR analyses to test the causal effect of pancreatic cancer on a higher relative abundance of bacteria within the genus* Erysipelatoclostridium (on genus.Erysipelatoclostridium.id.11381*) using all SNPs associated with* pancreatic cancer *at a genome-wide significant P-value threshold (P<5x10*$^{-8}$*) in the Klein et al. (2018) pancreatic cancer GWAS. A) Scatter plot*

*comparing four MR methods; B) forest plot comparing the effect of individual SNP-level effect estimates derived using the Wald ratio method and in combination with the inverse variance weighted (IVW) and MR-Egger estimates; C) leave-one-out analysis to check if any one SNP is driving pleiotropy or asymmetry in the estimate; and D) funnel plot comparing the effect estimate and precision of each SNP-level Wald ratio estimate.*

**Supplementary Figure 45: Reverse MR results for effect of lung cancer on G.Parabacteroides.RNT**

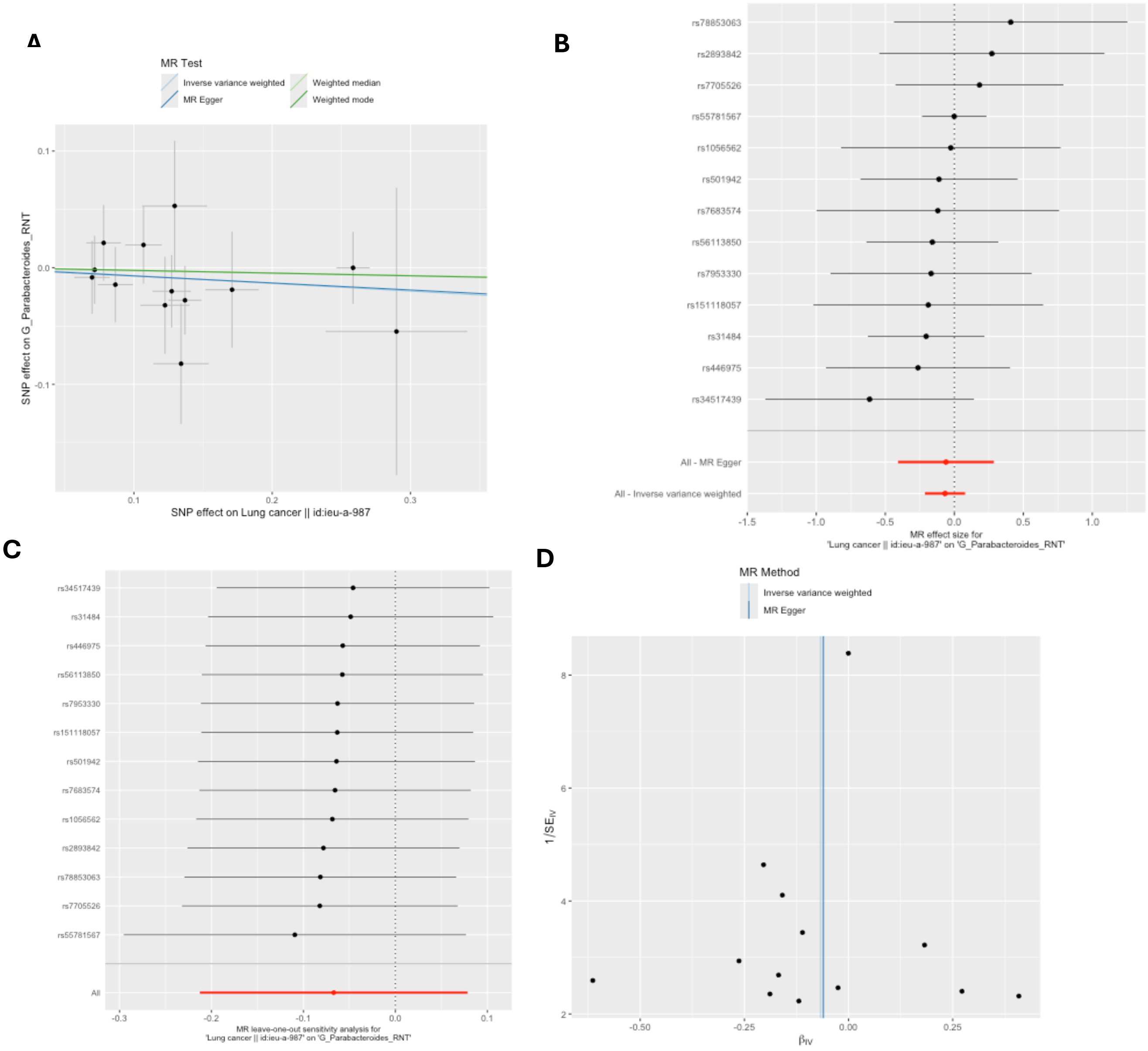


*MR = Mendelian randomization; SNP = single nucleotide polymorphism; G = genus; RNT = reverse normal transformed. These plots show the results of reverse MR analyses to test the causal effect of lung cancer on a higher relative abundance of bacteria within the genus* Parabacteroides (G.Parabacteroides.RNT*) using all SNPs associated with* lung cancer *at a genome-wide significant P-value threshold (P<5x10*$^{-8}$*) in the Transdisciplinary Research Into Cancer of the Lung (TRICL) lung cancer GWAS (accessed by IEU Open GWAS using id: ieu-a-987). A) Scatter plot comparing four MR methods; B) forest plot comparing the effect of individual SNP-level effect estimates derived using the Wald ratio method and in combination with the inverse variance weighted (IVW) and MR-Egger estimates; C) leave-one-out analysis to check if any one SNP is driving pleiotropy or asymmetry in the estimate; and D) funnel plot comparing the effect estimate and precision of each SNP-level Wald ratio estimate.*

**Supplementary Figure 46: Reverse MR results for effect of lung cancer on genus.Streptococcus.id.1853**

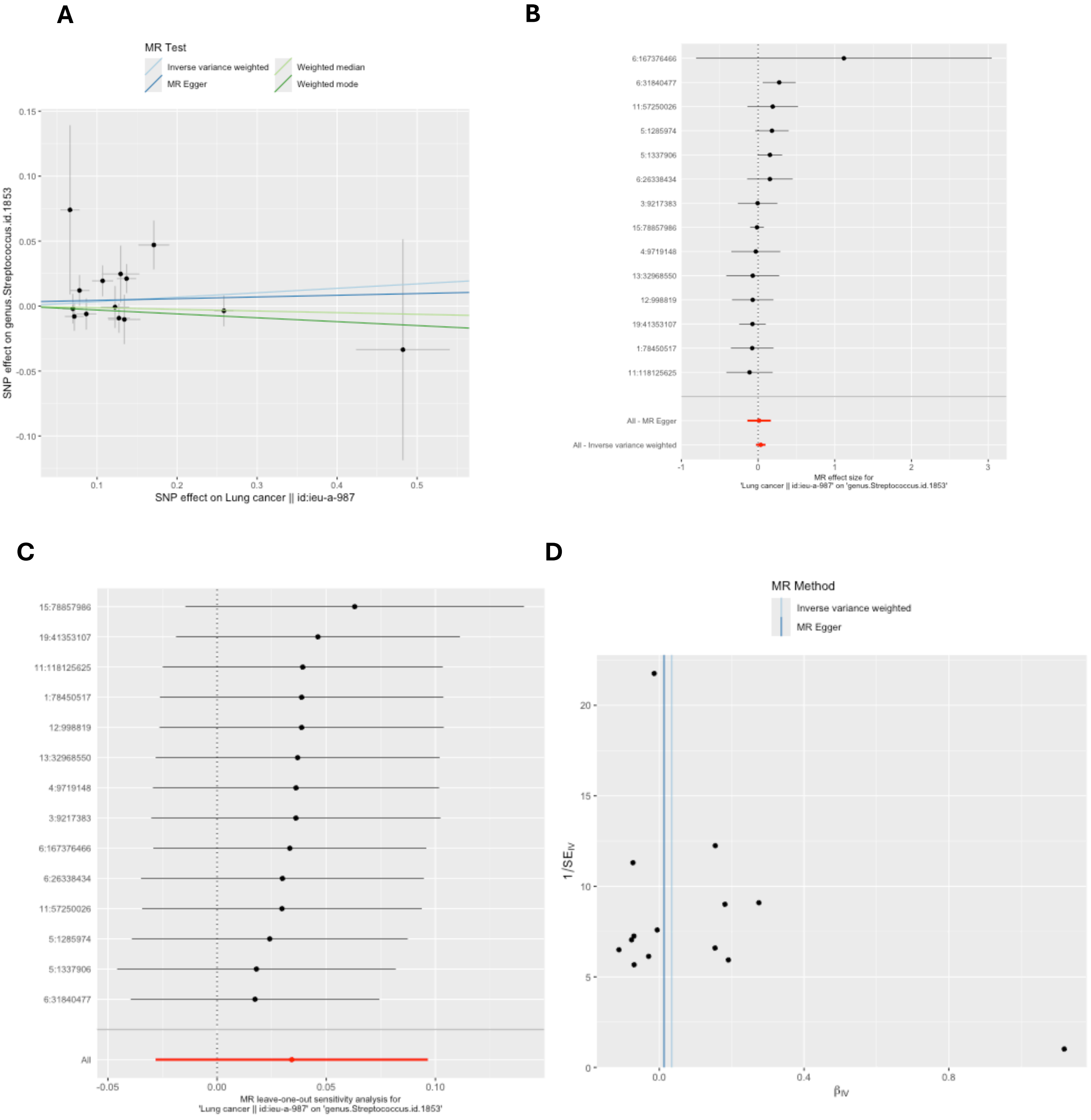


*MR = Mendelian randomization; SNP = single nucleotide polymorphism. These plots show the results of reverse MR analyses to test the causal effect of lung cancer on a higher relative abundance of bacteria within the genus* Streptococcus (genus.Streptococcus.id.1853*) using all SNPs associated with* lung cancer *at a genome-wide significant P-value threshold (P<5x10$^{-8}$) in the Transdisciplinary Research Into Cancer of the Lung (TRICL) lung cancer GWAS (accessed by IEU Open GWAS using id: ieu-a-987). A) Scatter plot comparing four MR methods; B) forest plot comparing the effect of individual SNP-level effect estimates derived using the Wald ratio method and in combination with the inverse variance weighted (IVW)*

*and MR-Egger estimates; C) leave-one-out analysis to check if any one SNP is driving pleiotropy or asymmetry in the estimate; and D) funnel plot comparing the effect estimate and precision of each SNP-level Wald ratio estimate.*

**Supplementary Figure 47: Reverse MR results for effect of lung cancer on family.Streptococcaceae.id.1850**

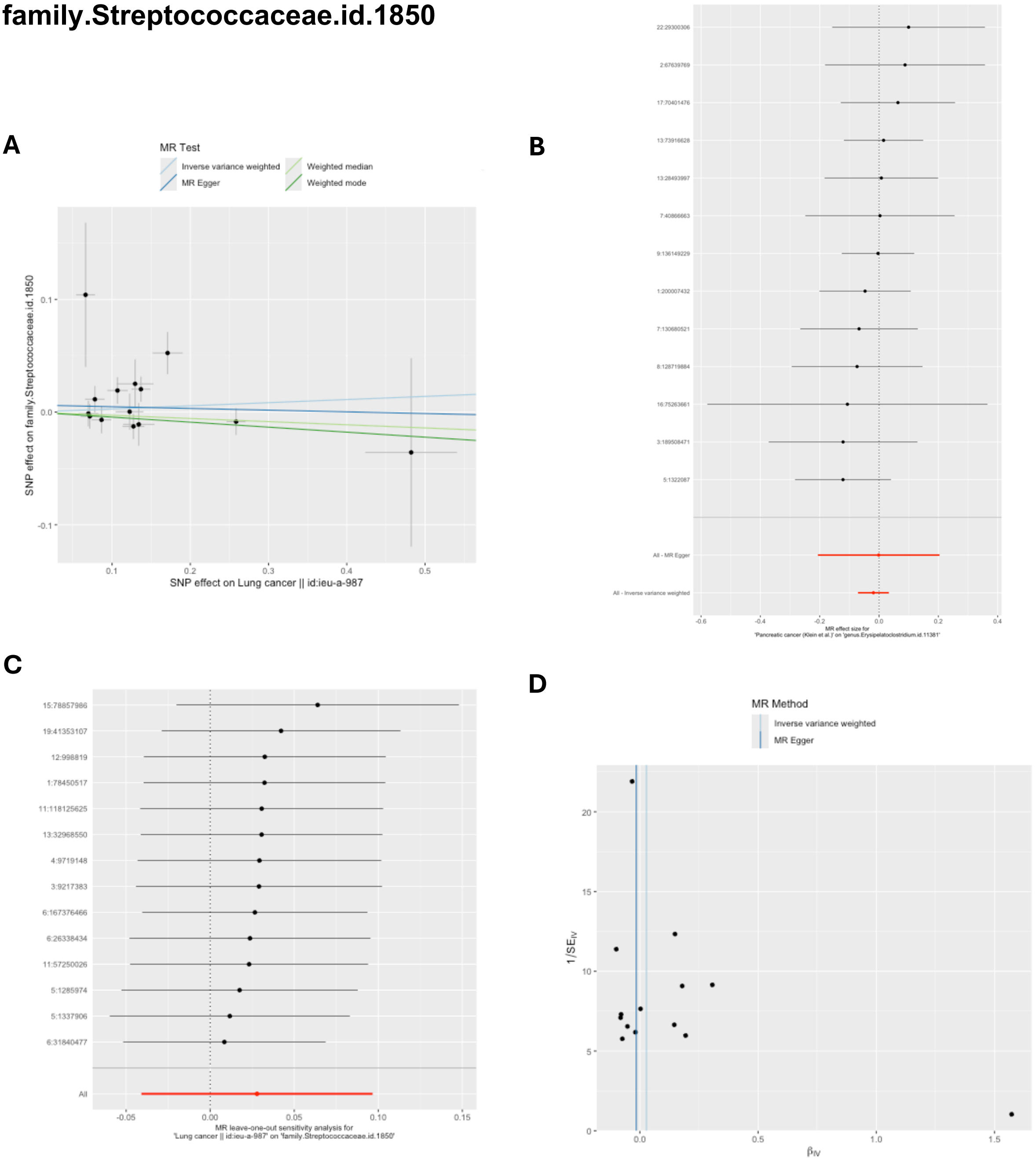


*MR = Mendelian randomization; SNP = single nucleotide polymorphism. These plots show the results of reverse MR analyses to test the causal effect of lung cancer on a higher relative abundance of bacteria within the family Streptococcaceae (family.Streptococcaceae.id.1850) using all SNPs associated with* lung cancer *at a genome-wide significant P-value threshold (P<5x10$^{-8}$) in the Transdisciplinary Research Into Cancer of the Lung (TRICL) lung cancer GWAS (accessed by IEU Open GWAS using id: ieu-a-987). A) Scatter plot comparing four MR methods; B) forest plot comparing the effect of individual SNP-level effect estimates derived using the Wald ratio method and in combination with the inverse variance weighted (IVW) and MR-Egger estimates; C) leave-one-out analysis to check if any one SNP is driving pleiotropy or asymmetry in the estimate; and D) funnel plot comparing the effect estimate and precision of each SNP-level Wald ratio estimate.*

### Supplementary Figure 48: Reverse MR results for effect of breast cancer on genus.Enterorhabdus.id.820

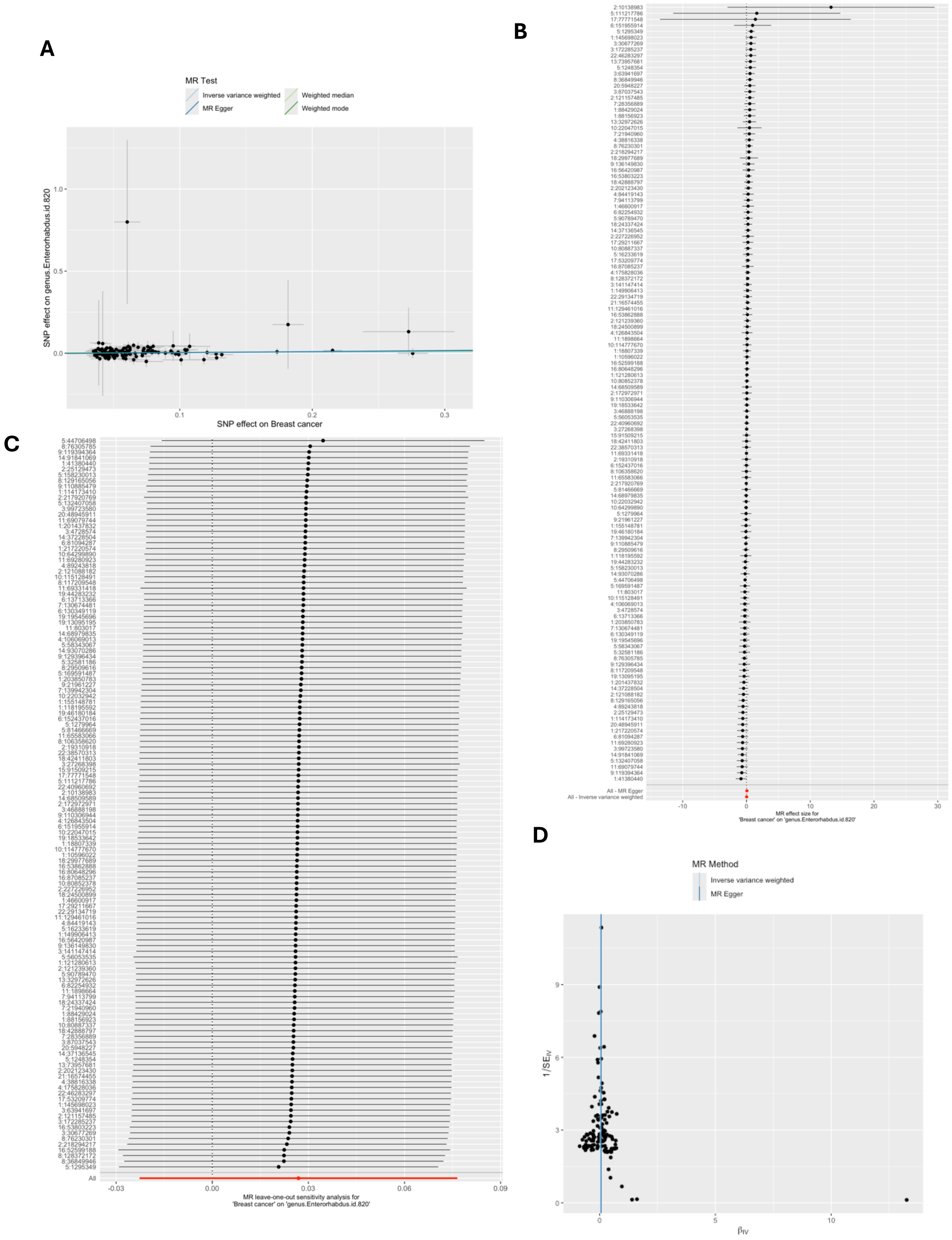

*MR = Mendelian randomization; SNP = single nucleotide polymorphism. These plots show the results of reverse MR analyses to test the causal effect of breast cancer on a higher relative abundance of bacteria within the genus Enterorhabdus (genus.Enterorhabdus.id.820) using all SNPs associated with breast cancer at a genome-wide significant P-value threshold ($P<5x10^{-8}$) in the Breast Cancer Association Consortium (BCAC) breast cancer GWAS (Michailidou et al. 2017, accessed by IEU Open GWAS using id: ieu-a-1126). A) Scatter plot comparing four MR methods; B) forest plot comparing the effect of individual SNP-level effect estimates derived using the Wald ratio method and in combination with the inverse variance weighted (IVW) and MR-Egger estimates; C) leave-one-out analysis to check if any one SNP is driving pleiotropy or asymmetry in the estimate; and D) funnel plot comparing the effect estimate and precision of each SNP-level Wald ratio estimate.*

### Supplementary Figure 49: Reverse MR results for effect of breast cancer on family.Oxalobacteraceae.id.2966

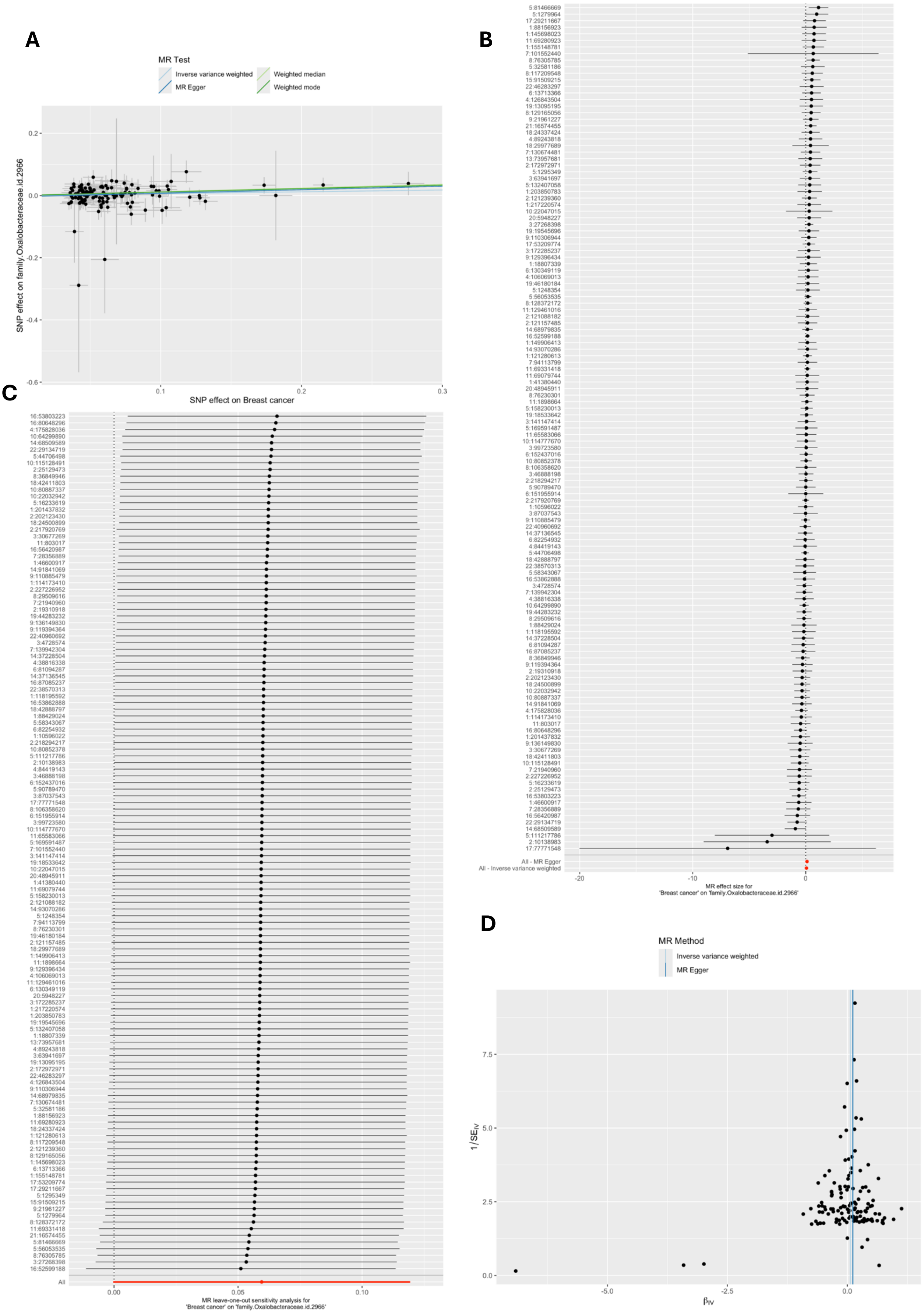

*MR = Mendelian randomization; SNP = single nucleotide polymorphism. These plots show the results of reverse MR analyses to test the causal effect of breast cancer on a higher relative abundance of bacteria within the family Oxalobacteraceae (family.Oxalobacteraceae.id.2966) using all SNPs associated with breast cancer at a genome-wide significant P-value threshold ($P<5x10^{-8}$) in the Breast Cancer Association Consortium (BCAC) breast cancer GWAS (Michailidou et al. 2017, accessed by IEU Open GWAS using id: ieu-a-1126). A) Scatter plot comparing four MR methods; B) forest plot comparing the effect of individual SNP-level effect estimates derived using the Wald ratio method and in combination with the inverse variance weighted (IVW) and MR-Egger estimates; C) leave-one-out analysis to check if any one SNP is driving pleiotropy or asymmetry in the estimate; and D) funnel plot comparing the effect estimate and precision of each SNP-level Wald ratio estimate.*

### Supplementary Figure 50: Reverse MR results for effect of colorectal cancer on family.Oxalobacteraceae.id.2966

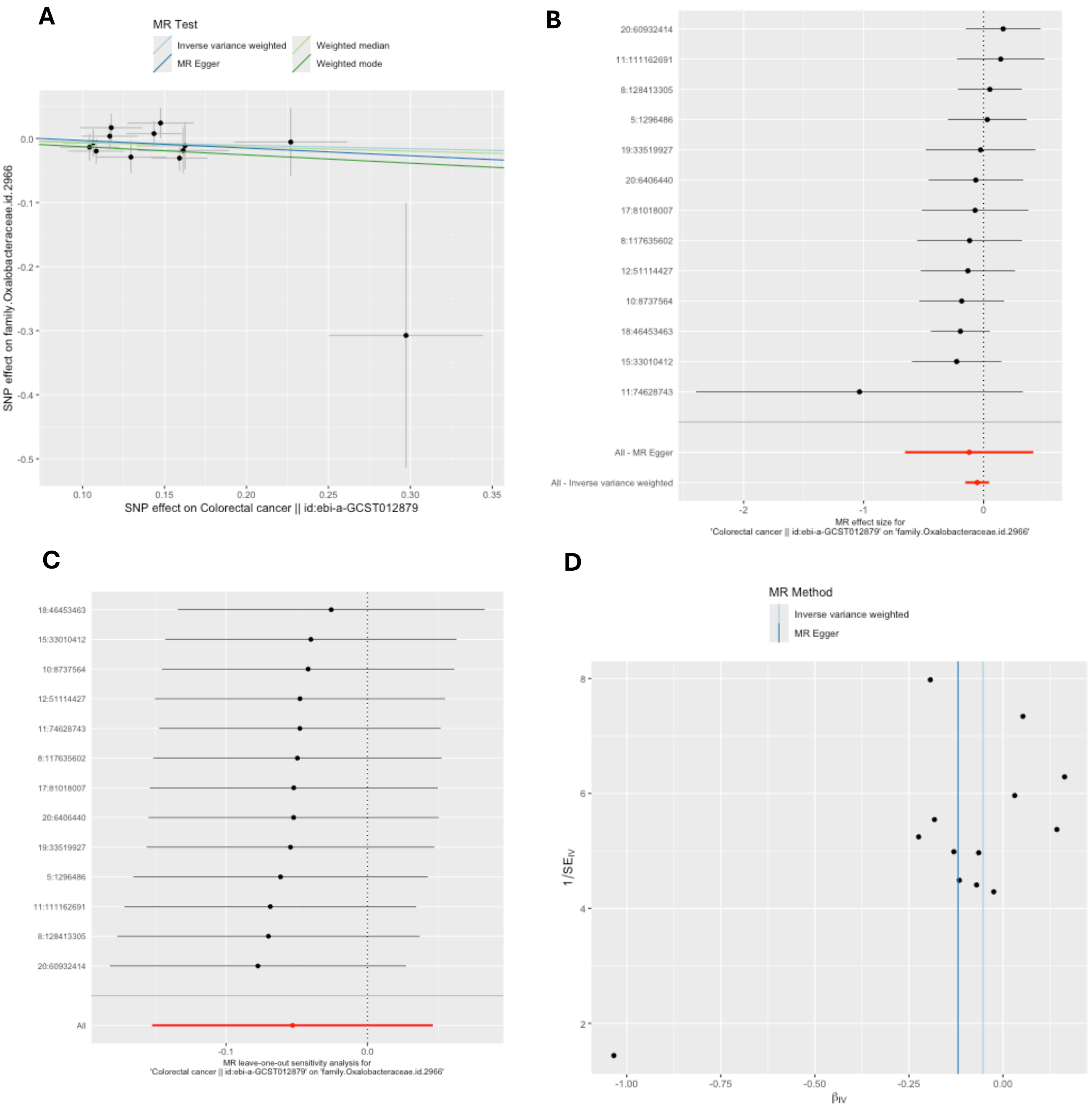


*MR = Mendelian randomization; SNP = single nucleotide polymorphism. These plots show the results of reverse MR analyses to test the causal effect of colorectal cancer on a higher relative abundance of bacteria within the family Oxalobacteraceae (family.Oxalobacteraceae.id.2966) using all SNPs associated with colorectal cancer at a genome-wide significant P-value threshold ($P<5x10^{-8}$) in the Huyghe et al. (2018) colorectal cancer GWAS (accessed by IEU Open GWAS using id: ebi-a-GCST012879). A) Scatter plot comparing four MR methods; B) forest plot comparing the effect of individual SNP-level effect estimates derived using the Wald ratio*

*method and in combination with the inverse variance weighted (IVW) and MR-Egger estimates; C) leave-one-out analysis to check if any one SNP is driving pleiotropy or asymmetry in the estimate; and D) funnel plot comparing the effect estimate and precision of each SNP-level Wald ratio estimate.*

**Supplementary Figure 51: Reverse MR results for effect of colorectal cancer on genus.Ruminococcustorquesgroup.id.14377**

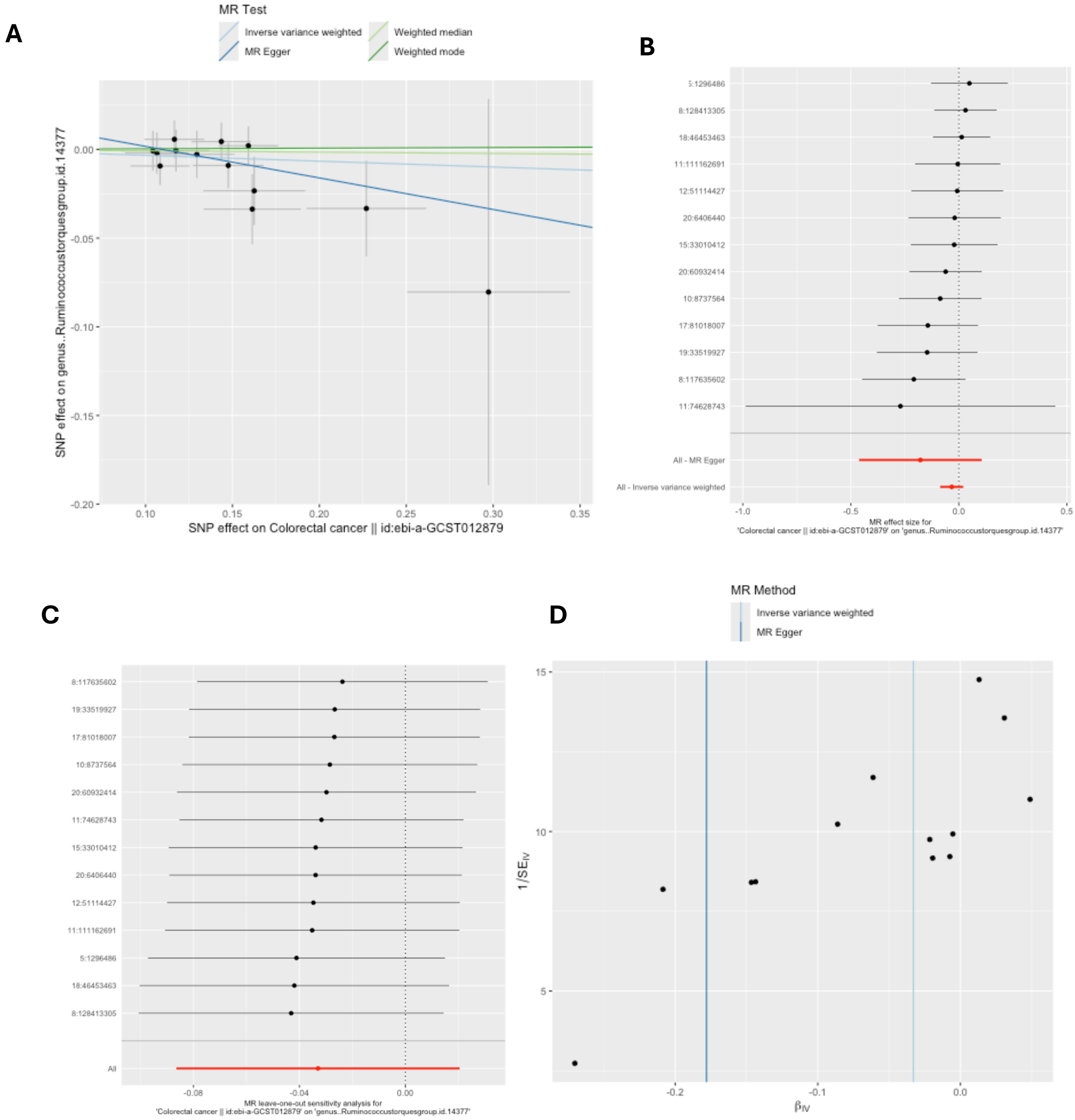


*MR = Mendelian randomization; SNP = single nucleotide polymorphism. These plots show the results of reverse MR analyses to test the causal effect of colorectal cancer on a higher relative abundance of bacteria within the genus Ruminococcus in the Torques group (genus..Ruminococcustorquesgroup.id.14377) using all SNPs associated with colorectal cancer at a genome-wide significant P-value threshold ($P<5x10^{-8}$) in the Huyghe et al. (2018) colorectal cancer GWAS (accessed by IEU Open GWAS using id: ebi-a-GCST012879). A) Scatter plot comparing four MR methods; B) forest plot comparing the effect of individual SNP-level effect estimates derived using the Wald ratio method and in combination with the inverse variance weighted (IVW) and MR-Egger estimates; C) leave-one-out analysis to check if any*

*one SNP is driving pleiotropy or asymmetry in the estimate; and D) funnel plot comparing the effect estimate and precision of each SNP-level Wald ratio estimate.*

### Supplementary Figure 52: Reverse MR results for effect of colorectal cancer on phylum.Actinobacteria.id.400

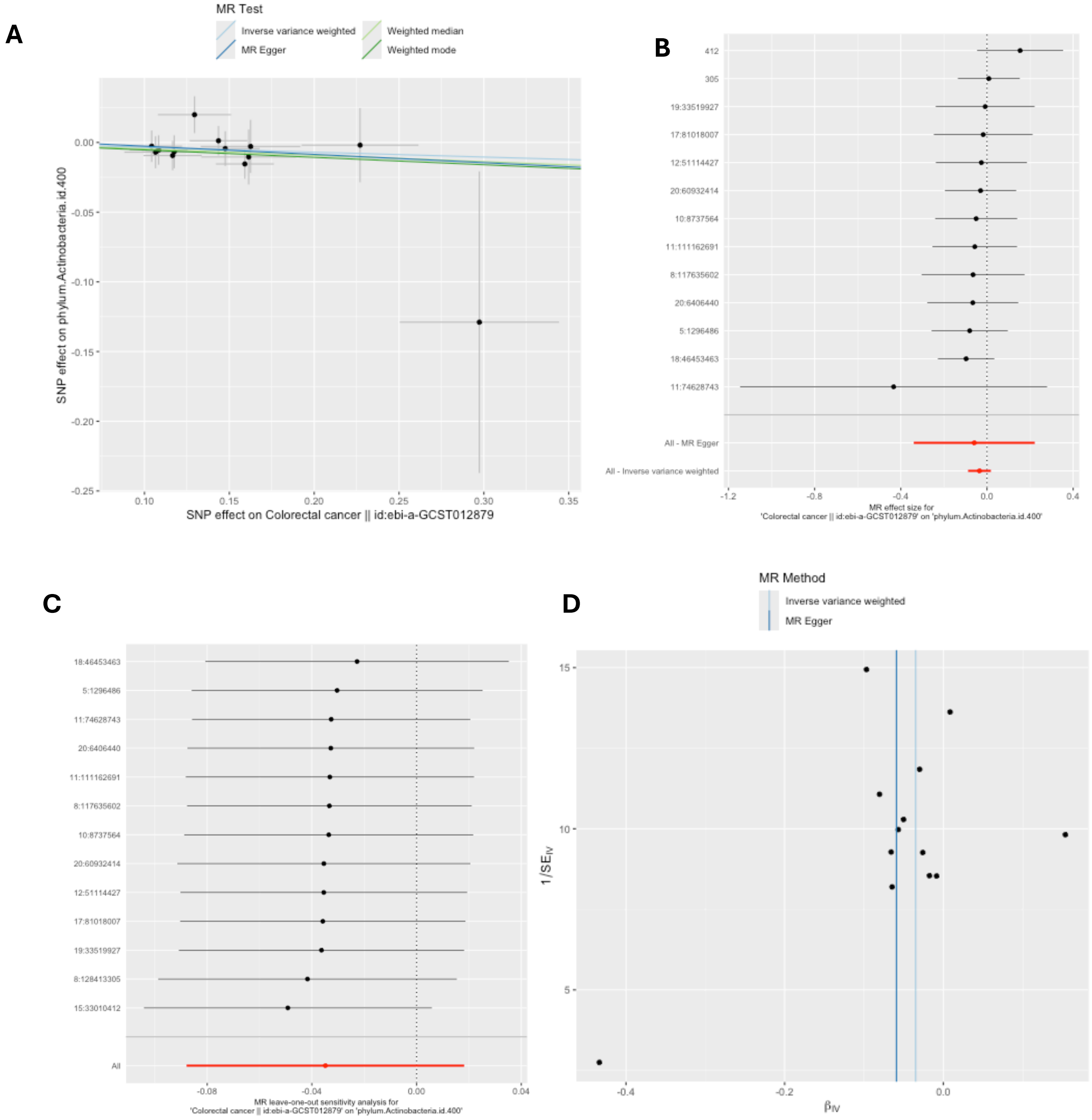


*MR = Mendelian randomization; SNP = single nucleotide polymorphism. These plots show the results of reverse MR analyses to test the causal effect of colorectal cancer on a higher relative abundance of bacteria within the phylum Actinobacteria (phylum.Actinobacteria.id.400) using all SNPs associated with colorectal cancer at a genome-wide significant P-value threshold ($P<5x10^{-8}$) in the Huyghe et al. (2018) colorectal cancer GWAS (accessed by IEU Open GWAS using id: ebi-a-GCST012879). A) Scatter plot comparing four MR methods; B) forest plot comparing the effect of individual SNP-level effect estimates derived using the Wald ratio method and in combination with the inverse variance weighted (IVW) and MR-Egger estimates; C) leave-one-out analysis to check if any one SNP is driving pleiotropy or*

*asymmetry in the estimate; and D) funnel plot comparing the effect estimate and precision of each SNP-level Wald ratio estimate.*

### Supplementary Figure 53: Reverse MR results for effect of colorectal cancer on class.Actinobacteria.id.419

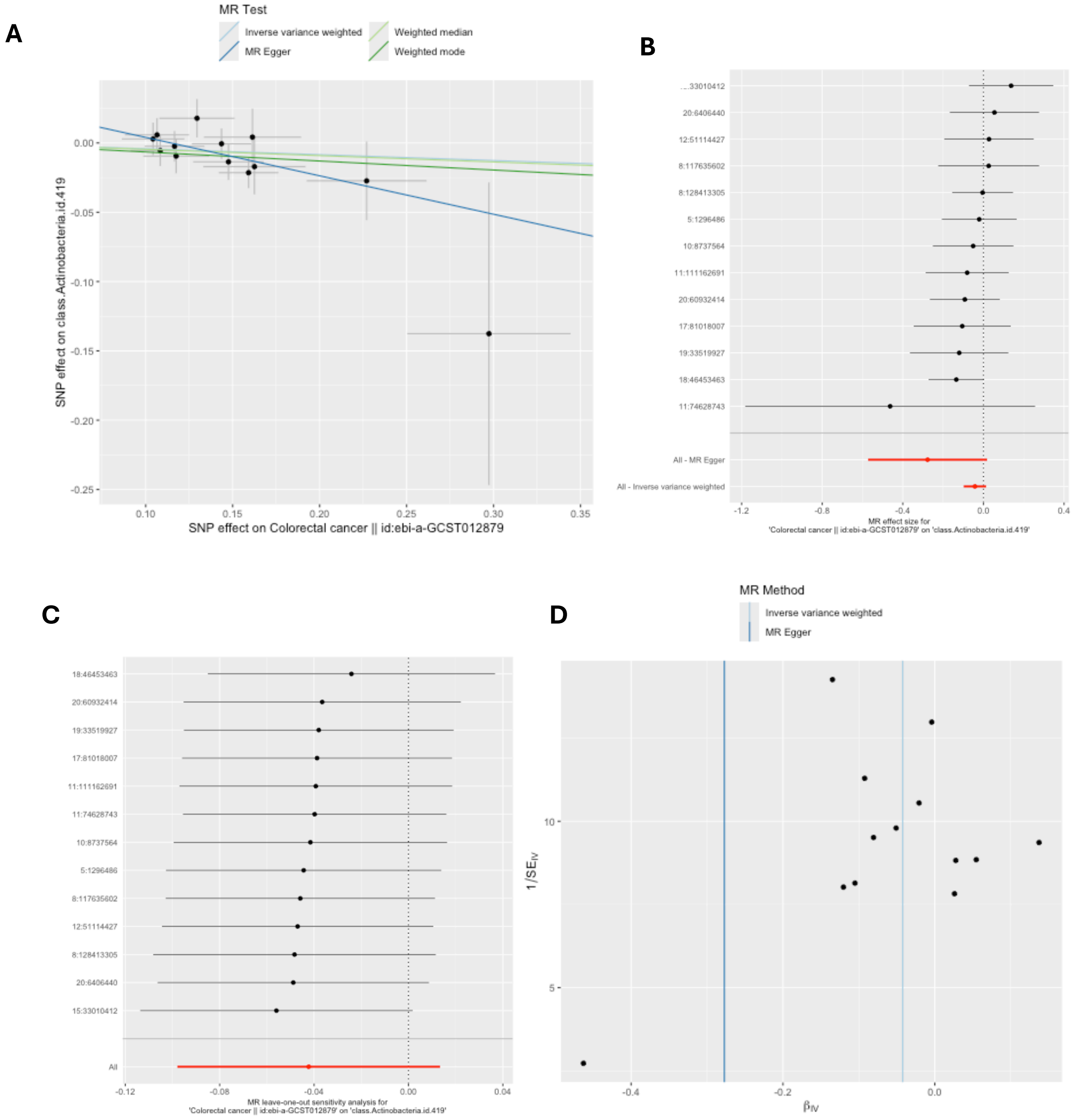


*MR = Mendelian randomization; SNP = single nucleotide polymorphism. These plots show the results of reverse MR analyses to test the causal effect of colorectal cancer on a higher relative abundance of bacteria within the class Actinobacteria (class.Actinobacteria.id.419) using all SNPs associated with colorectal cancer at a genome-wide significant P-value threshold ($P<5x10^{-8}$) in the Huyghe et al. (2018) colorectal cancer GWAS (accessed by IEU Open GWAS using id: ebi-a-GCST012879). A) Scatter plot comparing four MR methods; B) forest plot comparing the effect of individual SNP-level effect estimates derived using the Wald ratio method and in combination with the inverse variance weighted (IVW) and MR-Egger estimates; C) leave-one-out analysis to check if any one SNP is driving pleiotropy or*

*asymmetry in the estimate; and D) funnel plot comparing the effect estimate and precision of each SNP-level Wald ratio estimate.*

### Supplementary Figure 54: Reverse MR results for effect of endometrial cancer on family.Oxalobacteraceae.id.2966

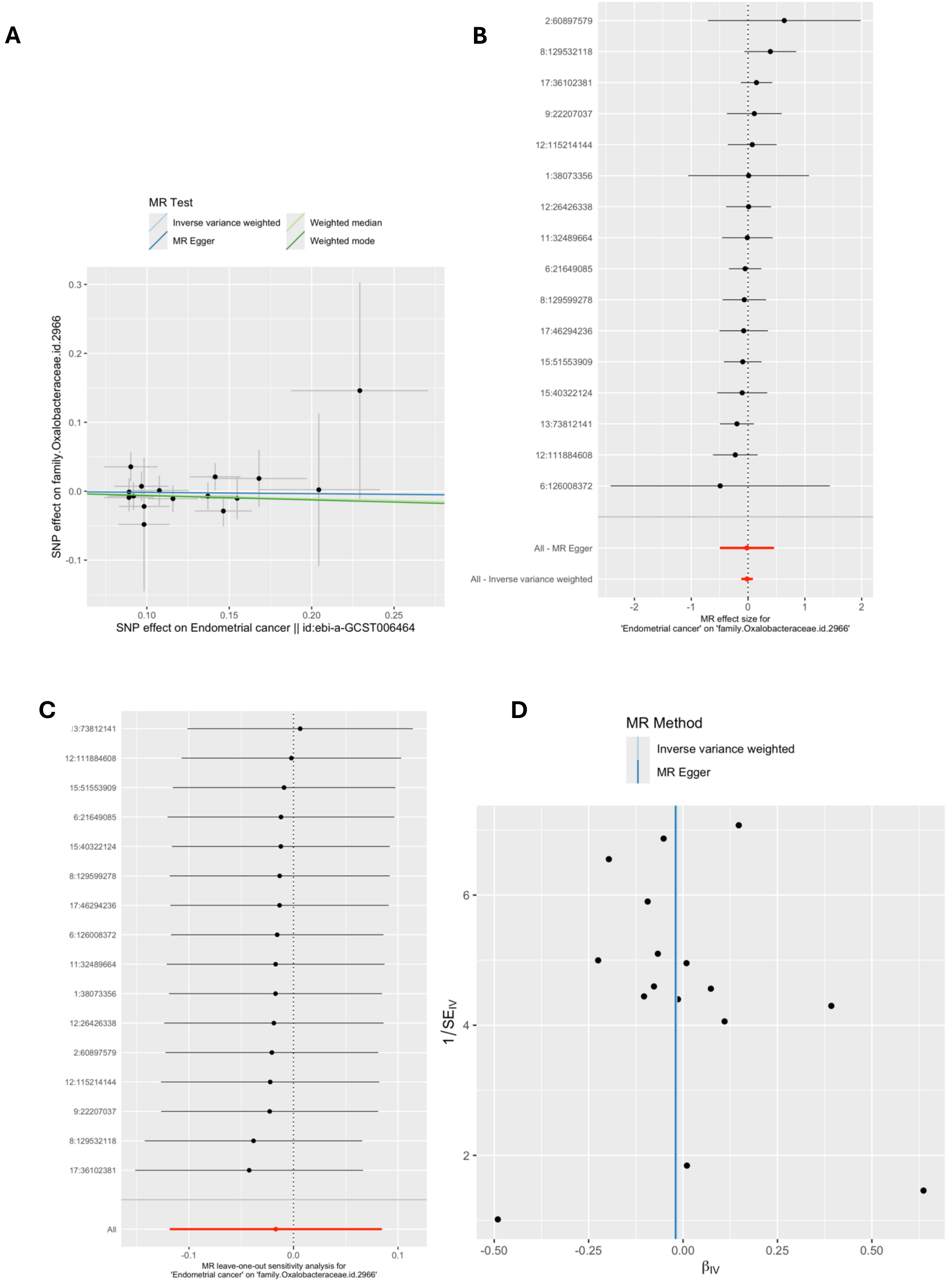


*MR = Mendelian randomization; SNP = single nucleotide polymorphism. These plots show the results of reverse MR analyses to test the causal effect of endometrial cancer on a higher relative abundance of bacteria within the* family

Oxalobacteraceae *(*family.Oxalobacteraceae.id.2966*) using all SNPs associated with endometrial cancer at a genome-wide significant P-value threshold ($P<5x10^{-8}$) in the O'Mara et al. (2018) endometrial cancer GWAS (accessed by IEU Open GWAS using id: ebi-a-GCST006464). A) Scatter plot comparing four MR methods; B) forest plot comparing the effect of individual SNP-level effect estimates derived using the Wald ratio method and in combination with the inverse variance weighted (IVW) and MR-Egger estimates; C) leave-one-out analysis to check if any one SNP is driving pleiotropy or asymmetry in the estimate; and D) funnel plot comparing the effect estimate and precision of each SNP-level Wald ratio estimate.*

**Supplementary Figure 55: Reverse MR results for effect of endometrial cancer on family.Streptococcaceae.id.1850**

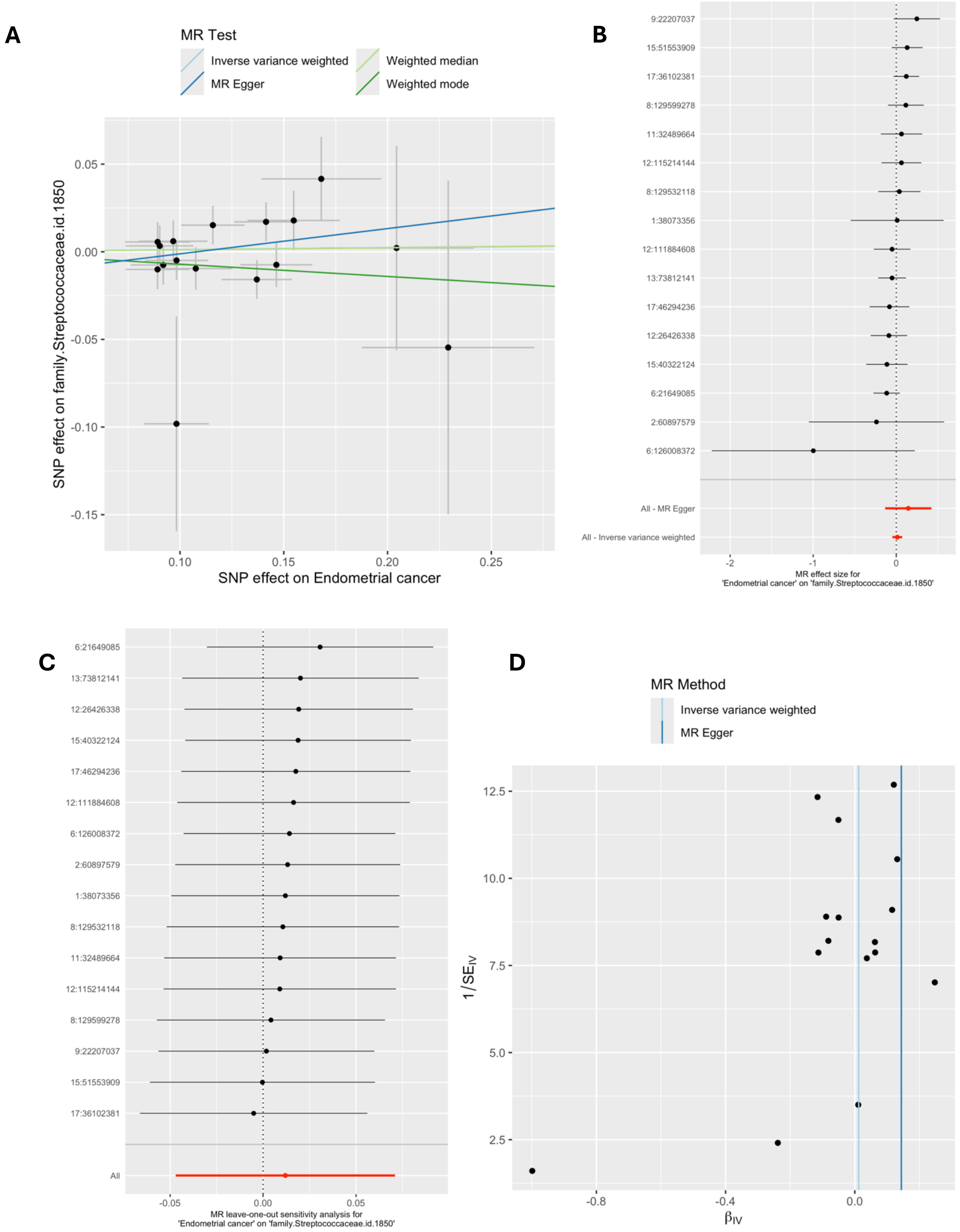


*MR = Mendelian randomization; SNP = single nucleotide polymorphism. These plots show the results of reverse MR analyses to test the causal effect of endometrial cancer on a higher relative abundance of bacteria within the family Streptococcaceae (family.Streptococcaceae.id.1850) using all SNPs associated with*

*endometrial cancer at a genome-wide significant P-value threshold ($P<5x10^{-8}$) in the O'Mara et al. (2018) endometrial cancer GWAS (accessed by IEU Open GWAS using id: ebi-a-GCST006464). A) Scatter plot comparing four MR methods; B) forest plot comparing the effect of individual SNP-level effect estimates derived using the Wald ratio method and in combination with the inverse variance weighted (IVW) and MR-Egger estimates; C) leave-one-out analysis to check if any one SNP is driving pleiotropy or asymmetry in the estimate; and D) funnel plot comparing the effect estimate and precision of each SNP-level Wald ratio estimate.*

### Supplementary Figure 56: Reverse MR results for effect of endometrial cancer on genus.Enterorhabdus.id.820

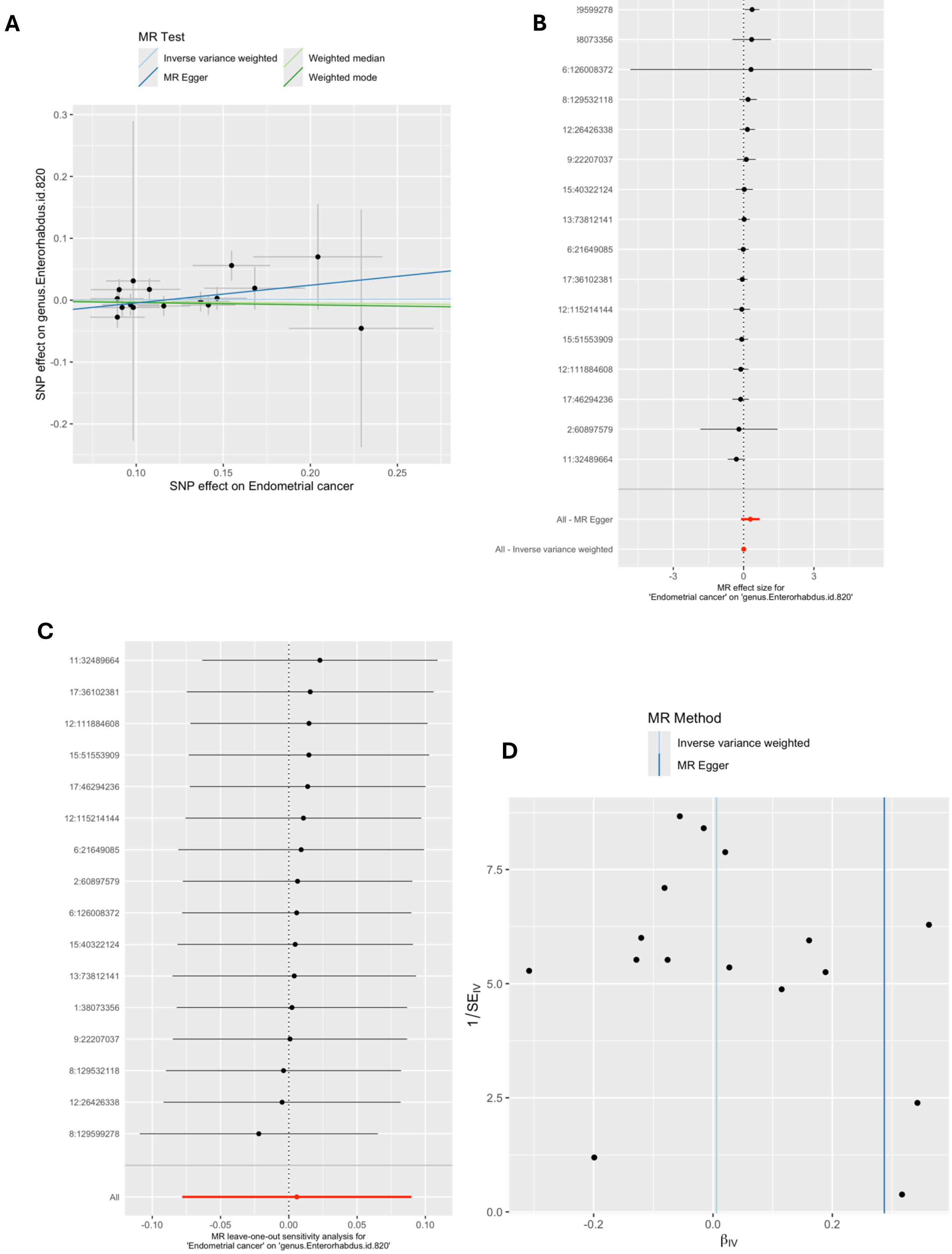

*MR = Mendelian randomization; SNP = single nucleotide polymorphism. These plots show the results of reverse MR analyses to test the causal effect of endometrial cancer on a higher relative abundance of bacteria within the genus Enterorhabdus (genus.Enterorhabdus.id.820) using all SNPs associated with endometrial cancer at a genome-wide significant P-value threshold ($P<5x10^{-8}$) in the O'Mara et al. (2018) endometrial cancer GWAS (accessed by IEU Open GWAS using id: ebi-a-GCST006464). A) Scatter plot comparing four MR methods; B) forest plot comparing the effect of individual SNP-level effect estimates derived using the Wald ratio method and in combination with the inverse variance weighted (IVW) and MR-Egger estimates; C) leave-one-out analysis to check if any one SNP is driving pleiotropy or asymmetry in the estimate; and D) funnel plot comparing the effect estimate and precision of each SNP-level Wald ratio estimate.*

**Supplementary Figure 57: Reverse MR results for effect of endometrial cancer on genus.Streptococcus.id.1853**

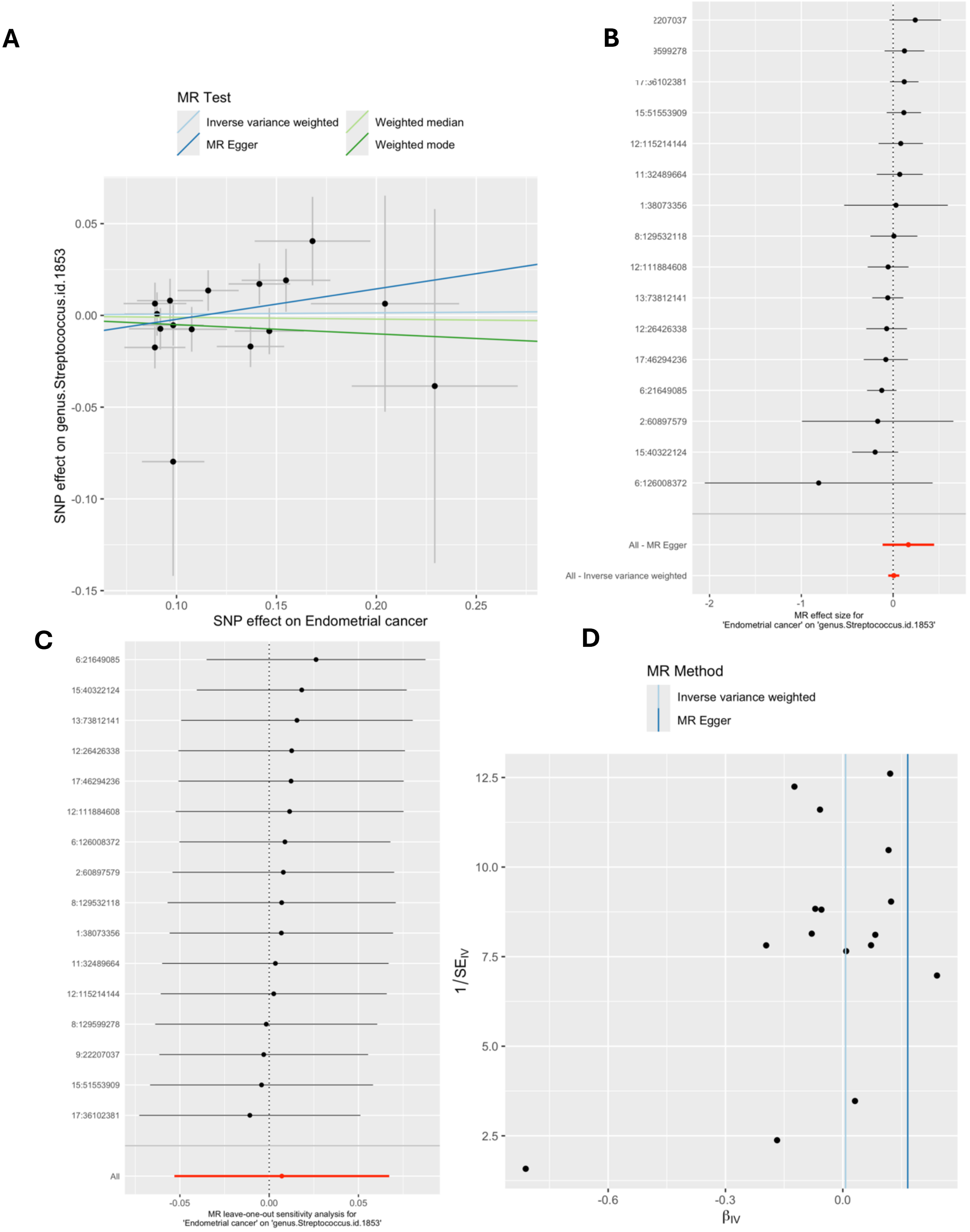


*MR = Mendelian randomization; SNP = single nucleotide polymorphism. These plots show the results of reverse MR analyses to test the causal effect of endometrial cancer on a higher relative abundance of bacteria within the genus Streptococcus (genus.Streptococcus.id.1853) using all SNPs associated with endometrial cancer at a genome-wide significant P-value threshold ($P<5x10^{-8}$) in the O'Mara et al. (2018) endometrial cancer GWAS (accessed by IEU Open GWAS using id: ebi-a-GCST006464). A) Scatter plot comparing four MR methods; B) forest plot comparing the effect of individual SNP-level effect estimates derived using the Wald ratio*

*method and in combination with the inverse variance weighted (IVW) and MR-Egger estimates; C) leave-one-out analysis to check if any one SNP is driving pleiotropy or asymmetry in the estimate; and D) funnel plot comparing the effect estimate and precision of each SNP-level Wald ratio estimate.*

**Supplementary Figure 58: Reverse MR results for effect of oesophageal cancer on genus..Ruminococcustorquesgroup.id.14377**

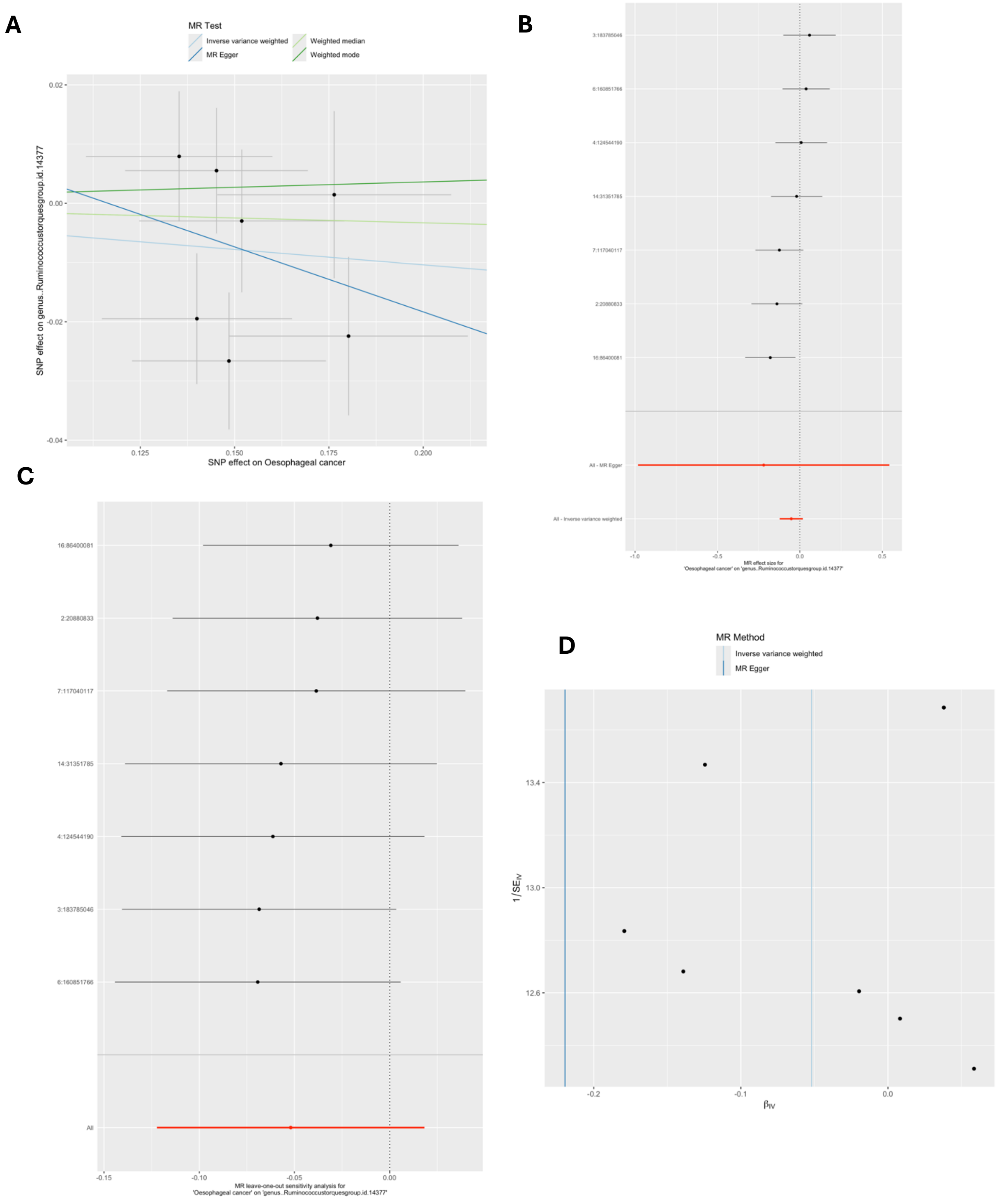


*MR = Mendelian randomization; SNP = single nucleotide polymorphism. These plots show the results of reverse MR analyses to test the causal effect of oesophageal cancer on a higher relative abundance of bacteria within the genus Ruminococcus and within the Torques group (genus..Ruminococcustorquesgroup.id.14377) using all SNPs associated with oesophageal cancer at a genome-wide significant P-value threshold (P<5x10$^{-8}$) in the Schröder et al. (2023) oesophageal cancer GWAS. A) Scatter plot comparing four MR methods; B) forest plot comparing the effect of*

*individual SNP-level effect estimates derived using the Wald ratio method and in combination with the inverse variance weighted (IVW) and MR-Egger estimates; C) leave-one-out analysis to check if any one SNP is driving pleiotropy or asymmetry in the estimate; and D) funnel plot comparing the effect estimate and precision of each SNP-level Wald ratio estimate.*

### Supplementary Figure 59: Reverse MR results for effect of prostate cancer on genus.Allisonella.id.2174

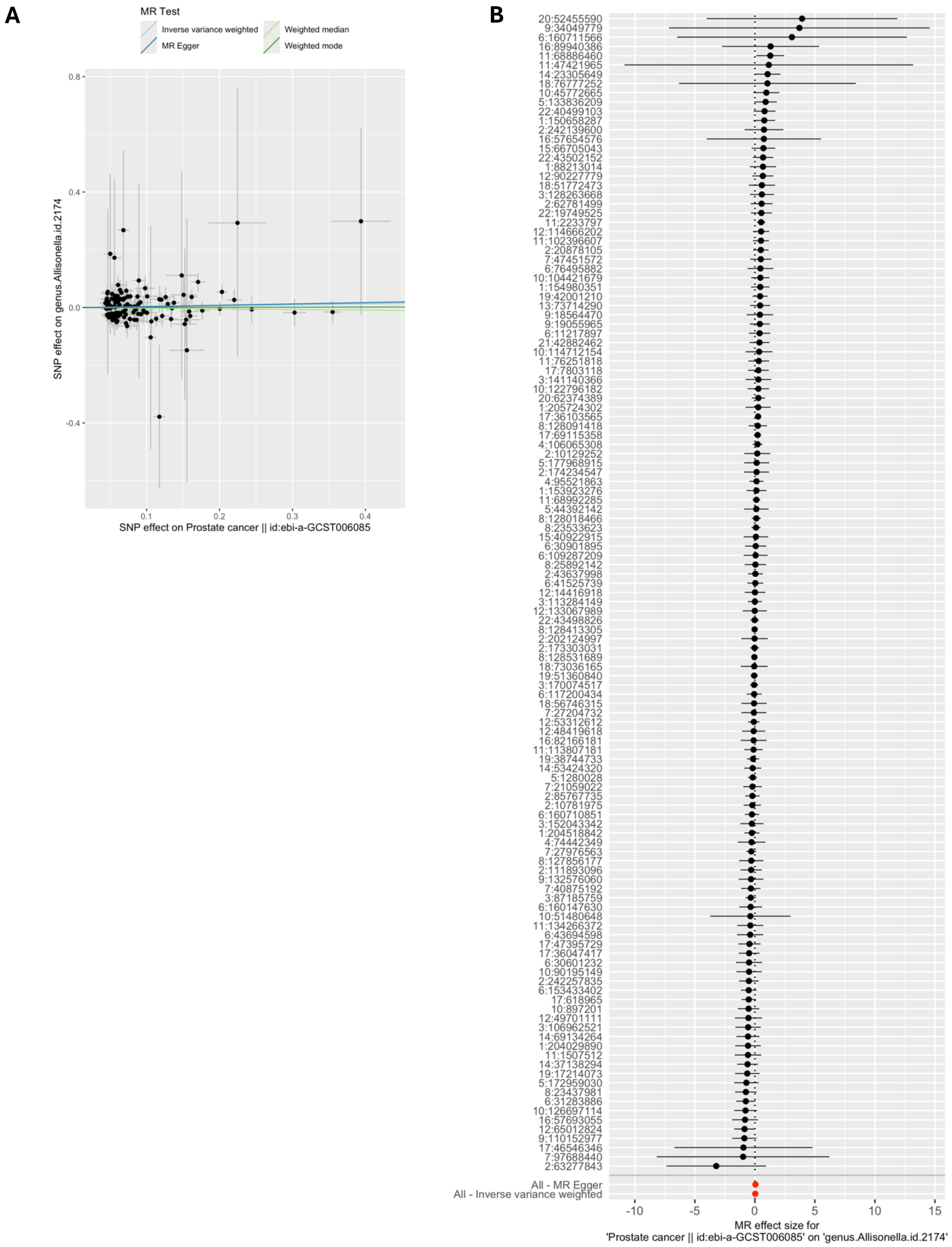

C

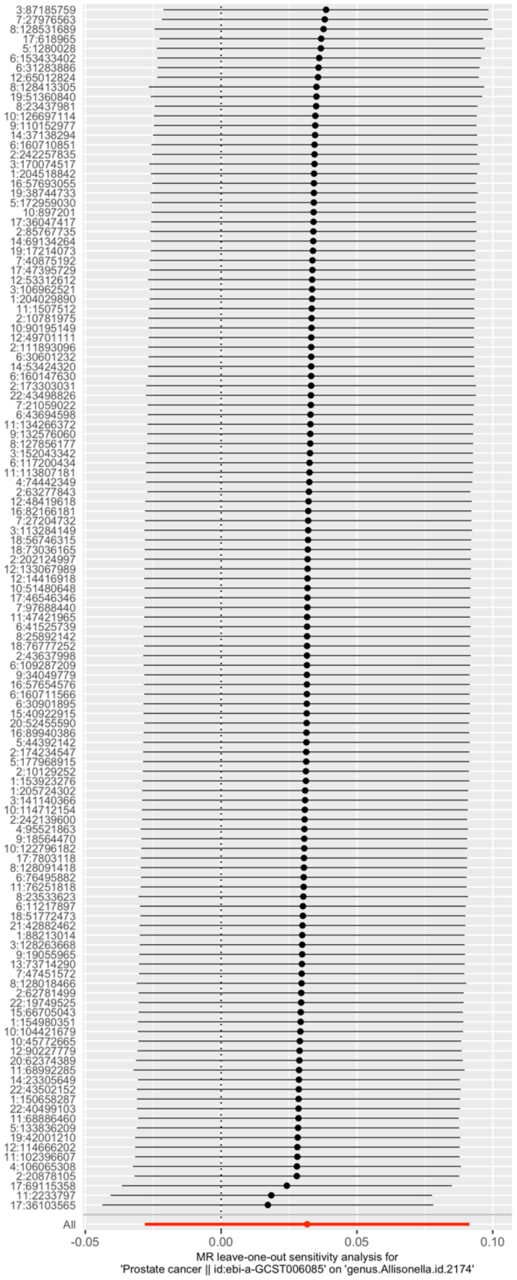


D

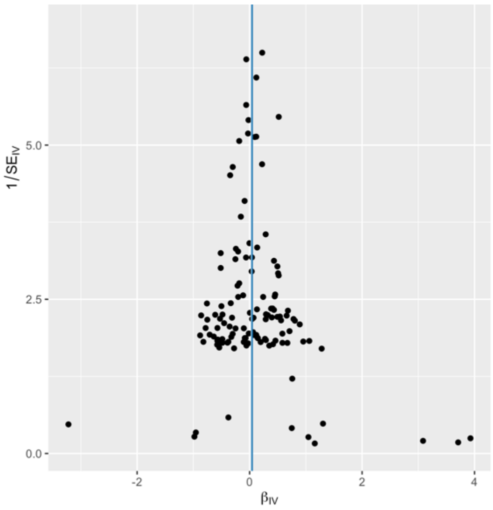


*MR = Mendelian randomization; SNP = single nucleotide polymorphism. These plots show the results of reverse MR analyses to test the causal effect of prostate cancer on a higher relative abundance of bacteria within the genus Allisonella (genus.Allisonella.id.2174) using all SNPs associated with prostate cancer at a genome-wide significant P-value threshold ($P<5x10^{-8}$) in the Schumacher et al 2018 prostate cancer GWAS. A) Scatter plot comparing four MR methods; B) forest plot comparing the effect of individual SNP-level effect estimates derived using the Wald ratio method and in combination with the inverse variance weighted (IVW) and MR-Egger estimates; C) leave-one-out analysis to check if any one SNP is driving pleiotropy or asymmetry in the estimate; and D) funnel plot comparing the effect estimate and precision of each SNP-level Wald ratio estimate.*

### Supplementary Figure 60: Reverse MR results for effect of prostate cancer on G.Parabacteroides

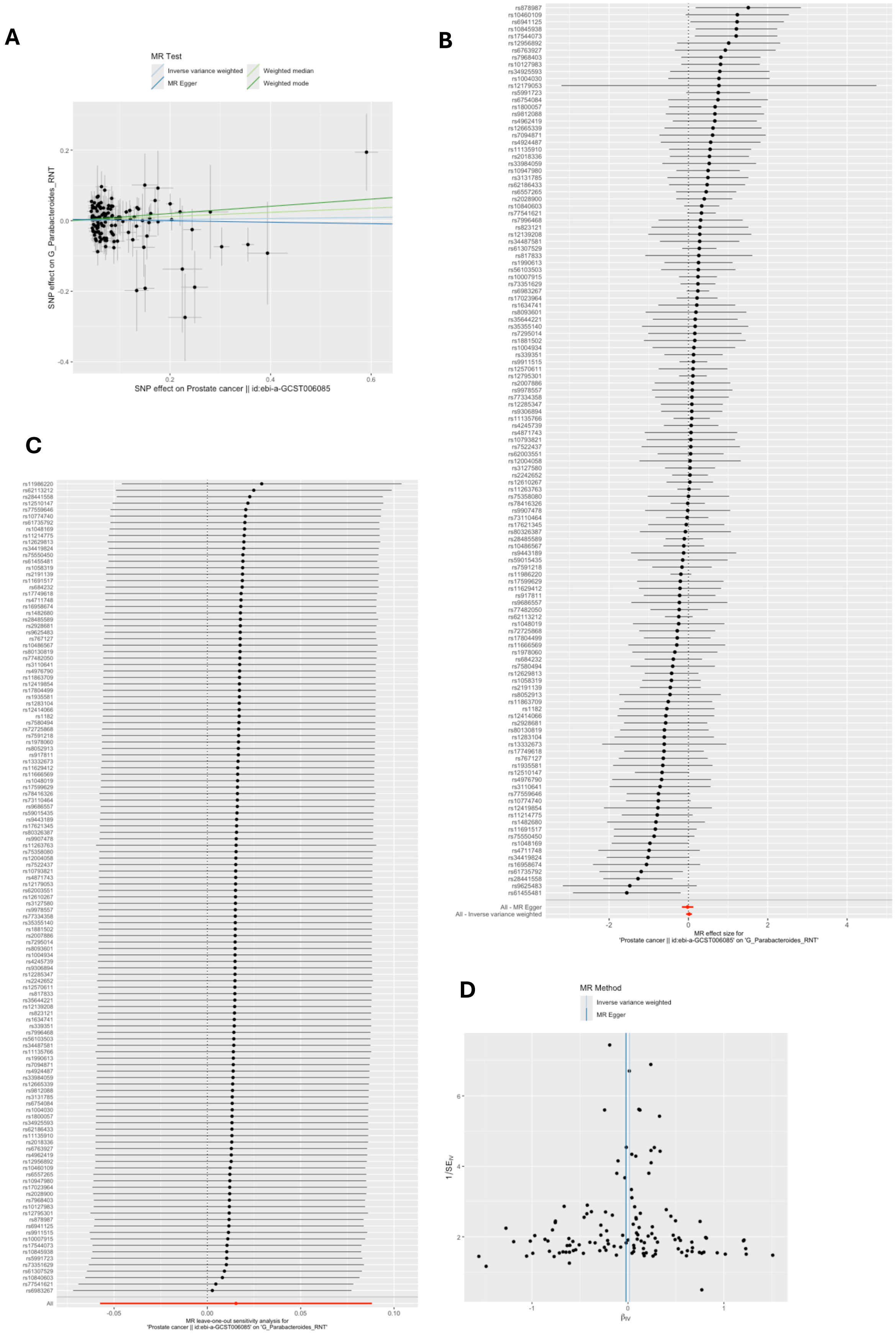

*G = genus; RNT = reverse normal transformation; MR = Mendelian randomization; SNP = single nucleotide polymorphism. These plots show the results of reverse MR analyses to test the causal effect of prostate cancer on a higher relative abundance of bacteria within the genus Parabacteroides (G_Parabacteroides_RNT) using all SNPs associated with prostate cancer at a genome-wide significant P-value threshold ($P<5x10^{-8}$) in the Schumacher et al 2018 prostate cancer GWAS. A) Scatter plot comparing four MR methods; B) forest plot comparing the effect of individual SNP-level effect estimates derived using the Wald ratio method and in combination with the inverse variance weighted (IVW) and MR-Egger estimates; C) leave-one-out analysis to check if any one SNP is driving pleiotropy or asymmetry in the estimate; and D) funnel plot comparing the effect estimate and precision of each SNP-level Wald ratio estimate.*

### Supplementary Figure 61: Reverse MR results for effect of prostate cancer on G.unclassified.F.Porphyromonadaceae

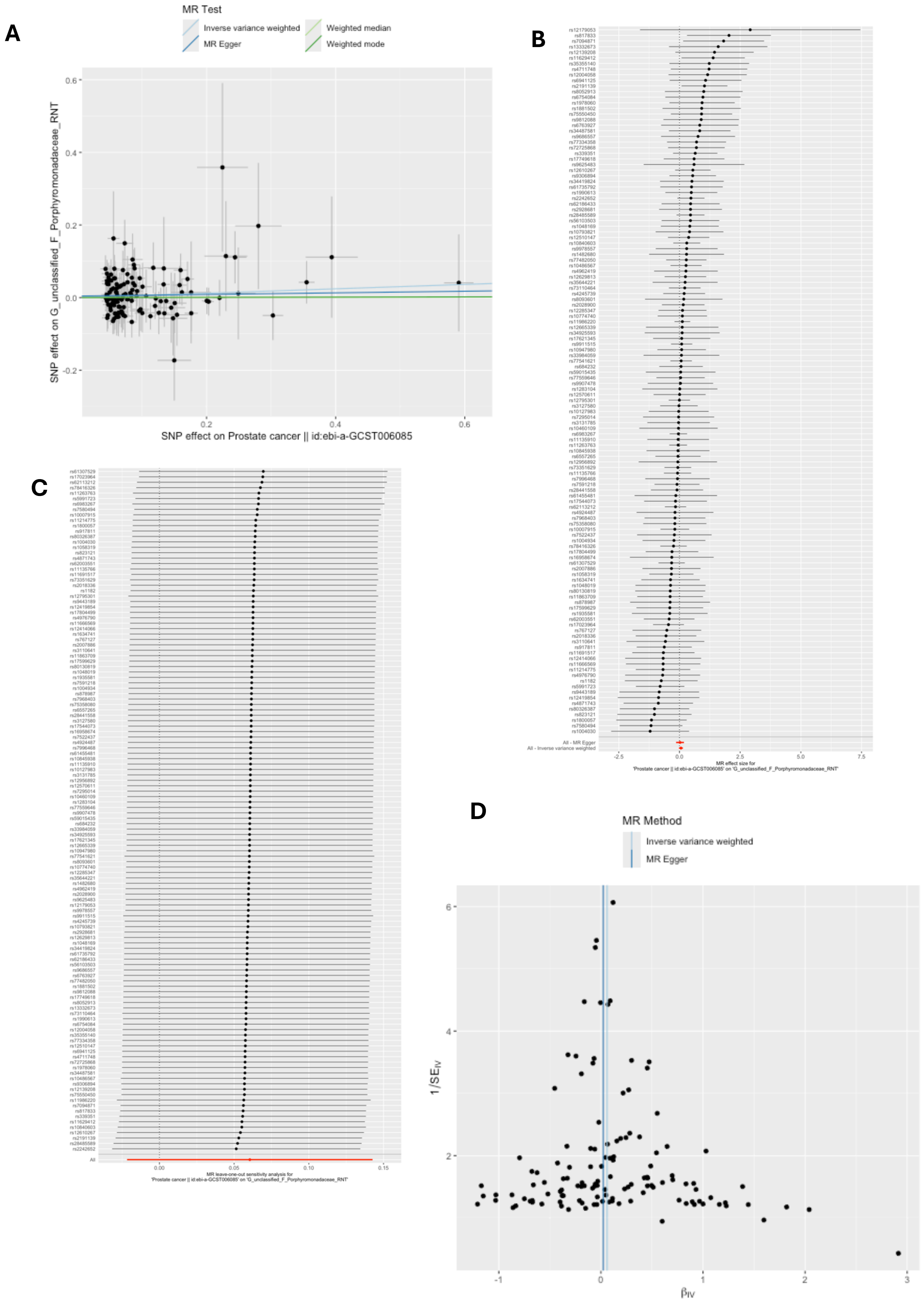

*G = genus; F = family; RNT = reverse normal transformation; MR = Mendelian randomization; SNP = single nucleotide polymorphism. These plots show the results of reverse MR analyses to test the causal effect of prostate cancer on a higher relative abundance of bacteria within the genus of unclassified family Porphyromonadaceae (G_unclassified_F_Porphyromonadaceae_RNT) using all SNPs associated with prostate cancer at a genome-wide significant P-value threshold ($P<5x10^{-8}$) in the Schumacher et al 2018 prostate cancer GWAS. A) Scatter plot comparing four MR methods; B) forest plot comparing the effect of individual SNP-level effect estimates derived using the Wald ratio method and in combination with the inverse variance weighted (IVW) and MR-Egger estimates; C) leave-one-out analysis to check if any one SNP is driving pleiotropy or asymmetry in the estimate; and D) funnel plot comparing the effect estimate and precision of each SNP-level Wald ratio estimate.*

**Supplementary Figure 62: Reverse MR results for effect of prostate cancer on G.unclassified.P.Firmicutes**

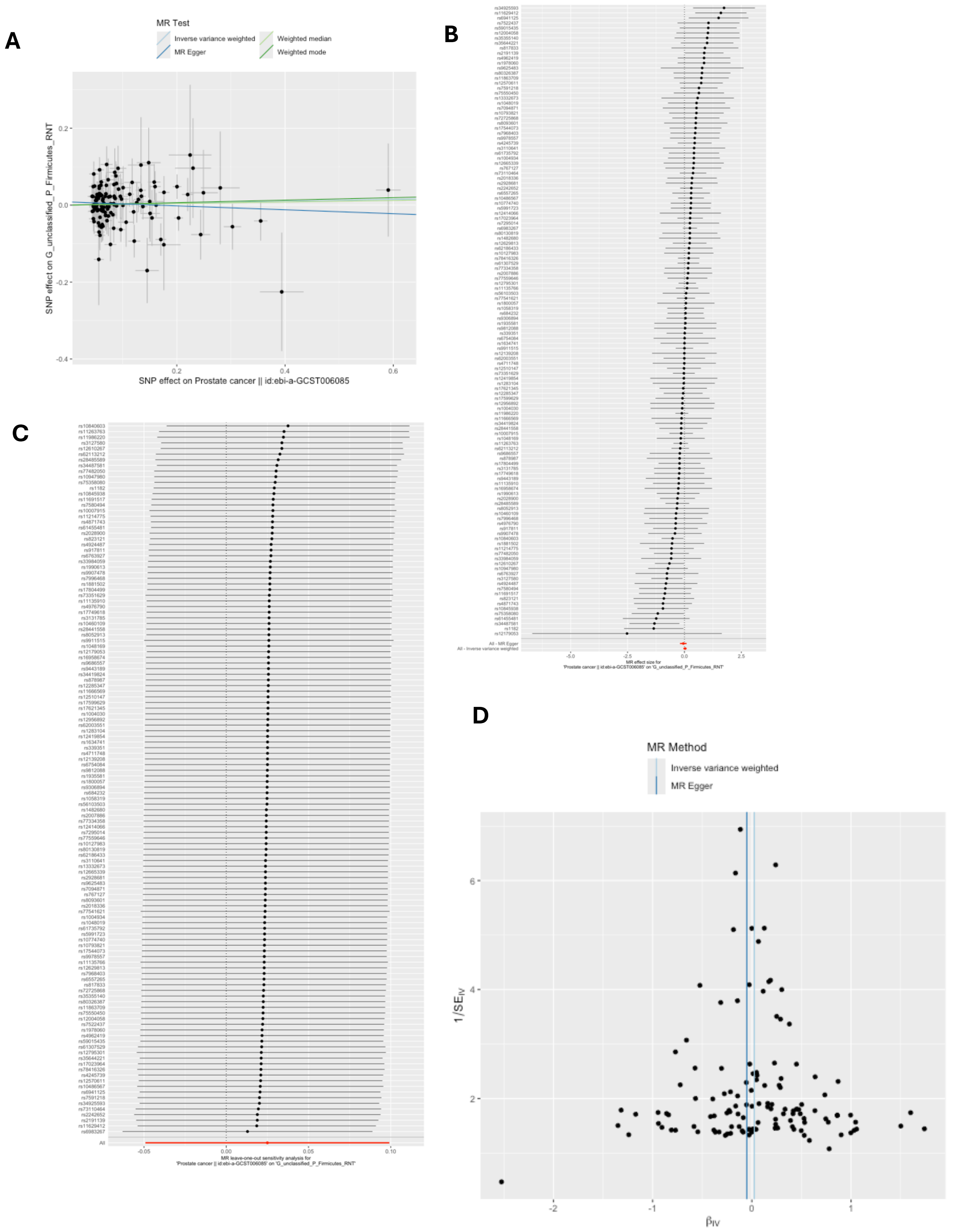


*G = genus; P = phylum; RNT = reverse normal transformation; MR = Mendelian randomization; SNP = single nucleotide polymorphism. These plots show the results of reverse MR analyses to test the causal effect of prostate cancer on a higher relative abundance of bacteria within the genus of unclassified phylum Firmicutes*

*(G_unclassified_P_Firmicutes_RNT) using all SNPs associated with prostate cancer at a genome-wide significant P-value threshold ($P<5x10^{-8}$) in the Schumacher et al 2018 prostate cancer GWAS. A) Scatter plot comparing four MR methods; B) forest plot comparing the effect of individual SNP-level effect estimates derived using the Wald ratio method and in combination with the inverse variance weighted (IVW) and MR-Egger estimates; C) leave-one-out analysis to check if any one SNP is driving pleiotropy or asymmetry in the estimate; and D) funnel plot comparing the effect estimate and precision of each SNP-level Wald ratio estimate.*

**Supplementary Figure 63: Reverse MR results for effect of ovarian cancer on G.unclassified.P.Firmicutes.HB**

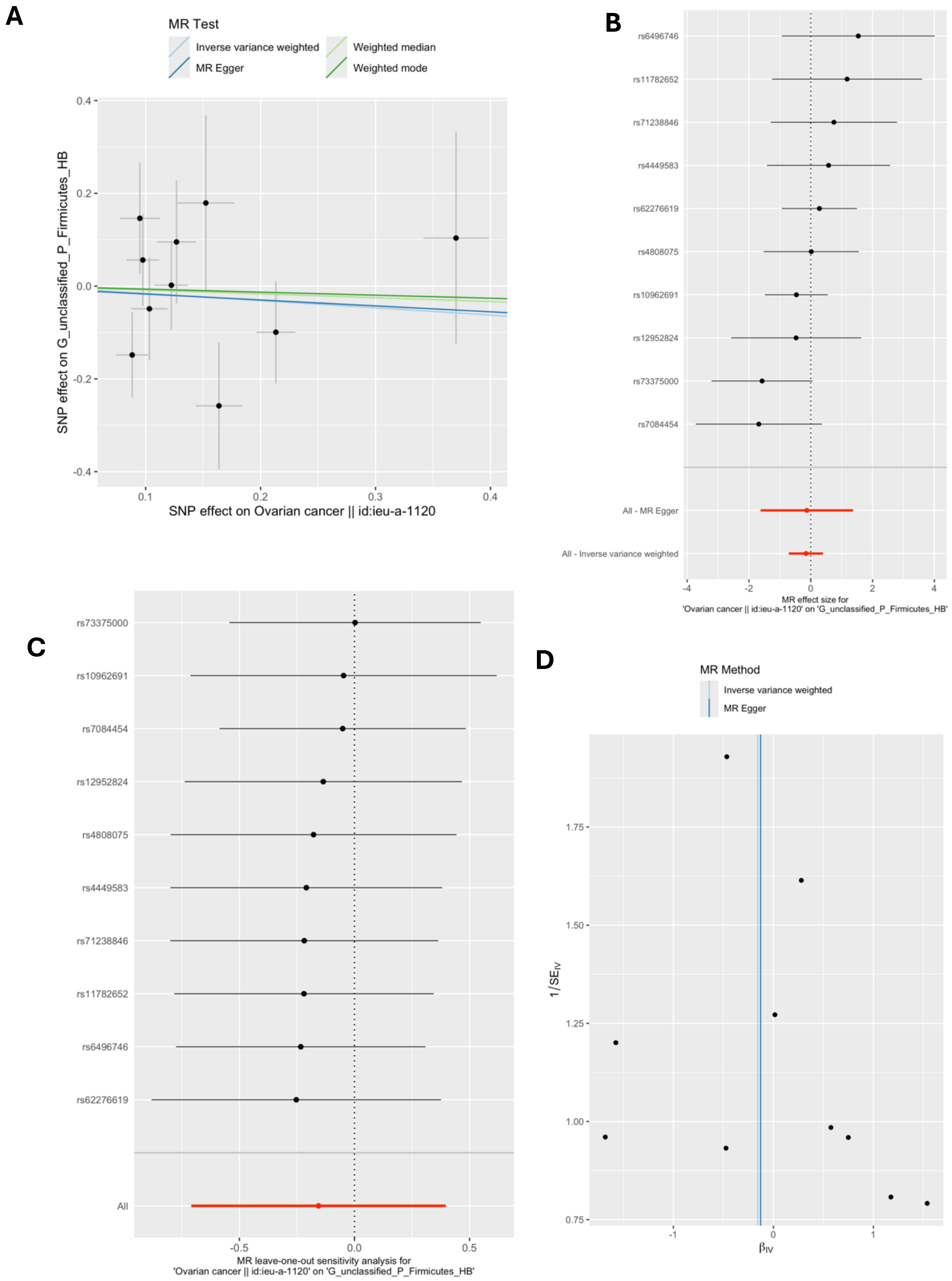


*G = genus; P = phylum; HB = hurdle binary (presence vs absence) ; MR = Mendelian randomization; SNP = single nucleotide polymorphism. These plots show the results of reverse MR analyses to test the causal effect of ovarian cancer on a higher relative abundance of bacteria within the genus of unclassified phylum Firmicutes (G_unclassified_P_Firmicutes_HB) using all SNPs associated with ovarian cancer at a genome-wide significant P-value threshold (P<5x10$^{-8}$) in the Phelan et al 2017 ovarian cancer GWAS (accessed by IEU Open GWAS ID: ieu-a-*

*1120). A) Scatter plot comparing four MR methods; B) forest plot comparing the effect of individual SNP-level effect estimates derived using the Wald ratio method and in combination with the inverse variance weighted (IVW) and MR-Egger estimates; C) leave-one-out analysis to check if any one SNP is driving pleiotropy or asymmetry in the estimate; and D) funnel plot comparing the effect estimate and precision of each SNP-level Wald ratio estimate.*

1. Hughes DA, Bacigalupe R, Wang J, Rühlemann MC, Tito RY, Falony G, et al. Genome-wide associations of human gut microbiome variation and implications for causal inference analyses. Nat Microbiol. 2020;5(9):1079-87.
2. Goodrich JK, Davenport ER, Beaumont M, Jackson MA, Knight R, Ober C, et al. Genetic Determinants of the Gut Microbiome in UK Twins. Cell Host Microbe. 2016;19(5):731-43.
3. Blekhman R, Goodrich JK, Huang K, Sun Q, Bukowski R, Bell JT, et al. Host genetic variation impacts microbiome composition across human body sites. Genome Biol. 2015;16(1):191.
4. Bonder MJ, Kurilshikov A, Tigchelaar EF, Mujagic Z, Imhann F, Vila AV, et al. The effect of host genetics on the gut microbiome. Nat Genet. 2016;48(11):1407-12.
5. Kurilshikov A, Medina-Gomez C, Bacigalupe R, Radjabzadeh D, Wang J, Demirkan A, et al. Large-scale association analyses identify host factors influencing human gut microbiome composition. Nature Genetics. 2021;53(2):156-65.
6. Fernandez-Rozadilla C, Timofeeva M, Chen Z, Law P, Thomas M, Schmit S, et al. Deciphering colorectal cancer genetics through multi-omic analysis of 100,204 cases and 154,587 controls of European and east Asian ancestries.
7. Zhang H, Ahearn TU, Lecarpentier J, Barnes D, Beesley J, Qi G, et al. Genome-wide association study identifies 32 novel breast cancer susceptibility loci from overall and subtype-specific analyses. Nat Genet. 2020;52(6):572-81.
8. O'Mara TA, Glubb DM, Amant F, Annibali D, Ashton K, Attia J, et al. Identification of nine new susceptibility loci for endometrial cancer.
9. Fryer E, Hatcher C, Knight R, Wade K. Exploring the causal role of the human gut microbiome in endometrial cancer: a Mendelian randomization approach. medRxiv. 2024:2024.03.06.24303765.
10. De Vivo I, Prescott J, Setiawan VW, Olson SH, Wentzensen N, Attia J, et al. Genome-wide association study of endometrial cancer in E2C2. Hum Genet. 2014;133(2):211-24.
11. Cheng TH, Thompson DJ, O'Mara TA, Painter JN, Glubb DM, Flach S, et al. Five endometrial cancer risk loci identified through genome-wide association analysis. Nat Genet. 2016;48(6):667-74.
12. O'Mara TA, Glubb DM, Amant F, Annibali D, Ashton K, Attia J, et al. Identification of nine new susceptibility loci for endometrial cancer.
13. Fuchsberger C, Abecasis GR, Hinds DA. minimac2: faster genotype imputation. Bioinformatics. 2015;31(5):782-4.
14. Bycroft C, Freeman C, Petkova D, Band G, Elliott LT, Sharp K, et al. The UK Biobank resource with deep phenotyping and genomic data.
15. Wang Y, McKay JD, Rafnar T, Wang Z, Timofeeva MN, Broderick P, et al. Rare variants of large effect in BRCA2 and CHEK2 affect risk of lung cancer. Nature Genetics. 2014;46(7):736-41.
16. Transdisciplinary Research Into Cancer of the Lung (TRICL) - Meta Analysis [Internet]. Available from: https://www.ncbi.nlm.nih.gov/projects/gap/cgi-bin/study.cgi?study_id=phs000877.v1.p1.
17. Schumacher FR, Al Olama AA, Berndt SI, Benlloch S, Ahmed M, Saunders EJ, et al. Association analyses of more than 140,000 men identify 63 new prostate cancer susceptibility loci.
18. Klein AP, Wolpin BM, Risch HA, Stolzenberg-Solomon RZ, Mocci E, Zhang M, et al. Genome-wide meta-analysis identifies five new susceptibility loci for pancreatic cancer.

19. Amundadottir L, Kraft P, Stolzenberg-Solomon RZ, Fuchs CS, Petersen GM, Arslan AA, et al. Genome-wide association study identifies variants in the ABO locus associated with susceptibility to pancreatic cancer. Nat Genet. 2009;41(9):986-90.
20. Petersen GM, Amundadottir L, Fuchs CS, Kraft P, Stolzenberg-Solomon RZ, Jacobs KB, et al. A genome-wide association study identifies pancreatic cancer susceptibility loci on chromosomes 13q22.1, 1q32.1 and 5p15.33. Nat Genet. 2010;42(3):224-8.
21. Wolpin BM, Rizzato C, Kraft P, Kooperberg C, Petersen GM, Wang Z, et al. Genome-wide association study identifies multiple susceptibility loci for pancreatic cancer. Nat Genet. 2014;46(9):994-1000.
22. Childs EJ, Mocci E, Campa D, Bracci PM, Gallinger S, Goggins M, et al. Common variation at 2p13.3, 3q29, 7p13 and 17q25.1 associated with susceptibility to pancreatic cancer. Nat Genet. 2015;47(8):911-6.
23. Wang Z, Zhu B, Zhang M, Parikh H, Jia J, Chung CC, et al. Imputation and subset-based association analysis across different cancer types identifies multiple independent risk loci in the TERT-CLPTM1L region on chromosome 5p15.33. Hum Mol Genet. 2014;23(24):6616-33.
24. Auton A, Abecasis GR, Altshuler DM, Durbin RM, Bentley DR, Chakravarti A, et al. A global reference for human genetic variation.
25. Howie BN, Donnelly P, Marchini J. A flexible and accurate genotype imputation method for the next generation of genome-wide association studies. PLoS Genet. 2009;5(6):e1000529.
26. Schröder J, Chegwidden L, Maj C, Gehlen J, Speller J, Böhmer AC, et al. GWAS meta-analysis of 16 790 patients with Barrett's oesophagus and oesophageal adenocarcinoma identifies 16 novel genetic risk loci and provides insights into disease aetiology beyond the single marker level. Gut. 2023;72(4):612-23.
27. Levine DM, Ek WE, Zhang R, Liu X, Onstad L, Sather C, et al. A genome-wide association study identifies new susceptibility loci for esophageal adenocarcinoma and Barrett's esophagus. Nat Genet. 2013;45(12):1487-93.
28. Gharahkhani P, Fitzgerald RC, Vaughan TL, Palles C, Gockel I, Tomlinson I, et al. Genome-wide association studies in oesophageal adenocarcinoma and Barrett's oesophagus: a large-scale meta-analysis. Lancet Oncol. 2016;17(10):1363-73.
29. Iglesias AI, van der Lee SJ, Bonnemaijer PWM, Höhn R, Nag A, Gharahkhani P, et al. Haplotype reference consortium panel: Practical implications of imputations with large reference panels. Human Mutation. 2017;38(8):1025-32.
30. Walter K, Min JL, Huang J, Crooks L, Memari Y, McCarthy S, et al. The UK10K project identifies rare variants in health and disease. Nature. 2015;526(7571):82-90.
31. Auton A, Brooks LD, Durbin RM, Garrison EP, Kang HM, Korbel JO, et al. A global reference for human genetic variation. Nature. 2015;526(7571):68-74.
32. Song H, Ramus SJ, Tyrer J, Bolton KL, Gentry-Maharaj A, Wozniak E, et al. A genome-wide association study identifies a new ovarian cancer susceptibility locus on 9p22.2. Nat Genet. 2009;41(9):996-1000.
33. Phelan CM, Kuchenbaecker KB, Tyrer JP, Kar SP, Lawrenson K, Winham SJ, et al. Identification of 12 new susceptibility loci for different histotypes of epithelial ovarian cancer. Nature Genetics. 2017;49(5):680-91.
34. Pharoah PD, Tsai YY, Ramus SJ, Phelan CM, Goode EL, Lawrenson K, et al. GWAS meta-analysis and replication identifies three new susceptibility loci for ovarian cancer. Nat Genet. 2013;45(4):362-70, 70e1-2.
35. Burrows K, Haycock P. Genome-wide Association Study of Cancer Risk in UK Biobank. 2021 [Available from: https://data.bris.ac.uk/data/dataset/aed0u12w0ede20olb0m77p4b9.

36. Ruth E Mitchell, Gibran Hemani, Tom Dudding, Laura Corbin, Sean Harrison, Paternoster L. UK Biobank Genetic Data: MRC-IEU Quality Control, version 2, 18/01/2019. 2019.
37. Kurki MI, Karjalainen J, Palta P, Sipila TP, Kristiansson K, Donner KM, et al. FinnGen provides genetic insights from a well-phenotyped isolated population. Nature. 2023;613(7944):508-18.
38. Elsworth B, Lyon M, Alexander T, Liu Y, Matthews P, Hallett J, et al. The MRC IEU OpenGWAS data infrastructure. bioRxiv. 2020:2020.08.10.244293.
39. Michailidou K, Lindström S, Dennis J, Beesley J, Hui S, Kar S, et al. Association analysis identifies 65 new breast cancer risk loci. Nature. 2017;551(7678):92-4.
40. Huyghe JR, Bien SA, Harrison TA, Kang HM, Chen S, Schmit SL, et al. Discovery of common and rare genetic risk variants for colorectal cancer.
41. Davey Smith G, Hemani G. Mendelian randomization: genetic anchors for causal inference in epidemiological studies. Hum Mol Genet. 2014;23(R1):R89-98.
42. Haycock PC, Burgess S, Wade KH, Bowden J, Relton C, Davey Smith G. Best (but oft-forgotten) practices: the design, analysis, and interpretation of Mendelian randomization studies. Am J Clin Nutr. 2016;103(4):965-78.
43. Hemani G, Tilling K, Davey Smith G. Orienting the causal relationship between imprecisely measured traits using GWAS summary data. PLOS Genetics. 2017;13(11):e1007081.
44. Hemani G, Zheng J, Elsworth B, Wade KH, Haberland V, Baird D, et al. The MR-Base platform supports systematic causal inference across the human phenome. Elife. 2018;7.
45. Hatcher C, Richenberg G, Waterson S, Nguyen LH, Joshi AD, Carreras-Torres R, et al. Application of Mendelian randomization to explore the causal role of the human gut microbiome in colorectal cancer. Scientific Reports. 2023;13(1):5968.
46. Kurilshikov A, Medina-Gomez C, Bacigalupe R, Radjabzadeh D, Wang J, Demirkan A, et al. Large-scale association analyses identify host factors influencing human gut microbiome composition. Nature Genetics. 2021;53(2):156-65.
47. Kurilshikov A. MiBioGen miQTL pipeline. 2018.
48. Giambartolomei C, Vukcevic D, Schadt EE, Franke L, Hingorani AD, Wallace C, et al. Bayesian test for colocalisation between pairs of genetic association studies using summary statistics. PLoS Genet. 2014;10(5):e1004383.
49. Lee M. mattlee821 / functions. 2024.
50. Hemani G, Elsworth B, Palmer T, Rasteiro R. *ieugwasr: Interface to the 'OpenGWAS' Database API* . R package version 1.0.2, https://mrcieu.github.io/ieugwasr/. 2024.
51. Sanna S, van Zuydam NR, Mahajan A, Kurilshikov A, Vich Vila A, Võsa U, et al. Causal relationships among the gut microbiome, short-chain fatty acids and metabolic diseases. Nat Genet. 2019;51(4):600-5.
52. Wang J, Kurilshikov A, Radjabzadeh D, Turpin W, Croitoru K, Bonder MJ, et al. Meta-analysis of human genome-microbiome association studies: the MiBioGen consortium initiative. Microbiome. 2018;6(1):101.
53. Bowden J, Davey Smith G, Haycock PC, Burgess S. Consistent Estimation in Mendelian Randomization with Some Invalid Instruments Using a Weighted Median Estimator. Genet Epidemiol. 2016;40(4):304-14.
54. Hartwig FP, Davey Smith G, Bowden J. Robust inference in summary data Mendelian randomization via the zero modal pleiotropy assumption. Int J Epidemiol. 2017;46(6):1985-98.
55. Bowden J, Davey Smith G, Burgess S. Mendelian randomization with invalid instruments: effect estimation and bias detection through Egger regression. Int J Epidemiol. 2015;44(2):512-25.

56. Burgess S, Butterworth A, Thompson SG. Mendelian randomization analysis with multiple genetic variants using summarized data. Genet Epidemiol. 2013;37(7):658-65.
57. Burgess S, Labrecque JA. Mendelian randomization with a binary exposure variable: interpretation and presentation of causal estimates. Eur J Epidemiol. 2018;33(10):947-52.
58. Davies NM, Holmes MV, Davey Smith G. Reading Mendelian randomisation studies: a guide, glossary, and checklist for clinicians. BMJ. 2018;362:k601.
