## Supplementary material for "Application of Mendelian randomization to appraise causality in relationships between the gut microbiome and cancer": STROBE-MR checklist

**STROBE-MR checklist of recommended items to address in reports of Mendelian randomization studies**[1 2]

| Item No. | Section | Checklist item | Page No. | Relevant text from manuscript |
|---|---|---|---|---|
| 1 | **TITLE and ABSTRACT** | Indicate Mendelian randomization (MR) as the study's design in the title and/or the abstract if that is a main purpose of the study | 1-2 | Title: "Application of Mendelian randomization to appraise causality of the relationship between the gut microbiome and cancer". Abstract "Here, we aimed to appropriately apply MR to understand the role of the gut microbiome in cancer aetiology…" (p.1). details the purpose for application of MR in this study. |
| | **INTRODUCTION** | | | |
| 2 | **Background** | Explain the scientific background and rationale for the reported study. What is the exposure? Is a potential causal relationship between exposure and outcome plausible? Justify why MR is a helpful method to address the study question | 1-4 | The rationale for the study is provided, and the exposure and outcomes stated and rationale for these provided. Previous studies have reported a causal relationship between gut microbial traits and cancer, for which we have explained the purpose of the study was to "use two-sample MR to estimate the causal relationship between the gut microbiome and cancer using some of the largest GWASs to date", and "demonstrate the utility of a series of sensitivity analyses, also introduced in our previous work, which test the robustness of findings to violations of MR assumptions, in appraising causal evidence for observed estimates." See Cancer is among the leading causes of death worldwide… Previous work has implicated a role of the gut microbiome in cancer aetiology; however, its causal relevance is not clear." (p.1). "Mendelian randomisation (MR) is a method that offers improved causal inference…" (p.2). |
| 3 | **Objectives** | State specific objectives clearly, including pre-specified causal hypotheses (if any). State that MR is a method that, under specific assumptions, intends to estimate causal effects | 2-3 | Objectives clearly outlined in the last three sentences of the introduction. "We aimed to appropriately apply MR to understand the role of the gut microbiome in cancer aetiology, whilst demonstrating the range of sensitivity analyses required…" (p.2–3). |
| | **METHODS** | | | |

| | | | | |
|---|---|---|---|---|
| 4 | **Study design and data sources** | Present key elements of the study design early in the article. Consider including a table listing sources of data for all phases of the study. For each data source contributing to the analysis, describe the following: | 22-23 | A summary of the study design is provided on page 4 with the cancer outcomes data sources overview being provided in Supplementary Table 4, and Supplementary material pages 1-14 provide detailed information on the cancer GWAS included in the study. Information on the MiBioGen and Hughes microbiome data are provided on pages 4-6, and details of the included cohorts within the MiBioGen consortia are included in Supplementary Table 3. |
| | a) | Setting: Describe the study design and the underlying population, if possible. Describe the setting, locations, and relevant dates, including periods of recruitment, exposure, follow-up, and data collection, when available. | 22 | Two-sample MR was used to examine the causal relationship between features of the human gut microbiome and overall cancer risk…" Supplementary material pages 1-14 provide information on the study. |
| | b) | Participants: Give the eligibility criteria, and the sources and methods of selection of participants. Report the sample size, and whether any power or sample size calculations were carried out prior to the main analysis | 23 | "Genetic variants associated with microbial traits were obtained from two of the largest microbiome GWASs…" (p.21). "FGFP (n = 2,223), FoCus (n = 950), PopGen (n = 717)" (p.21–22). |
| | c) | Describe measurement, quality control and selection of genetic variants | 3-4, 21-22 | Information on genetic measurements and selection of instruments from the exposure GWAS are provided in pages 3-4. Quality control of genetic variants has been previously described in literature and this is referenced in the text. "Information on the microbial traits analysed and SNPs selected as genetic instruments… Effect estimates… represent the SD change for rank normalised relative abundance…" (p.21–22). The selection of variants is also described in Supplementary Table |
| | d) | For each exposure, outcome, and other relevant variables, describe methods of assessment and diagnostic criteria for diseases | 22-23 | Measurements of the exposures and outcomes are detailed throughout supplementary methods pages 1-16. |
| | e) | Provide details of ethics committee approval and participant informed consent, if relevant | 27 | Not relevant in this study. |
| 5 | **Assumptions** | Explicitly state the three core IV assumptions for the main analysis (relevance, independence and exclusion restriction) as well assumptions for any additional or sensitivity analysis | 24-25 | These are stated in Supplementary Methods pages 20 and in Figure 1 page 39. |

| | | | | | |
|---|---|---|---|---|---|
| 6 | **Statistical methods: main analysis** | | Describe statistical methods and statistics used | 24-26 | Details of tests incluing Wald ratio method, IVW, weighted mean and weighted median are provided. Further details of statistics methods are provided in Supplementary Methods pages 16-26. |
| | | a) | Describe how quantitative variables were handled in the analyses (i.e., scale, units, model) | 21-22 | Units and transformations are provided for all quantitative analyses, as well as how these estimates can be interpreted. |
| | | b) | Describe how genetic variants were handled in the analyses and, if applicable, how their weights were selected | 23-24 | Weights are not provided (as this is a two-sample MR) but the methodology for MR effect estimation are provided (i.e., the Wald ratio/IVW for main analyses). See Supplementary Methods pages 16-17. Microbial trait units are discussed on pages 20-21, and also provided on page 4. |
| | | c) | Describe the MR estimator (e.g. two-stage least squares, Wald ratio) and related statistics. Detail the included covariates and, in case of two-sample MR, whether the same covariate set was used for adjustment in the two samples | 24-26 | The methodology for MR effect estimation are provided (i.e., the Wald ratio and the inverse variance weighted method) as well as how the F-statistics and R-squared are calculated. |
| | | d) | Explain how missing data were addressed | 25 | "Non-inferable SNPs… were removed…" (p.25). |
| | | e) | If applicable, indicate how multiple testing was addressed | 28 | "We did not correct for multiple testing… P-values were interpreted as continuous indicators of evidence strength." (p.28). |
| 7 | **Assessment of assumptions** | | Describe any methods or prior knowledge used to assess the assumptions or justify their validity | 24-26 | "Colocalisation was used… A phenome-wide analysis was performed…" (p.24–26). |
| 8 | **Sensitivity analyses and additional analyses** | | Describe any sensitivity analyses or additional analyses performed (e.g. comparison of effect estimates from different approaches, independent replication, bias analytic techniques, validation of instruments, simulations) | P25-26 | "Sensitivity analyses included… European-only analyses… colocalisation… pleiotropy exploration… Steiger tests… reverse MR analyses." (p.25–26). |
| 9 | **Software and pre-registration** | | | p.23, 27 | We did not preregister this study as this was not deemed necessary given that we had consent from authors where required to perform the analysis using pre-existing GWAS data. |

| | | | | | |
|---|---|---|---|---|---|
| | | a) | Name statistical software and package(s), including version and settings used | p.23, | "Two-sample MR using the TwoSampleMR package… in R (version 4.4.1)." (p.23). |
| | | b) | State whether the study protocol and details were pre-registered (as well as when and where) | P27 | No preregistration noted |
| | **RESULTS** | | | | |
| 10 | **Descriptive data** | | | | |
| | | a) | Report the numbers of individuals at each stage of included studies and reasons for exclusion. Consider use of a flow diagram | 4-5 | The numbers of individuals in the exposure datasets are presented in the 'Gut microbiome GWAS data and instrument selection' in the third and fourth paragraph in the sections of the methods, and provided in Supplementary Table 1-3. The numbers of individuals in the outcome datasets are presented in the Supplementary Material/ Supplementary Methods 'Data' section. |
| | | b) | Report summary statistics for phenotypic exposure(s), outcome(s), and other relevant variables (e.g. means, SDs, proportions) | 4-7 | These are not provided by the individual genome-wide association studies. Numbers of cases and controls for the outcome are provided in Supplementary Table S4 |
| | | c) | If the data sources include meta-analyses of previous studies, provide the assessments of heterogeneity across these studies | P23-24 | "Cochran's Q test…" (p.23–24). |
| | | d) | For two-sample MR:<br>i. Provide justification of the similarity of the genetic variant-exposure associations between the exposure and outcome samples<br>ii. Provide information on the number of individuals who overlap between the exposure and outcome studies | P23-24 | "Exposure and outcome datasets were harmonised… proxy SNPs used…" (p.23–24). |
| 11 | **Main results** | | | | |
| | | a) | Report the associations between genetic variant and exposure, and between genetic variant and outcome, preferably on an interpretable scale | 4-7 | "There was evidence for five causal effects… OR: 0.84… OR: 1.17…" (p.4–5). "There was evidence for 18 causal effects…" (p.5–7). |
| | | b) | Report MR estimates of the relationship between exposure and outcome, and the measures of uncertainty from the MR analysis, on an interpretable scale, such as odds ratio or relative risk per SD difference | | "There was evidence for five causal effects… OR: 0.84… OR: 1.17…" (p.4–5). "There was evidence for 18 causal effects…" (p.5–7). |

| | | | | | |
|---|---|---|---|---|---|
| | | c) | If relevant, consider translating estimates of relative risk into absolute risk for a meaningful time period | | Not perceived relevant for the purposes of this study. |
| | | d) | Consider plots to visualize results (e.g. forest plot, scatterplot of associations between genetic variants and outcome versus between genetic variants and exposure) | P41-42 | Plots are provided on pages 41-42. |
| 12 | **Assessment of assumptions** | | | P8-10 | “Colocalisation analyses… only four provided more evidence for colocalisation (H4)… many regions suggested multiple outcome-associated SNPs…” (p.9–10). |
| | | a) | Report the assessment of the validity of the assumptions | P19-22 | Discussed on pages 19-22 |
| | | b) | Report any additional statistics (e.g., assessments of heterogeneity across genetic variants, such as $I^2$, Q statistic or E-value) | | These statistics are not reported here in this paper as these were only possible with reverse MR and sensitivity analyses using a lenient P1x10-5 threshold, but were carried out in this study, an can be made available if required. |
| 13 | **Sensitivity analyses and additional analyses** | | | | |
| | | a) | Report any sensitivity analyses to assess the robustness of the main results to violations of the assumptions | 7-13, 24-27 | Sensitivity analyses are provided across pages 7-13, and methods pages |
| | | b) | Report results from other sensitivity analyses or additional analyses | P7-12 | Results from a variety of sensitivity analyses are provide from pages 7-12, and in Supplementary material |
| | | c) | Report any assessment of direction of causal relationship (e.g., bidirectional MR) | P13 | Reverse MR analyses and steiger filtering analyses have been conducted, and results are presented in Supplementary Material and in the main text on page 13. |
| | | d) | When relevant, report and compare with estimates from non-MR analyses | | Given this was a entirely focused MR paper, and the results are subjected to poteintial bias which id discusse in the paper, non-MR analyses were not conducte in this study. |
| | | e) | Consider additional plots to visualize results (e.g., leave-one-out analyses) | Supplementary | Plots for Sensitivity analyses are provided in Supplementary Material and referenced in the main text. |

| | | | | | |
|---|---|---|---|---|---|
| | **DISCUSSION** | | | | |
| 14 | **Key results** | | Summarize key results with reference to study objectives | P14-18 | See pages 14-18 Findings, and sensitivity analyses results discussed, in relation to MR assumptions, and in contect of wider literature. Study stengths are provided on page 19 |
| 15 | **Limitations** | | Discuss limitations of the study, taking into account the validity of the IV assumptions, other sources of potential bias, and imprecision. Discuss both direction and magnitude of any potential bias and any efforts to address them | | |
| 16 | **Interpretation** | | | | |
| | | a) | Meaning: Give a cautious overall interpretation of results in the context of their limitations and in comparison with other studies | | A cautious interpretation is provided in the discussion throughout pages 14-22. |
| | | b) | Mechanism: Discuss underlying biological mechanisms that could drive a potential causal relationship between the investigated exposure and the outcome, and whether the gene-environment equivalence assumption is reasonable. Use causal language carefully, clarifying that IV estimates may provide causal effects only under certain assumptions | | There is no need for an explanation or exploration of mechanism due to the likely complexity of the results. |
| | | c) | Clinical relevance: Discuss whether the results have clinical or public policy relevance, and to what extent they inform effect sizes of possible interventions | | This study does not directly inform clinical decisions, nor public policy. However, the key findings suggest that further research is required before clinical decisions or public policy decisions can be made on the application of modifying aspects or features of the gut microbiome to treat or prevent cancer. |
| 17 | **Generalizability** | | Discuss the generalizability of the study results (a) to other populations, (b) across other exposure periods/timings, and (c) across other levels of exposure | | Results in our study our discussed with those presented in other studies, and similarities and discrepencies are discussed, along with justification as to why this might be the case. Generalisability and recommendations for future studies are provided throughout the disciussion in pages 14-22. |
| | **OTHER INFORMATION** | | | | |
| 18 | **Funding** | | Describe sources of funding and the role of funders in the present study and, if applicable, sources of funding for the databases and original study or studies on which the present study is based | | See page 29 |

| | | | |
|---|---|---|---|
| 19 | **Data and data sharing** | Provide the data used to perform all analyses or report where and how the data can be accessed, and reference these sources in the article. Provide the statistical code needed to reproduce the results in the article, or report whether the code is publicly accessible and if so, where | See page 28 |
| 20 | **Conflicts of Interest** | All authors should declare all potential conflicts of interest | See page 29 |

This checklist is copyrighted by the Equator Network under the Creative Commons Attribution 3.0 Unported (CC BY 3.0) license.

1. Skrivankova VW, Richmond RC, Woolf BAR, Yarmolinsky J, Davies NM, Swanson SA, et al. Strengthening the Reporting of Observational Studies in Epidemiology using Mendelian Randomization (STROBE-MR) Statement. JAMA. 2021;under review.

2. Skrivankova VW, Richmond RC, Woolf BAR, Davies NM, Swanson SA, VanderWeele TJ, et al. Strengthening the Reporting of Observational Studies in Epidemiology using Mendelian Randomisation (STROBE-MR): Explanation and Elaboration. BMJ. 2021;375:n2233.
